# Retrospective analysis of *The Two Sister Study* using haplotype-based association testing to identify loci associated with early-onset breast cancer

**DOI:** 10.1101/2020.12.04.20244251

**Authors:** James R. Gilbert, James J. Cray, Joseph E. Losee, Gregory M. Cooper

**Affiliations:** Department of Plastic Surgery, University of Pittsburgh/Children’s Hospital of Pittsburgh, Pittsburgh, PA 15201; Division of Anatomy, The Ohio State University College of Medicine, Columbus, OH 43210; Department of Oral Biology, University of Pittsburgh/Children’s Hospital of Pittsburgh, Pittsburgh, PA 15201; Department of Bioengineering, University of Pittsburgh/Children’s Hospital of Pittsburgh, Pittsburgh, PA 15201

**Keywords:** young-onset, early-onset, cancer, familial, breast cancer

## Abstract

Breast cancer is a polygenic disorder and is the leading cause of cancer related mortality among women. Early-onset breast cancer (EOBC) is diagnosed in women prior to 45 years-of-age and is associated with worse clinical outcomes, a more aggressive disease phenotype, and poor prognosis for disease-free survival. While substantial progress has been made in defining the genetics of breast cancer, EOBC remains less well understood. In the current study we perform a retrospective analysis of data derived from *The Two Sister Study*. The use of alternate strategies for handling age-at-diagnosis in conjunction with haplotype-based methods yielded novel findings that help to explain the heritability of EOBC. These findings are validated through comparison against discordant sibs from *The Two Sister Study* as well as using data derived The Cancer Genome Atlas (TCGA).

## INTRODUCTION

Breast cancer is the most frequently diagnosed oncogenic malignancy and a leading cause of cancer-related mortality among women worldwide (*1, 2*). Early-onset breast cancer (EOBC) accounts for approximately 5-10% of all new female breast cancer cases and young age at diagnosis correlates with worse clinical outcomes (*3, 4*). Germline variants play a prominent role in the etiology of breast cancer and an estimated 10-15% of women who develop breast cancer report a familial history of the disease. Germline variants in *BRCA1* or *BRCA2* are observed in 15-20% of familial breast cancer cases (*5*). Direct evidence for genetic modifiers of breast and ovarian cancer risk for *BRCA1* and *BRCA2* mutation carriers has been provided through genome-wide association study (GWAS) (*6*). Patients affected by EOBC exhibit shared patterns of gene expression that differ from their older counterparts (*7*). These combined observations suggest a genetic component contributes to EOBC although only a fraction of the heritability of EOBC has been explained.

Deciphering the genetic basis for phenotypic heterogeneity in complex diseases remains a major challenge. Single marker association studies often lack sufficient statistical power to support the discovery of rare variants or epistatic interactions within a polygenic architecture. Haplotype-based analysis is thought to have greater power than single marker association tests in the study of complex disease (*8-10*). Haplotypes, which consist of a series of sequentially ordered single nucleotide variants (SNVs), are a potentially more informative format for association testing than single markers and may have improved sensitivity and specificity for discovery (*11, 12*). Haplotype-based analysis has been used to gain insight in a wide array of complex disease models including mood disorders, multiple sclerosis, orofacial clefting, and cancer (*13-18*). Moreover, haplotype-based analysis has been effectively applied to investigate age-of-onset in human disease although relatively few studies have specifically addressed EOBC (*19-23*).

Several approaches have been used to investigate the genetic regulation of breast cancer age-of-onset. *The Two Sister Study* made use of a familial case-control design with affected cases diagnosed ≤ 50 years-of-age and discordant sibs of EOBC patients defining a control population. Parental samples were included in *The Two Sister Study* to allow for the identification of maternally-mediated effects and Mendelian errors in transmission (*24-26*). Other studies have instead used categorical thresholding with diagnosis at 35, 40, 45, and 50-years-of-age to define EOBC populations contrasted against either unaffected familial controls or unrelated age-matched controls (*3, 27-29*). Age-of-onset has further been evaluated in terms of phenotypic extremes by comparing individuals diagnosed at ≤ 35 years-of-age against cancer-free controls at age ≥ 60 or against an age-specific cohort diagnosed with breast cancer at ≥ 65 years-of-age (*30, 31*). Still others have investigated breast cancer in terms of age-stratified risk or using quantitative trait analysis to support discovery. Internally consistent logic has justified the use of these and other study designs. Yet the genetic basis for EOBC remains poorly understood and more recent studies have turned towards meta-analyses aimed at achieving sufficient statistical power to identify rare variants with small effect size (*32-34*).

Design considerations for the study of complex polygenic disorders have been evaluated across a range of disease models. For example, Peyrot and colleagues have convincingly argued against familial trio designs when investigating complex disease traits with a polygenic architecture or a lifetime risk ≥ 1% (*35*). Reasons given included a potential for reduced statistical power, ascertainment bias, and a significant underestimation of SNV heritability. Additional considerations in sib pair study design include the potential for misclassification and/or overmatching (*36*). Misclassification of discordant sibs presents a challenge primarily in cases associated with pronounced variation in age-of-onset. Overmatching presents a more significant challenge in complex disease models where discordant sibs are likely to share an indeterminate number of disease-related alleles. As a result, allele-frequency differences between affected and unaffected sibs are generally underestimated relative to randomly selected affected and unaffected individuals (*36*). Recent investigation of polygenic risk in multiplex melanoma families indicated that familial controls may carry a significantly elevated polygenic load relative to unrelated cases or healthy controls and thereby introduce bias (*37*). Kerber and colleagues likewise argued that familial studies should be approached with caution, particularly when investigating complex diseases such as cancer where variable onset, incomplete genetic penetrance, gene-environment interactions, and environmental phenocopies have a dramatic potential to impact disease occurrence and phenotype (*38*). The authors further argued in favor of a case-only analysis for an initial scan followed by more comprehensive analysis of regions surrounding initial hits using both affected and unaffected study participants. In keeping with this reasoning, we speculated that a comparison of younger and older patients diagnosed with breast cancer might provide insight into the genetic architecture of breast cancer age-of-onset.

We performed a retrospective analysis of “*The Two Sister Study: A Family-Based Study of Genes and Environment in Young-Onset Breast Cancer*,” hereafter referred to as “*The Two Sister Study*.” *The Two Sister Study* is one of the longest standing and best characterized studies of early-onset breast cancer and hence was chosen to establish proof-of-principle. Initial screening was performed using a case-only design and haplotype-based association testing while treating age-at-diagnosis as a categorical variable. Candidate regions identified through this initial screen were subsequently evaluated against discordant sibs defined within *The Two Sister Study* by variance partition analysis and haplotype-trend regression. Findings were validated using data derived from phase III of the 1000 Genomes Project and mutation and The Cancer Genome Atlas (TCGA).

## RESULTS

Our objective in this study was to investigate the genetic basis for EOBC. Access to *The Two Sister Study* was obtained through the Database of Genotypes and Phenotypes (dbGAP; accession phs000678.v1.p1). The demographics of the study population have been described elsewhere (*25, 26, 39*). The original study compared patients affected by young-onset breast cancer (age-at-diagnosis ≤ 50) with familial controls using a case-control design and affected status as a binary outcome. Pertinent populations for the purpose of this study included 1,456 cases affected by breast cancer and 525 discordant sibs.

For initial screening, the affected population was dichotomized by virtue of age-at-diagnosis using statistical modules within the %findcut SAS macro. The %findcut macro calculates thresholds for the dichotomization of continuous variables and plots a local linear regression (LOESS) curve which may be used to determine whether dichotomization is appropriate for the variable in question (*40*). While a continuous trait would be expected to produce a linear trend line with a slope ≅0, the LOESS curve generated while analyzing *The Two Sister Study* case population failed to meet the assumption of linearity **(Supplemental Fig S1)**. The observed slope and steep bend in the LOESS curve exhibited characteristics of a categorical variable justifying dichotomization of the affected population based upon age-at-diagnosis. Theoretical cutpoints were calculated using the %findcut macro and the mean value was used to distinguish between younger (diagnosis ≤ 45 years-of-age; N = 735) and older (diagnosis > 45 years-of-age; N = 721) populations.

Candidate prioritization initially involved a comparison of the younger and older affected populations using haplotype-based association testing with an expectation-maximization (EM) algorithm and a dynamic window size of 10 kilobases (kb) **(Fig 1a)**. Quantile-quantile plotting verified that the resulting data was normally distributed **(Supplemental Fig S2)**. Several peaks were observed by Manhattan plot **(Fig 1a)** with 6 haplotypes located at chromosome 6: 111,936,275-111,964,664 surpassing the threshold for genome-wide significance (p ≤ 5 x 10^−8^). This preliminary analysis identified 762 haploblocks representing 4,126 haplotypes **(Table 1)**. Upon filtering using p > 5 x 10^−4^ as a threshold for exclusion, 322 haplotypes within 282 haploblocks spanning 64 discrete autosomal regions were retained. Of these 64 regions, 15 were associated with a single haploblock and 49 included two or more adjacent haploblocks with block clusters ranging from 2-14 haploblocks in length.

**Table 1.** Summary representation of data in terms of methodology. **Sequential application of methods and filters defines a short list of candidates which may associate with breast cancer age-of-onset**. HTR = Haplotype trend regression; SNVs = single nucleotide variants retained at each stage of analysis; Haplotypes = haplotypes retained at each stage of analysis; Haploblocks = haploblocks retained at each stage of analysis; CHR RoI = chromosomal regions defined by the remaining haploblocks; RoI = regions of interest retained after analysis; * indicates region of interest was evaluated by visual assessment of data representation as opposed to statistical measures. ND = Not determined.

| **Screening method** | **SNVs** | **Haplotypes** | **Haploblocks** | **CHR Rol** | **Rol** |
| --- | --- | --- | --- | --- | --- |
| **Haplotype (10 kb)** | 762 | 4126 | 762 | ND | ND |
| **10<sup>-4</sup> filter** | 415 | 322 | 282 | 64 | ND |
| **2-6 SNV windows** | 417 | 466 | 165 | 33 | 13* |
| **HTR** | 417 | 466 | 165 | 33 | 33 |
| **Permuted p ≤ 0.05** | 64 | 14 | 14 | 14 | 14 |

**Figure 1.**
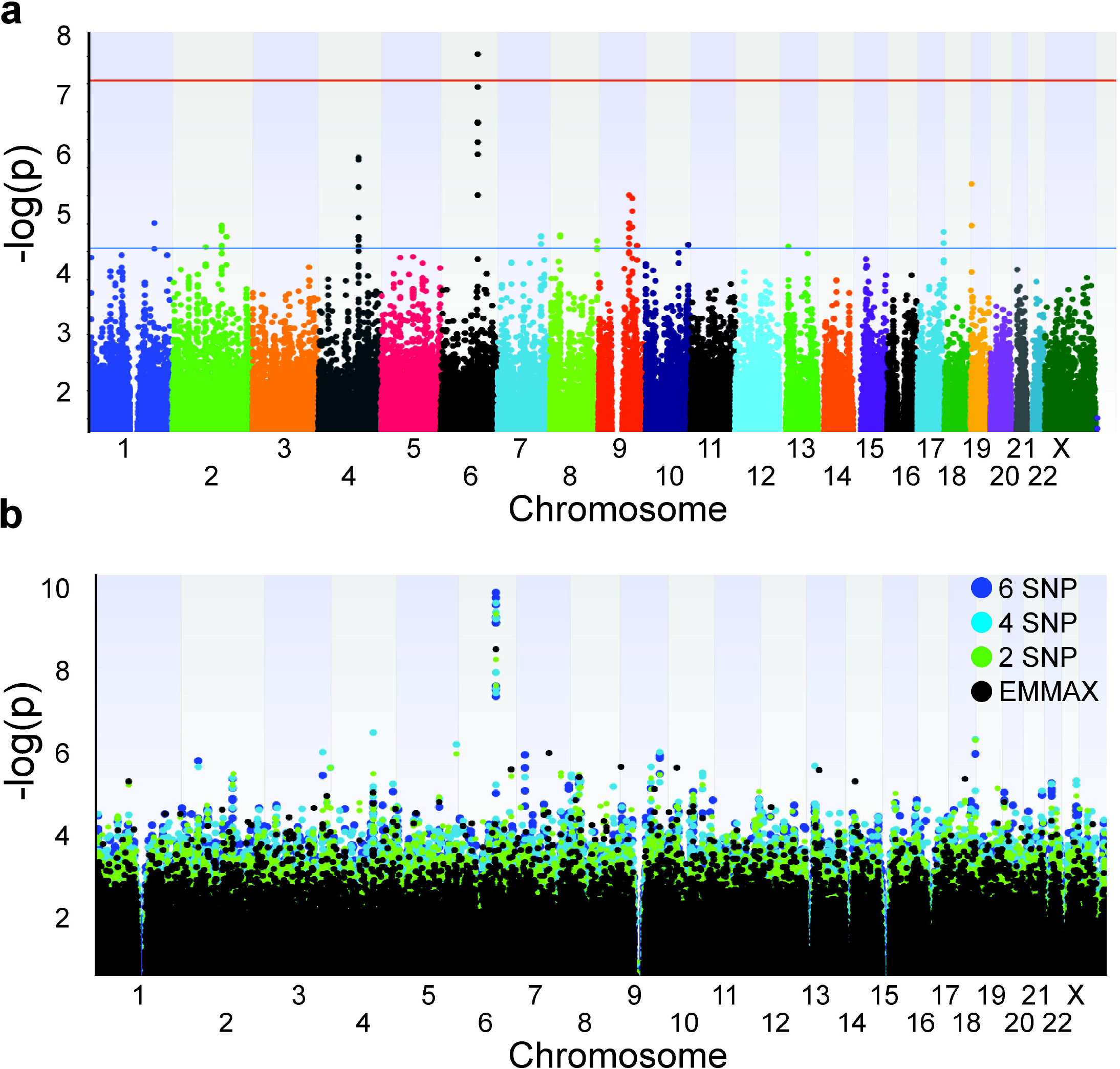
Manhattan plot. Haplotype-based analysis was performed using a fixed window size of 10 kb and an expectation-maximization (EM) algorithm with a maximum of 50 iterations. P values were derived from χ^2^. A Manhattan plot was constructed by plotting -log[p] against chromosomal position. The horizontal blue line corresponds to a suggestive threshold of p ≤ 5 x 10^−5^. The horizontal red line corresponds to the conventional threshold for genome-wide significance at p ≤ 5 x 10^−8^.

Fine mapping of the aforementioned chromosomal regions was performed using haplotype-based association testing and sliding windows of 2-6 SNVs in length as previously described by Mathias *et al*. (*41*). Assuming a panel of 684,126 variants and 3,420,615 independent tests across all windows a Bonferroni corrected threshold for genome-wide significance was calculated as p ≤ 2.92 x 10^−8^ using the method described by Song *et al* (*42*). A single-locus mixed model analysis was performed for comparison using an identity-by-state kinship matrix and an Efficient Mixed-Model Association eXpedited (EMMAX) algorithm as implemented in Sequence Variation Suite software (Golden Helix, Bozeman, MO). To facilitate direct comparison of single marker and haplotype-based analyses, Manhattan plots were overlaid **(Fig 1b)**. Only windows of 2, 4, and 6 SNVs in length were included within the composite plot for visual clarity. The composite image indicated haplotype-based testing generally outperformed single marker association testing and increasing haplotype window size generally correlated with improved statistical strength. Haplotypes located at chromosome 6: 111,936,275-111,964,664 including the rs17754910 marker consistently exceeded the threshold for genome-wide significance with the most significant haplotype (GACGAA; p ≤ 3.34 x 10^−10^) consisting of markers rs671271, rs17754910, rs490080, rs1327199, rs9487771, and rs585057. Composite windows representing all haplotypes of 2-6 SNVs in length were filtered selecting for haplotypes with a χ2 p value ≤ 5 x 10^−5^. This filter was applied as an incremental step towards achieving our objective which was to identify regions where increasing haplotype structure correlated with improved significance. In total 466 haplotypes consisting of 417 unique variants spanning 165 unique haploblocks remained **(Table 1; Supplemental Table S1)**. Of the 165 haploblocks, ten were isolated and nonadjacent. Five of these blocks were excluded from further analysis because: 1) the component SNVs were represented in adjacent clusters (blocks 6611, 7202, 8926, and 9337); or, 2) the isolated block was weakly associated with an existing cluster (block 7489). The remaining isolated haploblocks (blocks 3738, 6951, 7182, 8759, and 9862) failed to exhibit a significant association with age-of-onset based on regression analysis and hence were excluded from further consideration. The remaining 155 haploblocks consisted of 264 unique SNVs and formed consecutive clusters defining 33 discrete chromosomal regions **(Supplemental Table S2)**. The most significant haploblocks within each of the 33 chromosomal regions are defined in **Supplemental Table S3**.

Visualization of haplotype structure was performed using the “Graphical Assessment of Sliding P-values” (GrASP) excel macro (*41*). The GrASP macro concisely depicts haplotype windows of varying length with corresponding p values, providing an efficient means for screening regions of interest while visualizing haplotype substructure. Of the chromosomal regions examined using the GrASP macro, 13 exhibited improvement in statistical significance and incremental changes in haplotype structure with increasing window size **(Table 1)**. Composite images reflecting sliding window p values and the relative position of functional elements within candidate regions are depicted in **Fig 2**. Regions of interest exhibiting improved significance with increasing window size were associated with *TP73, LYPD6B, KIAA1109, ADAD1, IL2*, a regulatory enriched region on chromosome 6, *ARHGEF10, AGO2, CNNM1, LINC00941, PPFIBP1*, and non-coding loci including *AL160035*.*1* and the *NEK4P1* pseudogene. As displayed in **Fig 2** the region defined on chromosome 6 was the only region to exceed the threshold for genome-wide significance. The region defined on chromosome 6 is functionally enriched consisting of predicted regulatory elements including promoter and promoter flanking regions, multiple enhancers, CTCF binding sites, and putative transcription factor binding sites. The nearest sequence element was *LINC02527* located within ∼11 kb of the defined region on chromosome 6. Other regions of particular note identified through this screen included *ARHGEF10* and *IL2* both of which are listed within the COSMIC census of known cancer drivers. Odds ratios and 95% confidence intervals for the aforementioned chromosomal regions are portrayed as a forest plot comparing younger and older breast cancer populations in **Fig 3**. Positive correlations between candidate haplotypes and younger breast cancer patients (diagnosis ≤ 45 years-of-age) relative to older breast cancer patients (diagnosis = 46-50 years-of-age) were associated with *LYPD6B*, the long arm of chromosome 6, *AGO2, LINC00941*, and *PPFIBP1*. The most striking comparison was associated with *LYPD6B* with an OR = 6.95 and a 95% CI of 2.47-19.51. Negative correlations between candidate haplotypes and younger breast cancer patients (diagnosis ≤ 45 years-of-age) relative to older breast cancer patients (diagnosis = 46-50 years-of-age) were associated with *TP73, ADAD1, IL2, ARHGEF10, CNNM1*, the long arm of chromosome 13, and the long arm of chromosome 21.

**Figure 2.**
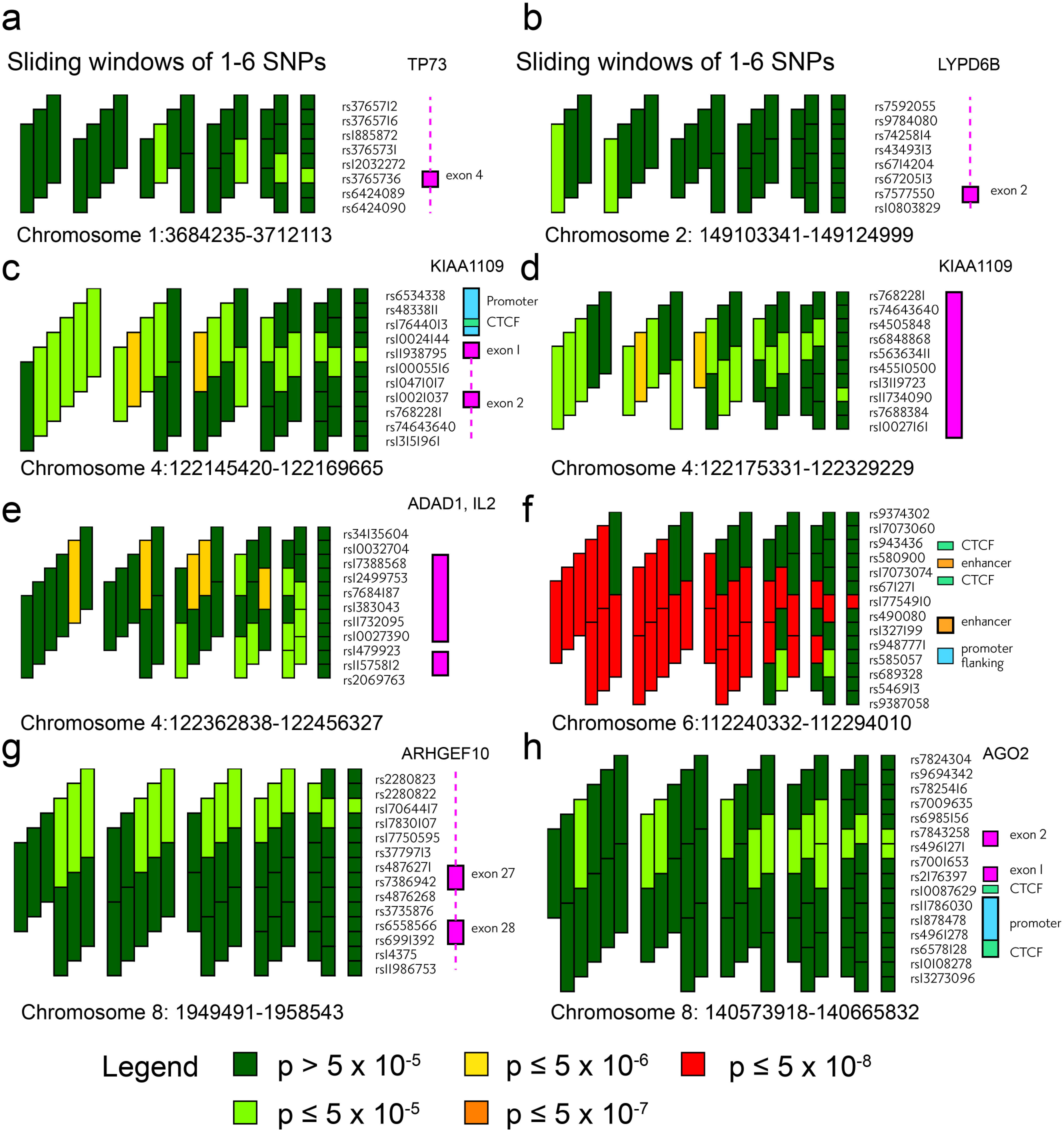
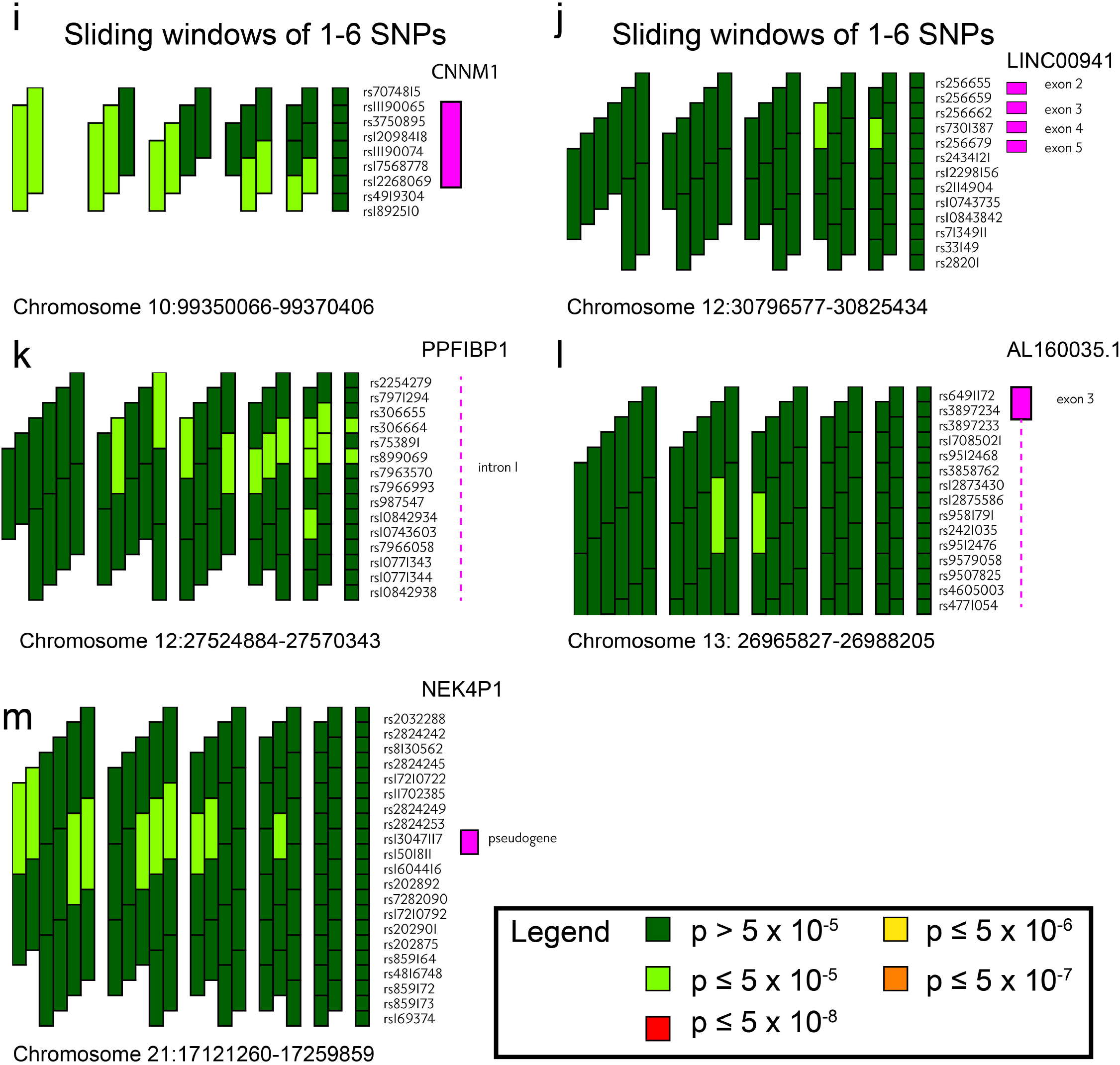
Fine mapping of targeted chromosomal regions. Haplotype analysis was performed using an EM algorithm with a maximum of 50 iterations and sliding windows consisting of 2-6 SNPs. Haplotype windows were aligned and graphically depicted using the GrASP excel macro. Individual haploblocks are color-coded to represent p values (dark green p > 5 x 10^−5^; light green p ≤ 5 x 10^−5^; yellow p ≤ 5 x 10^−6^; orange p ≤ 5 x 10^−7^; red p ≤ 5 x 10^−8^).

**Figure 3.**
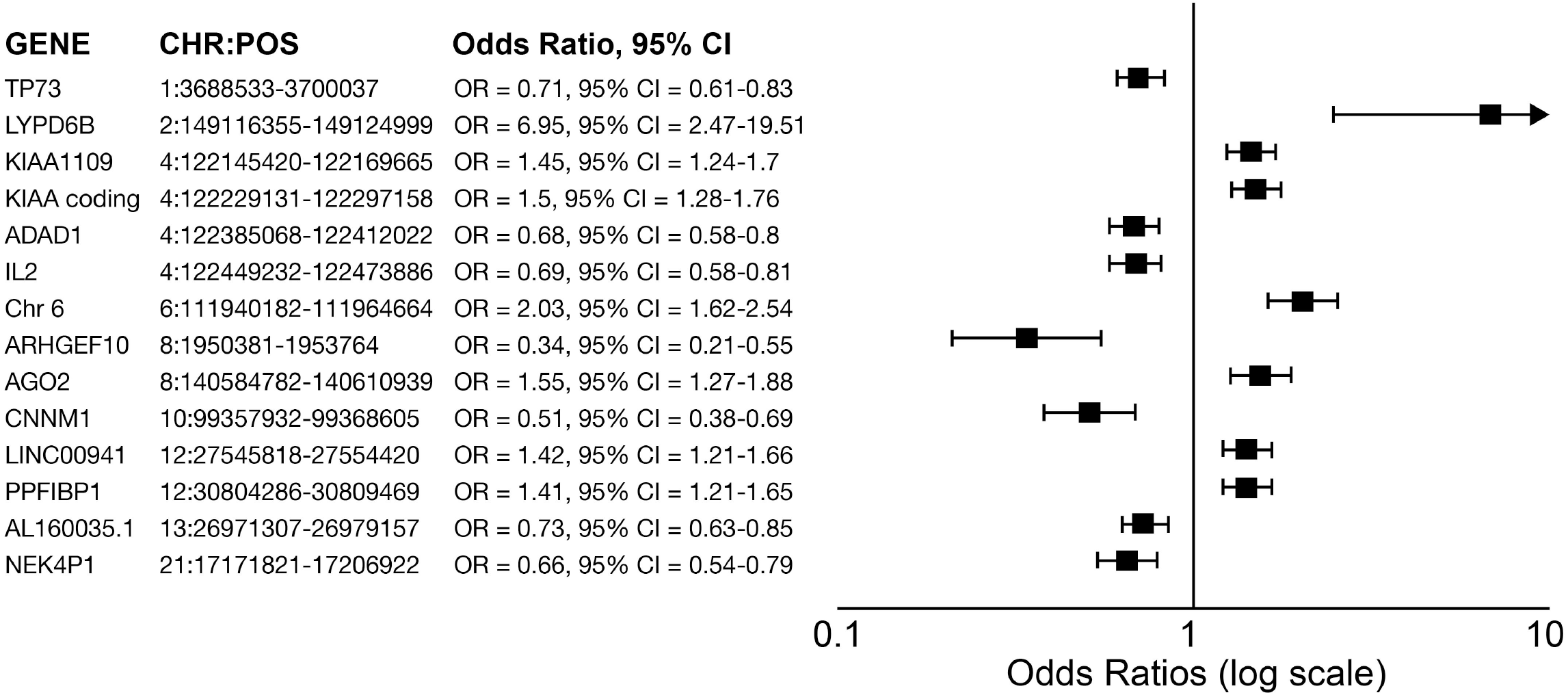
Odds ratios associated with candidate haplotypes. EM frequencies were used to calculate odds ratios and 95% confidence intervals comparing frequencies between younger and older populations within *The Two Sister Study*.

Comparison of haplotype frequencies between siblings, however, failed to fully address the potential for overmatching as previously described (*36*). Correction for hidden population stratification through the use of kinship matrices provides an important and essential control in genotypic analyses but may be more robustly controlled for through population-based haplotype frequency analysis drawing upon data outside of the discovery population. Hence, we evaluated haplotype frequencies observed within *The Two Sister Study* against phase III data from the 1,000 Genomes Project **(Fig 4)**. Towards this end haplotype frequencies derived from The Two Sister Study were compared to haplotype frequencies observed in African (AFR), American (AMR), East Asian (EAS), and non-Finnish European (EUR) populations. Viewed within this context, haplotype frequencies within *The Two Sister Study* were elevated in comparison to AFR and/or AMR populations for *TP73*, the *KIAA1109* promoter, *ADAD1, IL2, ARHGEF10, CNNM1*, and the long arms of chromosomes 6, 13, and 21. Eight haplotypes did not occur within the EAS population. Minimal variation was observed in the non-Finnish EUR population relative to *The Two Sister Study*.

**Figure 4.**
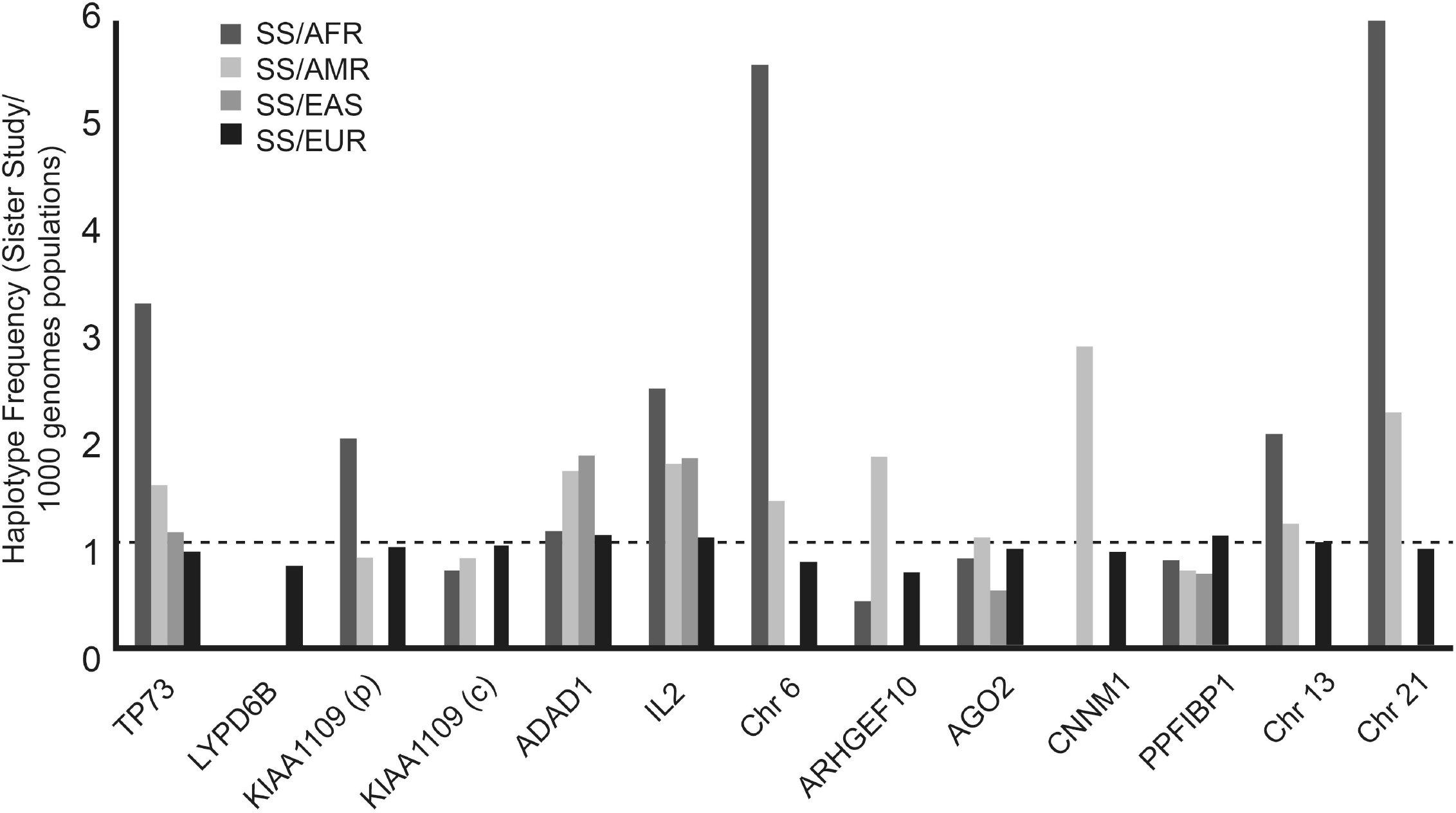
Comparison of haplotype frequencies in The Two Sister Study and phase III of the 1000 Genomes Project. Bar graphs present the ratios formed by dividing *The Two Sister Study* haplotype frequency with population-specific haplotype frequencies obtained through the 1000 Genomes Project. AFR = African, AMR = American, EAS = East Asian, EUR = non-Finnish European.

Haplotype trend regression was used to analyze the aforementioned 33 autosomal regions of interest in both affected (1,456 breast cancer patients) and unaffected (525 discordant sibs) populations as originally defined within *The Two Sister Study*. Whereas visual representation of data using the GrASP macro provided an intuitive sense of evolving haplotype structure, haplotype trend regression provided robust measures of statistical significance. Full model permuted p values indicated 14 of the 33 regions investigated were significantly associated with EOBC within the affected population **(Table 2)**. Conversely, haplotype trend regression failed to detect significant associations between the candidate regions and discordant sibs. Haplomaps summarizing marker distributions are presented in **Supplemental Figure S3** and marker characteristics are described in **Supplemental Table S4**.

**Table 2.** Haplotype trend regression comparing discordant sibs. **Haplotype trend regression underscores significant differences between breast cancer patients and unaffected sibs**. Haplotype trend regression was performed using haplotypes as defined in **Table 1**. By way of contrast, regression analysis was performed using age-at-diagnosis as a quantitative trait comparing breast cancer patients to unaffected sibs. CHR = chromosome, POS = position of the first marker, FM P = the p value resulting from full model trend regression, Bon.P = the Bonferroni adjusted p value, and PermP = the permuted p value after 1,000 permutations. The 14 haplotypes all showed a significant association with age-of-onset in breast cancer patients, whereas no significance was observed in discordant sibs. Although not displayed, similar analysis using age-at-diagnosis as a categorical variable yielded similar findings.

| Candidates |  | Affected |  |  | Unaffected Sibs |  |  |
| --- | --- | --- | --- | --- | --- | --- | --- |
| CHR | POS | FM P | Bon.P | PermP | FM P | Bon.P | PermP |
| 1 | 3605097 | 3.44E-18 | 6.55E-03 | 4.9E-02 | 6.57E-39 | 1 | 1 |
| 2 | 149978783 | 8.19E-21 | 1.21E-05 | 1.0E-03 | 6.85E-40 | 1 | 0.99 |
| 4 | 123066575 | 9.94E-19 | 1.71E-03 | 1.0E-02 | 3.58E-39 | 1 | 1 |
| 4 | 123150286 | 1.18E-20 | 1.53E-05 | 1.0E-03 | 4.47E-40 | 0.36 | 0.86 |
| 4 | 123306223 | 1.34E-19 | 2.01E-04 | 2.0E-03 | 1.45E-39 | 0.11 | 0.59 |
| 4 | 123370387 | 1.82E-19 | 2.77E-04 | 2.0E-03 | 4.98E-41 | 0.051 | 0.45 |
| 6 | 112261385 | 3.65E-18 | 6.47E-03 | 4.7E-02 | 5.79E-39 | 1 | 1 |
| 8 | 1898547 | 2.77E-18 | 5.18E-03 | 3.3E-02 | 5.90E-40 | 0.76 | 0.95 |
| 8 | 141594881 | 7.49E-19 | 1.26E-03 | 5.0E-03 | 6.56E-40 | 0.88 | 0.97 |
| 10 | 101117689 | 2.23E-18 | 3.05E-03 | 1.9E-02 | 1.44E-39 | 1 | 1 |
| 12 | 27698751 | 3.04E-18 | 5.74E-03 | 4.0E-02 | 1.70E-40 | 0.073 | 0.52 |
| 12 | 30957220 | 1.28E-18 | 2.25E-03 | 1.4E-02 | 7.46E-39 | 1 | 1 |
| 13 | 27545444 | 7.29E-19 | 1.23E-03 | 5.0E-03 | 2.91E-39 | 1 | 1 |
| 21 | 18544139 | 2.15E-18 | 3.94E-03 | 2.5E-02 | 2.10E-37 | 1 | 1 |

In an attempt to validate our findings, we first analyzed breast cancer expression data obtained through cbioportal (*43*). The expression data included the “mRNA expression z-scores relative to normal samples (log RNA Seq V2 RSEM)” file and included representing 994 donors. Variance partition analysis was performed to evaluate associations between gene expression and age-at-diagnosis. Summary findings indicated that expression of *AGO2, KIAA1109*, and *PPFIBP1* was significantly associated with breast cancer age-at-diagnosis and explained 4.47% of age-related variance within the population **(Table 3)**.

**Table 3.** Correlating gene expression with age-at-diagnosis. **Expression of AGO2, KIAA1109, and PPFIBP1 correlates with age-at diagnosis in breast cancer patients**. Gene expression data was regressed using the VariancePartition R package. The additive effect of AGO2, KIAA1109, and PPFIBP1 expression contributed to 4.47% of the variance in age across the TCGA breast cancer data set.

| GENE | T | PR(>\|T\|) | F-STAT | P | VARIANCE |
| --- | --- | --- | --- | --- | --- |
| AGO2 | -3.86 | 1.19E-04 | 14.93 | 1.19E-04 | 1.48% |
| ARHGEF10 | -0.89 | 3.72E-01 | 0.7984 | 3.72E-01 | 0.08% |
| CNNM1 | 0.056 | 9.56E-01 | 0.0031 | 9.56E-01 | 0.00% |
| IL2 | 1.32 | 1.88E-01 | 1.733 | 1.88E-01 | 0.17% |
| KIAA1109 | -4.41 | 1.13E-05 | 19.48 | 1.13E-05 | 1.92% |
| LYPD6B | 1.41 | 1.59E-01 | 1.99 | 1.59E-01 | 0.20% |
| PPFIBP1 | -3.28 | 1.07E-03 | 10.77 | 1.07E-03 | 1.07% |
| TP73 | -0.55 | 5.83E-01 | 0.3013 | 5.83E-01 | 0.03% |

Subsequent analysis was performed in an attempt to correlate gene-specific mutation types with age-at-diagnosis. Due to the rarity of discrete mutation types the study population was expanded to include 30 studies across various tissues that were accessed through cbioportal. Pediatric studies were excluded from analysis and 20 years-of-age was applied as a cutoff to exclude minors. Subpopulations were defined by the affected gene and type of mutation (amplification, deletion, missense/truncating mutation). Significant associations linking age-at-diagnosis to (candidate x mutation type) were identified by two sample Z-test **(Table 4)**. The mean age-at-diagnosis ± standard deviation for controls was 60.2 ± 13.13 years-of-age. Significant associations between age-at-diagnosis and gene-specific copy number variants involving amplifications were observed in *ARHGEF10* (63 ± 10.86; p = 0.029); *CNNM1* (54.2 ± 11.61; p = 0.004); *LYPD6B* (57.5 ± 13.39; p = 0.04); and *TP73* (62.9 ± 10.38; p = 0.021). Significant associations between age-at-diagnosis and gene-specific copy number variants involving deletions were observed in *ADAD1* (64.4 ± 10.06; p = 0.0047); *AGO2* (64.6 ± 11.02; p = 0.022); *CNNM1* (66.1 ± 10.42; p = 0.00011); *IL2* (64.4 ± 10.06; p = 0.0047); *KIAA1109* (64.6 ± 9.57; p = 0.0023); and *LYPD6B* (64.8 ± 10.36; p = 0.00091). Significant associations involving missense/truncating mutations followed a trend similar to that observed in association with deletions as might be expected in terms of functional consequence.

**Table 4.** Correlating mutation-type with age-at-diagnosis. **CNNM1 and LYPD6B exhibit bidirectionality of effect on age-at-diagnosis depending upon mutation type**. Two-sample Z testing was applied to compare gene-specific mutation types with the control population. Gene amplifications in CNNM1 and LYPD6B correlated with a lower age-at-diagnosis whereas deletions or truncating mutations correlated with an increased age-at-diagnosis.

| GROUP | AMPLIFICATION |  |  |  |  | DEEP DELETION |  |  |  |  | MUTATION |  |  |  |  |
| --- | --- | --- | --- | --- | --- | --- | --- | --- | --- | --- | --- | --- | --- | --- | --- |
|  | Mean | SD | N | Z | P | Mean | SD | N | Z | P | Mean | SD | N | Z | P |
| CONTROL | 60.2 | 13.13 | 7026 | 0.00 | N/A | 60.2 | 13.13 | 7026 | 0 | N/A | 60.2 | 13.13 | 7026 | 0.00 | N/A |
| ADAD1 | 58.3 | 12.65 | 22 | 1.13 | 0.26 | 64.4 | 10.06 | 26 | -2.83 | 4.70E-03 | 61.5 | 13.27 | 99 | -1.04 | 2.97E-01 |
| AGO2 | 60.3 | 12.53 | 910 | -0.10 | 0.92 | 64.6 | 11.20 | 11 | -2.30 | 2.17E-02 | 64.6 | 13.04 | 131 | -3.83 | 1.28E-04 |
| ARHGEF10 | 63.0 | 10.86 | 54 | -2.20 | 0.029 | 61.9 | 11.46 | 298 | -1.80 | 7.14E-02 | 65.2 | 12.59 | 131 | -4.40 | 1.10E-05 |
| CNNM1 | 54.2 | 11.61 | 8 | 2.90 | 0.004 | 66.1 | 10.42 | 23 | -3.86 | 1.14E-04 | 64.2 | 12.44 | 87 | -3.20 | 1.37E-03 |
| IL2 | 57.6 | 12.54 | 21 | 1.53 | 0.127 | 64.4 | 10.06 | 26 | -2.83 | 4.70E-03 | 60.9 | 13.05 | 23 | -0.39 | 7.00E-01 |
| KIAA1109 | 58.9 | 12.33 | 23 | 0.80 | 0.43 | 64.6 | 9.57 | 27 | -3.05 | 2.33E-03 | 63.4 | 12.74 | 400 | -3.55 | 3.82E-04 |
| LYPD6B | 57.5 | 13.39 | 21 | 2.01 | 0.044 | 64.8 | 10.36 | 35 | -3.32 | 9.16E-04 | 64.1 | 12.53 | 35 | -2.53 | 1.14E-02 |
| PPFIBP1 | 60.2 | 13.20 | 171 | 0.00 | 1 | 59.3 | 12.98 | 7 | 0.42 | 6.71E-01 | 63.2 | 13.74 | 94 | -2.35 | 1.86E-02 |
| TP73 | 62.9 | 10.38 | 42 | -2.30 | 0.021 | 62. | 12.61 | 33 | -1.63 | 1.02E-01 | 62.9 | 13.43 | 89 | -2.09 | 3.70E-02 |

## DISCUSSION

Our objective in this retrospective study was to gain insight into the genetics of EOBC using existing data sets. We proposed to do so through a subtle rephrasing of the initial hypothesis and by applying haplotype-based methods rather than single marker tests of association. *The Two Sister Study* data set was chosen for retrospective analysis because it is among the best characterized studies involving EOBC and because the data structure lends itself to formation of alternative hypotheses. We believe there is a need to explore alternatives in the study of complex disease in general because the greater portion of phenotypic heterogeneity in complex disease remains unexplained. By way of example the investigation of breast cancer has resulted in the identification of a handful of genetic drivers with large effect and more than 200 susceptibility loci with minor effect explaining less than half of breast cancer heritability. Known drivers associated with EOBC are less well defined. Yet EOBC accounts for an estimated 10% of all new breast cancer cases among women and an estimated 15% of breast cancer deaths result from breast cancers initially diagnosed prior to 45 years-of-age (*3, 44*).

*The Two Sister Study* made use of a familial study design to identify maternally-mediated affects and germline associations with EOBC by contrasting breast cancer patients diagnosed prior to the age of 50 against discordant siblings (*25, 26*). In the current study we addressed a different question and hypothesized that candidate associations with EOBC might more readily be identified by contrasting younger and older cases of breast cancer. This supposition is consistent with arguments presented by Kerber and colleagues (*38*), although the merits of treating age as a categorical variable remains a subject of debate (*45-47*).

Initial haplotype-based association studies to compare cases (age-at-diagnosis ≤ 45) and controls (age-at-diagnosis = 46-50) yielded normally distributed results as determined by QQ plot. Preliminary screening alone identified a single SNV exceeding genome-wide significance (rs17754910; p = 4.73 x 10^−9^, FDR = 0.0016). Haplotype-based association testing and sliding window analysis helped identify 33 chromosomal regions of interest, 13 of which exhibited increasing haplotype structure in conjunction with improved measures of significance. The qualitative observations resulting from sliding window analysis were subsequently corroborated by haplotype trend regression with 14 of the 33 candidate regions achieving a permuted p value ≤ 0.05. It should be noted that the only haplotypes to achieve genome-wide significance by means of haplotype-based association testing included the rs17754910 SNV on chromosome 6 in a region enriched with regulatory elements. The nearest sequence element approximately 11 kb upstream of the rs17754910 SNV is the non-coding *LINC02527* RNA (chromosome 6: 111,900,306-111,909,395). We note that alternating methylation patterns are observed within the *LINC02527* promoter in various cancers including breast cancer (*48*). Other candidates identified by haplotype trend regression included the known cancer drivers *IL2* and *ARHGEF10* (interleukin 2 and rho guanine nucleotide exchange factor 10, respectively) neither of which have previously been associated with an early-onset cancer phenotype. The remaining candidates identified by haplotype trend regression may be broadly categorized in terms of known involvement in cancer, metastasis, and age-of-onset in disease. *AGO2* (argonaute 2), *CNNM1* (cyclin and CBS domain divalent metal cation transport mediator 1), *KIAA1109*, and *TP73* (tumor promoter 73) have been implicated in breast cancer, metastasis, and disease age-of-onset (*49-57*). The noncoding *LINC00941* RNA, *LYPD6B* (LY6/PLAUR Domain Containing 6B), and *PPFIBP1* (liprin-beta-1) have been implicated in cancers that may or may not include breast cancer, have been implicated in metastasis, but have no known association with disease age-of-onset (*58-61*). *ADAD1* (adenosine deaminase domain containing 1) has no known association with cancer but has been associated with early-onset asthma and may have a role in childhood seizures. Last, the *NEK4P1* pseudogene and the AL160035.1 sequence have no known associations with cancer, metastasis, or disease onset. Though not addressed within the body of this study, we note that Ingenuity Pathway Analysis associated *AGO2, ARHGEF10, CNNM1, IL2, KIAA1109, LYPD6B, PPFIBP1*, and *TP73* with a single network centered around nodes formed by *TP53* and the estrogen receptor. We note the obvious absence of *BRCA1*/*BRCA2* within the network and mention it here as an anecdote worthy of speculation.

Haplotype frequency analysis within *The Two Sister Study* and phase III data from the 1000 Genomes Project yielded insight specifically with regard to the potential hazards of overmatching in study design. As mentioned overmatching presents a potentially significant challenge in complex disease models where discordant sibs are likely to share an indeterminate number of disease-related alleles (*36*). If, as suggested by the current literature, hundreds of discrete susceptibility loci control breast cancer occurrence and phenotypic expression, we must consider the possibility that familial controls carry a greater burden of polygenic risk alleles without necessarily experiencing disease occurrence. Because no single allele drives breast cancer occurrence, it logically follows that differences in allele or haplotype frequencies between discordant sibs may lack the capacity to distinguish between alleles associated with phenotypic heterogeneity in complex disease. It is known that familial controls may carry a significantly elevated polygenic load relative to unrelated cases or healthy controls creating an uncontrolled source of bias in discovery (*37*). By way of example we note that haplotype frequencies are elevated in discordant sibs relative to affected breast cancer patients for *TP73, IL2*, and *ARHGEF10* **(Fig 3)**. *IL2* and *ARHGEF10* are both listed within the COSMIC census of known cancer drivers and it does not require a stretch of the imagination to consider that *TP73* might play a role in breast cancer. Based solely upon haplotype frequencies observed in discordant sibs, it would appear that all three haplotypes are negatively correlated with EOBC. Yet the observed haplotype frequencies for these three genes within *The Two Sister Study* are elevated when compared to the 1000 genomes phase III AMR population by 1.54-fold, 1.68-fold, and 1.82-fold, respectively. The undefined element on chromosome 21 is elevated by 18.86-fold relative to the AFR population although the very same haplotype is more abundant in discordant sib controls relative to breast cancer patients diagnosed at ≤ 50 years-of-age.

Because this study was retrospective a replication of our findings would be challenging without a prospective collection of new data, something which is beyond the scope of the current study. In the absence of replication, we have attempted to validate our findings with supporting evidence as a matter of due diligence. Towards this goal breast cancer gene expression data derived from the TCGA pan-cancer study was evaluated using regression modeling and variance partition analysis to identify correlations between gene expression and age-at-diagnosis. Earlier age-at-diagnosis was associated with higher expression of *AGO2* (p = 1.19 x 10^−4^), *KIAA1109* (p = 1.13 x 10^−5^), and *PPFIBP1* (p = 1.07 x 10^−3^). These findings are consistent with prior studies involving *AGO2* and *KIAA1109* and provide new evidence suggesting a potential association of *PPFIBP1* expression in EOBC (*50, 54*). Under the assumption of an additive model, expression of these three genes was calculated to explain a combined 4.47% of age-related variance within the study population.

Subsequent validation involved the evaluation of age-at-diagnosis as a function of gene-specific mutations drawing upon available data from 30 distinct cancer studies for statistical purposes. Of the gene-specific mutations the vast majority were observed to result in a significant increase in the mean age-at-diagnosis. We speculate that most of these mutations are unlikely to be causally associated with late-onset disease and instead reflect the global accumulation of damage as a secondary consequence of errors in DNA repair. The candidate genes *CNNM1* and *LYPD6B* shared a unique feature, though, in that both exhibited bidirectionality of effect depending upon mutation status. Gene amplifications affecting *CNNM1* and *LYPD6B* were associated with a significantly lower mean age-at-diagnosis (54.2 ± 11.61 and 57.5 ± 13.39 years-of-age, respectively). Deletions affecting *CNNM1* and *LYPD6B* were conversely associated with a significant increase in mean age-at-diagnosis (66.1 ± 10.42 and 64.8 ± 10.36 years-of-age, respectively). This bidirectionality of effect, we believe, is sufficiently compelling to warrant further investigation of *CNNM1* and *LYPD6B* as contributory factors in EOBC.

Complex disease phenotypes remain a major challenge in the genomic sciences. Frequentist strategies, based upon the assumption that more data will translate into more insight, are currently in vogue and serve a valuable purpose. The identification of rare variants associated with disease is a matter of sample size and ongoing efforts to integrate disparate data sets for meta-analysis is a monumental challenge. Our objective in the current study is less ambitious and merely asks if we can repurpose data to improve our understanding of complex disease. To that limited extent, we have achieved our goal. We have identified a new candidate of genome-wide significance with a potential role in EOBC. We have provided strong supporting evidence justifying the pursuit of a handful of priority candidates with a potential role in EOBC. We have identified two known cancer drivers with a potential involvement in disease onset. And, we have highlighted conditions where frequentist analysis may lead to questionable conclusions in the analysis of familial data. Data-mining, in this instance, suggests that there may be merit in re-examining existing data and the assumptions made during initial inquiry.

## METHODS

### Data: *The Two Sister Study*

Discovery was performed using data derived from *The Two Sister Study: A Family-Based Study of Genes and Environment in Young-Onset Breast Cancer* (accession phs000678.v1.p1). Study contents were accessed under a Data Use Certification (DUC) Agreement via the Database of Genotypes and Phenotypes (dbGAP). The dataset includes genotypic, phenotypic, and demographic data for 1,456 patients, 525 discordant sib controls, and an additional 1,359 controls. The demographics of the population have been described (*25, 26, 39*). The parent study described 1,458 patients. We believe two of these patients were erroneously excluded from the present study during filtering to eliminate duplicate samples. Quality control filtering of the corresponding genotypic data retained a total of 684,126 variants with a call rate ≥ 0.99, a minor allele frequency ≥ 0.05, and a Hardy-Weinberg p value ≥ 1 x 10^−6^ within the older “control” population.

### Data: cbioportal

In order to validate initial findings clinical data spanning 30 studies representing 10,902 donors was accessed through cbioportal (*43*). A total of 220 donors were excluded due to cross-study differences in the definition of donor age. An additional 23 donors diagnosed prior to the age of 20 were excluded as minors. The studies were selected based on three criteria: 1) existing evidence of an early-onset cancer phenotype within the tissue; 2) the availability of data defining age-at-diagnosis; and 3) exclusion of pediatric studies. Composite data included the following studies: Acute Myeloid Leukemia (OHSU) (*62*), Breast Cancer (METABRIC) (*63, 64*), Breast Cancer (SMC) (*65*), Breast Fibroepithelial Tumors (Duke-NUS) (*66*), Breast Invasive Carcinoma (British Columbia) (*67*), Breast Invasive Carcinoma (Broad) (*68*), Breast Invasive Carcinoma (Sanger) (*69*), Breast Invasive Carcinoma (TCGA, PanCancer Atlas) (*70*), Cervical Squamous Cell Carcinoma (TCGA, PanCancer Atlas) (*71*), Clear Cell Renal Cell Carcinoma (DFCI) (*72*), Colorectal Adenocarcinoma (DFCI) (*73*), Colorectal Adenocarcinoma (TCGA, PanCancer Atlas) (*71*), Esophageal Adenocarcinoma (TCGA, PanCancer Atlas) (*71*), Esophageal Squamous Cell Carcinoma (ICGC) (*74*), Esophageal Squamous Cell Carcinoma (UCLA) (*75*), Kidney Chromophobe (TCGA, PanCancer Atlas) (*71*), Kidney Renal Clear Cell Carcinoma (BGI) (*76*), Kidney Renal Clear Cell Carcinoma (IRC) (*77*), Kidney Renal Clear Cell Carcinoma (TCGA, PanCancer Atlas), Kidney Renal Papillary Cell Carcinoma (TCGA, PanCancer Atlas) (*71*), Liver Hepatocellular Carcinoma (TCGA, PanCancer Atlas) (*71*), Lung Adenocarcinoma (OncoSG) (*78*), Lung Adenocarcinoma (TCGA, PanCancer Atlas) (*71*), Ovarian Serous Cystadenocarcinoma (TCGA, PanCancer Atlas) (*71*), Prostate Adenocarcinoma (Broad/Cornell) (*79*), Prostate Adenocarcinoma (Fred Hutchinson CRC) (*80*), Prostate Adenocarcinoma (TCGA, PanCancer Atlas) (*71*), Small Cell Carcinoma of the Ovary (MSKCC) (*81*), Uterine Carcinosarcoma (Johns Hopkins) (*82*), and Uterine Corpus Endometrial Carcinoma (TCGA, PanCancer Atlas) (*71*). Samples lacking mutation in any of the candidate genes were assembled into a control dataset (N = 7026). In order to validate initial findings, experimental data sets were assembled on a per gene basis and subcategorized according to mutation type (amplification, deep deletion, or missense/truncating mutations).

### Data: The 1000 Genomes Project

Genotypic data from phase III of the 1000 Genomes Project was obtained through the Ensembl data portal Donor identification numbers were matched to genotypic data in order to assemble haplotypes. Quantification of haplotype frequencies was subsequently performed using the Haploview software package.

### Cutpoint optimization

The %findcut SAS macro was used to calculate cutpoints as previously described (*83*). The mean value was used as the cutoff for dichotomizing the case population in all subsequent analyses.

### Association testing

All association testing was performed using the Sequence Variation Suite software package (Golden Helix, Bozeman, MO) and a custom workstation with dual Xeon Gold 12-core processors and 192 Gb RAM (Thinkmate, Waltham, MA). Genotypic data sourced from *The Two Sister Study* was filtered to exclude variants with a call rate ≤ 0.99, a minor allele frequency < 0.05, or extremes in Hardy-Weinberg disequilibrium within the control population (p ≤ 1 x 10^−6^). In order to identify regions of interest, haplotype-based association testing was performed using an EM algorithm (50-iterations, convergence tolerance = 0.0001, frequency threshold = 0.01) and a dynamic window size of 10 kilobases (kb). Covariates in the analysis included race, family history of disease occurrence, and age-at-menopause. Regions of interest were identified using a threshold of p ≤ 5 x 10^−4^.

### Sliding window haplotype-based association testing

Sliding window analysis was performed essentially as described by Mathias *et al* (*41*). Genotypic data from patients affected by breast cancer was filtered as described leaving a total of 684,126 variants for analysis. Sliding window analysis was subsequently performed using windows of varying size (2-6 SNPs) to evaluate unphased haplotypes. Analysis was performed using a case-control design and an EM algorithm (50-iterations, convergence tolerance = 0.0001, haplotype frequency threshold = 0.01). Applied test statistics consisted of a single test per sliding window and not the individual tests for each haplotype. For comparison, a single-locus Efficient Mixed-Model Association eXpedited (EMMAX) analysis was performed under an additive model using Sequence Variation Suite software (Golden Helix).

### Manhattan plots

Manhattan plots were generated by plotting of observed versus expected -log[p] values. A single plot was constructed using output from haplotype-based association testing with a static window size of 10 kb for reference. A second composite plot was constructed by overlaying the output from single marker association tests and sliding window association tests using windows of 2, 4, and 6 SNVs in length.

### Graphical assessment of p-values from sliding window haplotype tests (GrASP)

GrASP is a graphical tool for displaying p-values from sliding window tests (*41*). The Excel add-on produces a simple graphic that simultaneously depicts the width of the sliding windows while using user-specified color to specify varying levels of significance. GrASP allows the user to identify regions/blocks of interest, based jointly on the absolute p-value of the tests from these windows and the building of haplotypes of significance in the region. Graphical representations for regions of interest were assembled and trimmed to display regions of increasing significance while minimizing the length of flanking sequence falling below a threshold of p < 5 x 10^−5^. Assembled images were presented within the context of functional genomic elements as defined within the Ensembl human genome browser (GRCh38). GrASP is freely available for use at: http://research.nhgri.nih.gov/GrASP/.

### Forest plot

Odds ratios (OR) and 95% confidence intervals (95% CI) were derived from a single overall test per sliding window, and not the individual tests of deviation for each haplotype. Weighting was performed using –(log[p]) as the weighting variable so that symbol size directly correlated with significance. A Forest plot depicting the OR and 95% CI was generated using the “DistillerSR Forest Plot Generator from Evidence Partners” web resource (https://www.evidencepartners.com/resources/forest-plot-generator/).

### Haplotype trend regression

Haplotype trend regression was performed using the Sequence Variation Suite software package (Golden Helix). Analysis was performed using predefined blocks as described within the text and **Supplemental Table S3**. Stepwise regression was performed using backwards elimination and up to 50 EM iterations with a convergence tolerance of 0.0001 and frequency threshold of 0.01. Full versus reduced model regression was performed using age-at-diagnosis as a quantitative trait, with race, family history of disease, and menopause status as covariates. Correction for multiple testing was performed using Bonferroni adjusted p values and 1,000 full scan permutations.

### Haplotype frequency analysis

EM frequencies representing the 1,456 cases defined in *The Two Sister Study* were contrasted against population-specific haplotype frequencies. Population-specific data was gathered from phase III of the 1000 Genomes Project and haplotype frequencies were determined using Haploview software (*84*).

### Variance partition analysis

Data derived from the TCGA Pancancer Atlas Breast Invasive Carcinoma study (*70*) was evaluated in the R software environment using the VariancePartition application (*85*). Expression data consisted of z-score measures relative to normal samples obtained through cbioportal. Expression data was unavailable for *ADAD1* and hence this candidate was excluded from the analysis. Age-at-diagnosis was correlated with expression data as previously described.

### Pancancer mutation analysis

To evaluate the impact of mutation type on cancer age-of-onset meta-data representing 9 candidate genes from 30 studies was obtained as described. A total of 220 donors were excluded due to cross-study differences in the definition of donor age. Pertinent data included age-at-diagnosis/diagnosis age, gene-specific copy number variants, gene-specific coding variants (missense/truncating). Samples lacking mutation in any of the candidate genes were assembled into a control dataset (N = 7026). Experimental populations were defined on a per gene basis and were classified by mutation type (amplifications, deep deletions, or missense/truncating mutations). Distribution analysis was performed using a two sample Z-test and the equation 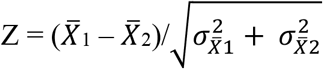 where 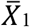 is the mean age-at-diagnosis for the control population,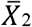 is the mean age-at-diagnosis for the case population, 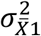 is the standard deviation for the control population divided by the square root of the number of data points, and 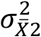 is the standard deviation for the case population divided by the square root of the number of data points. Corresponding p values were calculated for each independent test statistic.

## Data Availability

Pertinent data may be accessed through dbGAP. The Two Sister Study: A Family-Based Study of Genes and Environment in Young-Onset Breast Cancer (accession phs000678.v1.p1). All other data sources including but not limited to cbioportal, 1000 Genomes, and The Cancer Genome Atlas are within the public domain and may be freely accoessed using public data portals.

https://www.ncbi.nlm.nih.gov/projects/gap/cgi-bin/study.cgi?study_id=phs000678.v1.p1

## ACKNOWLEDGMENTS

We acknowledge the Two Sister Study PI, Clarice R. Weinberg, Susan G. Komen for the Cure (grant FAS703856) and the Intramural Program of the National Institute of Environmental Health Sciences. Fei, Cl, DeRoo, L., Sandler, D.P. and Weinberg, C.R. Fertility drugs and young**-**onset breast cancer: Results from the Two Sister Study. Journal of the National Cancer Institute 104(13): 1021-7, 2012 (PMID 22773825).(*86*) Without the invaluable work of the aforementioned investigators, the present analysis would not be possible.

## BIBLIOGRAPHY

1. K. Michailidou et al., Genome-wide association analysis of more than 120,000 individuals identifies 15 new susceptibility loci for breast cancer. Nature genetics 47, 373–380 (2015).

2. F. Bray et al., Global cancer statistics 2018: GLOBOCAN estimates of incidence and mortality worldwide for 36 cancers in 185 countries. CA Cancer J Clin 68, 394–424 (2018).

3. D. Chelmow et al., Executive Summary of the Early-Onset Breast Cancer Evidence Review Conference. Obstetrics & Gynecology 135, 1457–1478 (2020).

4. H. A. Assi et al., Epidemiology and prognosis of breast cancer in young women. Journal of Thoracic Disease, S2-S8 (2013).

5. K. E. Malone et al., Prevalence and predictors of BRCA1 and BRCA2 mutations in a population-based study of breast cancer in white and black American women ages 35 to 64 years. Cancer research 66, 8297–8308 (2006).

6. M. M. Gaudet et al., Identification of a BRCA2-specific modifier locus at 6p24 related to breast cancer risk. PLoS genetics 9, e1003173 (2013).

7. C. K. Anders et al., Young Age at Diagnosis Correlates With Worse Prognosis and Defines a Subset of Breast Cancers With Shared Patterns of Gene Expression. Journal of Clinical Oncology 26, 3324–3330 (2008).

8. R. W. Morris, N. L. Kaplan, On the advantage of haplotype analysis in the presence of multiple disease susceptibility alleles. Genetic epidemiology 23, 221–233 (2002).

9. J. Akey, L. Jin, M. Xiong, Haplotypes vs single marker linkage disequilibrium tests: what do we gain? European Journal of Human Genetics 9, 291–300 (2001).

10. Y. He et al., Accelerating haplotype-based genome-wide association study using perfect phylogeny and phase-known reference data. PloS one 6, e22097 (2011).

11. The International HapMap Project. Nature 426, 789–796 (2003).

12. Y. Wang, J. Lu, J. Yu, R. A. Gibbs, F. Yu, An integrative variant analysis pipeline for accurate genotype/haplotype inference in population NGS data. Genome research 23, 833–842 (2013).

13. D. M. Howard et al., Genome-wide haplotype-based association analysis of major depressive disorder in Generation Scotland and UK Biobank. Translational psychiatry 7, 1263 (2017).

14. M. Shi et al., Genome wide study of maternal and parent-of-origin effects on the etiology of orofacial clefts. American journal of medical genetics. Part A 158a, 784-794 (2012).

15. P. Khankhanian, P. A. Gourraud, A. Lizee, D. S. Goodin, Haplotype-based approach to known MS-associated regions increases the amount of explained risk. Journal of medical genetics 52, 587–594 (2015).

16. D. M. Howard et al., Genome-wide haplotype-based association analysis of major depressive disorder in Generation Scotland and UK Biobank. Translational psychiatry 7, 1263 (2017).

17. L. H. Pereira et al., The BRCA1 Ashkenazi founder mutations occur on common haplotypes and are not highly correlated with anonymous single nucleotide polymorphisms likely to be used in genome-wide case-control association studies. BMC Genet 8, 68 (2007).

18. Q. Wang et al., Genome-wide haplotype association study identifies BLM as a risk gene for prostate cancer in Chinese population. Tumour Biol 36, 2703–2707 (2015).

19. M. G. Kibriya et al., A pilot genome-wide association study of early-onset breast cancer. Breast cancer research and treatment 114, 463–477 (2009).

20. J. S. Barnholtz-Sloan et al., FGFR2 and other loci identified in genome-wide association studies are associated with breast cancer in African-American and younger women. Carcinogenesis 31, 1417–1423 (2010).

21. L. Jara et al., Genetic variants in FGFR2 and MAP3K1 are associated with the risk of familial and early-onset breast cancer in a South-American population. Breast cancer research and treatment 137, 559–569 (2013).

22. K. L. Huang et al., A common haplotype lowers PU.1 expression in myeloid cells and delays onset of Alzheimer’s disease. Nature neuroscience 20, 1052–1061 (2017).

23. K. S. Wang et al., Genetic association analysis of ITGB3 polymorphisms with age at onset of schizophrenia. J Mol Neurosci 51, 446–453 (2013).

24. in Global registry and database on craniofacial anomalies. (World Health Organization).

25. K.M. O’Brien et al., A family-based, genome-wide association study of young-onset breast cancer: inherited variants and maternally mediated effects. European journal of human genetics : EJHG 24, 1316–1323 (2016).

26. M. Shi et al., Previous GWAS hits in relation to young-onset breast cancer. Breast cancer research and treatment 161, 333–344 (2017).

27. G. S. Dite et al., Increased cancer risks for relatives of very early-onset breast cancer cases with and without BRCA1 and BRCA2 mutations. British Journal of Cancer 103, 1103–1108 (2010).

28. C. K. Anders, R. Johnson, J. Litton, M. Phillips, A. Bleyer, Breast cancer before age 40 years. Semin Oncol 36, 237–249 (2009).

29. M. V. Diaz-Santana et al., Perinatal and postnatal exposures and risk of young-onset breast cancer. Breast Cancer Research 22, 88 (2020).

30. I. Sepahi et al., Investigating the effects of additional truncating variants in DNA-repair genes on breast cancer risk in BRCA1-positive women. BMC Cancer 19, 787 (2019).

31. C. K. Anders et al., Young age at diagnosis correlates with worse prognosis and defines a subset of breast cancers with shared patterns of gene expression. J Clin Oncol 26, 3324–3330 (2008).

32. K. Michailidou et al., Genome-wide association analysis of more than 120,000 individuals identifies 15 new susceptibility loci for breast cancer. Nature genetics 47, 373–380 (2015).

33. K. Michailidou et al., Association analysis identifies 65 new breast cancer risk loci. Nature 551, 92–94 (2017).

34. L. Wu et al., A transcriptome-wide association study of 229,000 women identifies new candidate susceptibility genes for breast cancer. Nature genetics 50, 968–978 (2018).

35. W. J. Peyrot, D. I. Boomsma, B. W. Penninx, N. R. Wray, Disease and Polygenic Architecture: Avoid Trio Design and Appropriately Account for Unscreened Control Subjects for Common Disease. American journal of human genetics 98, 382–391 (2016).

36. M. Boehnke, C. D. Langefeld, Genetic association mapping based on discordant sib pairs: the discordant-alleles test. American journal of human genetics 62, 950–961 (1998).

37. M. H. Law et al., Multiplex melanoma families are enriched for polygenic risk. Human molecular genetics 29, 2976–2985 (2020).

38. R. A. Kerber, C. I. Amos, B. Y. Yeap, D. M. Finkelstein, D. C. Thomas, Design considerations in a sib-pair study of linkage for susceptibility loci in cancer. BMC Med Genet 9, 64 (2008).

39. M. Shi, K. M. O’Brien, C. R. Weinberg, Interactions between a Polygenic Risk Score and Non-genetic Risk Factors in Young-Onset Breast Cancer. Sci Rep 10, 3242 (2020).

40. J. Meyers, J. Mandrekar, in Proc SAS Glob Forum. (2015), vol. 3249.

41. R. A. Mathias et al., A graphical assessment of p-values from sliding window haplotype tests of association to identify asthma susceptibility loci on chromosome 11q. BMC Genet 7, 38 (2006).

42. C. Song et al., A genome-wide scan for breast cancer risk haplotypes among African American women. PloS one 8, e57298–e57298 (2013).

43. J. Gao et al., Integrative analysis of complex cancer genomics and clinical profiles using the cBioPortal. Sci Signal 6, pl1 (2013).

44. K. C. Oeffinger et al., Breast Cancer Screening for Women at Average Risk: 2015 Guideline Update From the American Cancer Society. Jama 314, 1599–1614 (2015).

45. D. G. Altman, P. Royston, The cost of dichotomising continuous variables. Bmj 332, 1080 (2006).

46. R. C. MacCallum, S. Zhang, K. J. Preacher, D. D. Rucker, On the practice of dichotomization of quantitative variables. Psychol Methods 7, 19–40 (2002).

47. O. Naggara et al., Analysis by Categorizing or Dichotomizing Continuous Variables Is Inadvisable: An Example from the Natural History of Unruptured Aneurysms. American Journal of Neuroradiology 32, 437–440 (2011).

48. L. Ma et al., LncBook: a curated knowledgebase of human long non-coding RNAs. Nucleic acids research 47, D128–d134 (2019).

49. T. Bellissimo et al., Argonaute 2 drives miR-145-5p-dependent gene expression program in breast cancer cells. Cell Death & Disease 10, 17 (2019).

50. M. C. Casey et al., Quantifying Argonaute 2 (Ago2) expression to stratify breast cancer. BMC Cancer 19, 712 (2019).

51. F. Chen, Y. Zhang, S. Varambally, C. J. Creighton, Molecular Correlates of Metastasis by Systematic Pan-Cancer Analysis Across The Cancer Genome Atlas. Mol Cancer Res 17, 476–487 (2019).

52. U. Chandran et al., Expression of Cnnm1 and Its Association with Stemness, Cell Cycle, and Differentiation in Spermatogenic Cells in Mouse Testis. Biol Reprod 95, 7 (2016).

53. Z. Qiao et al., Mutations in KIAA1109, CACNA1C, BSN, AKAP13, CELSR2, and HELZ2 Are Associated With the Prognosis in Endometrial Cancer. Frontiers in genetics 10, 909–909 (2019).

54. M. T. Kuo et al., Association of fragile site-associated (FSA) gene expression with epithelial differentiation and tumor development. Biochem Biophys Res Commun 340, 887–893 (2006).

55. M. Dutertre et al., Exon-based clustering of murine breast tumor transcriptomes reveals alternative exons whose expression is associated with metastasis. Cancer research 70, 896–905 (2010).

56. J. Yao et al., TP73-AS1 promotes breast cancer cell proliferation through miR-200a-mediated TFAM inhibition. J Cell Biochem 119, 680–690 (2018).

57. J. Zhang et al., FDXR regulates TP73 tumor suppressor via IRP2 to modulate aging and tumor suppression. The Journal of Pathology 251, 284–296 (2020).

58. H. Liu et al., Long Non-coding RNA LINC00941 as a Potential Biomarker Promotes the Proliferation and Metastasis of Gastric Cancer. Frontiers in genetics 10, 5 (2019).

59. Y. Shoji, G. Ch, ramouli, J. Risinger, Over-Expression of Ly6/Plaur Domain Containing 6b (Lypd6b) in Ovarian Cancer. Gynecology & Obstetrics 1, 1–10 (2011).

60. Serra-Pagh, Liprins, a Family of LAR Transmembrane Protein-tyrosine Phosphatase-interacting Proteins*.

61. M. Kriajevska et al., Liprin beta 1, a member of the family of LAR transmembrane tyrosine phosphatase-interacting proteins, is a new target for the metastasis-associated protein S100A4 (Mts1). J Biol Chem 277, 5229–5235 (2002).

62. J. W. Tyner et al., Functional genomic landscape of acute myeloid leukaemia. Nature 562, 526–531 (2018).

63. C. Curtis et al., The genomic and transcriptomic architecture of 2,000 breast tumours reveals novel subgroups. Nature 486, 346–352 (2012).

64. B. Pereira et al., The somatic mutation profiles of 2,433 breast cancers refine their genomic and transcriptomic landscapes. Nature communications 7, 11479 (2016).

65. Z. Kan et al., Multi-omics profiling of younger Asian breast cancers reveals distinctive molecular signatures. Nature communications 9, 1725 (2018).

66. J. Tan et al., Genomic landscapes of breast fibroepithelial tumors. Nature genetics 47, 1341–1345 (2015).

67. S. P. Shah et al., The clonal and mutational evolution spectrum of primary triple-negative breast cancers. Nature 486, 395–399 (2012).

68. S. Banerji et al., Sequence analysis of mutations and translocations across breast cancer subtypes. Nature 486, 405–409 (2012).

69. P. J. Stephens et al., The landscape of cancer genes and mutational processes in breast cancer. Nature 486, 400–404 (2012).

70. G. Ciriello et al., Comprehensive Molecular Portraits of Invasive Lobular Breast Cancer. Cell 163, 506–519 (2015).

71. K. A. Hoadley et al., Cell-of-Origin Patterns Dominate the Molecular Classification of 10,000 Tumors from 33 Types of Cancer. Cell 173, 291-304.e296 (2018).

72. D. Miao et al., Genomic correlates of response to immune checkpoint therapies in clear cell renal cell carcinoma. Science 359, 801–806 (2018).

73. M. Giannakis et al., Genomic Correlates of Immune-Cell Infiltrates in Colorectal Carcinoma. Cell Rep 15, 857–865 (2016).

74. Y. Song et al., Identification of genomic alterations in oesophageal squamous cell cancer. Nature 509, 91–95 (2014).

75. D. C. Lin et al., Genomic and molecular characterization of esophageal squamous cell carcinoma. Nature genetics 46, 467–473 (2014).

76. G. Guo et al., Frequent mutations of genes encoding ubiquitin-mediated proteolysis pathway components in clear cell renal cell carcinoma. Nature genetics 44, 17–19 (2011).

77. M. Gerlinger et al., Genomic architecture and evolution of clear cell renal cell carcinomas defined by multiregion sequencing. Nature genetics 46, 225–233 (2014).

78. J. Chen et al., Genomic landscape of lung adenocarcinoma in East Asians. Nature genetics 52, 177–186 (2020).

79. S. C. Baca et al., Punctuated evolution of prostate cancer genomes. Cell 153, 666–677 (2013).

80. A. Kumar et al., Substantial interindividual and limited intraindividual genomic diversity among tumors from men with metastatic prostate cancer. Nature medicine 22, 369–378 (2016).

81. P. Jelinic et al., Recurrent SMARCA4 mutations in small cell carcinoma of the ovary. Nature genetics 46, 424–426 (2014).

82. S. Jones et al., Genomic analyses of gynaecologic carcinosarcomas reveal frequent mutations in chromatin remodelling genes. Nature communications 5, 5006 (2014).

83. J. Meyers, J. Mandrekar. (2015).

84. J. C. Barrett, B. Fry, J. Maller, M. J. Daly, Haploview: analysis and visualization of LD and haplotype maps. Bioinformatics 21, 263-265 (2005).

85. G. E. Hoffman, E. E. Schadt, variancePartition: interpreting drivers of variation in complex gene expression studies. BMC bioinformatics 17, 483 (2016).

86. C. Fei, L. A. Deroo, D. P. Sandler, C. R. Weinberg, Fertility drugs and young-onset breast cancer: results from the Two Sister Study. Journal of the National Cancer Institute 104, 1021–1027 (2012).

