## Supplemental Table S1 for "Retrospective analysis of *The Two Sister Study* using haplotype-based association testing to identify loci associated with early-onset breast cancer"

**Supplemental Table S1. Filtered haplotypes and corresponding markers.**

| First Marker | CHR | POS | Block | Markers Used | Haplotype | EM Freq | Case | Control | P |
|---|---|---|---|---|---|---|---|---|---|
| rs1885872 | 1 | 3688533 | 57 | rs1885872, rs3765731, rs12032272, rs3765736 | AGGA | 3.42E-01 | 3.03E-01 | 3.82E-01 | 1.36E-05 |
| rs3765731 | 1 | 3695328 | 58 | rs3765731, rs12032272, rs3765736 | GGA | 3.46E-01 | 3.09E-01 | 3.85E-01 | 2.39E-05 |
| rs12032272 | 1 | 3698592 | 59 | rs12032272, rs3765736 | GA | 3.51E-01 | 3.15E-01 | 3.88E-01 | 2.92E-05 |
| rs12144282 | 1 | 62236069 | 162 | rs12144282, rs6688986 | AA | 6.63E-01 | 6.99E-01 | 6.27E-01 | 3.16E-05 |
| rs12144282 | 1 | 62236069 | 162 | rs12144282, rs6688986, rs1050334 | AAA | 6.63E-01 | 6.99E-01 | 6.27E-01 | 3.16E-05 |
| rs6688986 | 1 | 62237048 | 163 | rs6688986, rs1050334 | AA | 6.64E-01 | 7.00E-01 | 6.27E-01 | 2.99E-05 |
| rs2781130 | 1 | 94764958 | 340 | rs2781130, rs7525706, rs3860336, rs17414905, rs2391423, rs6541369 | GAAGGA | 1.17E-02 | 1.97E-02 | 3.47E-03 | 4.83E-05 |
| rs7525706 | 1 | 94770363 | 341 | rs7525706, rs3860336, rs17414905, rs2391423, rs6541369 | AAGGA | 1.20E-02 | 2.04E-02 | 3.47E-03 | 2.74E-05 |
| rs7525706 | 1 | 94770363 | 341 | rs7525706, rs3860336, rs17414905, rs2391423, rs6541369, rs4847214 | AAGGAG | 1.20E-02 | 2.04E-02 | 3.47E-03 | 2.74E-05 |
| rs3860336 | 1 | 94770726 | 342 | rs3860336, rs17414905, rs2391423, rs6541369 | AGGA | 1.20E-02 | 2.04E-02 | 3.47E-03 | 2.74E-05 |
| rs3860336 | 1 | 94770726 | 342 | rs3860336, rs17414905, rs2391423, rs6541369, rs4847214 | AGGAG | 1.20E-02 | 2.04E-02 | 3.47E-03 | 2.74E-05 |
| rs3860336 | 1 | 94770726 | 342 | rs3860336, rs17414905, rs2391423, rs6541369, rs4847214, rs12743267 | AGGAGG | 1.20E-02 | 2.04E-02 | 3.46E-03 | 2.81E-05 |
| rs7425814 | 2 | 149116355 | 574 | rs7425814, rs4349313, rs6714204, rs6720513, rs7577550, rs10803829 | AAGCGG | 1.13E-02 | 1.95E-02 | 2.87E-03 | 1.97E-05 |
| rs4349313 | 2 | 149118982 | 575 | rs4349313, rs6714204, rs6720513, rs7577550, rs10803829 | AGCGG | 1.21E-02 | 2.05E-02 | 3.62E-03 | 3.10E-05 |
| rs4349313 | 2 | 149118982 | 575 | rs4349313, rs6714204, rs6720513, rs7577550, rs10803829, rs12621827 | AGCGGG | 1.17E-02 | 2.04E-02 | 2.92E-03 | 1.17E-05 |
| rs6714204 | 2 | 149122269 | 576 | rs6714204, rs6720513, rs7577550, rs10803829, rs12621827 | GCGGG | 1.18E-02 | 2.01E-02 | 3.37E-03 | 2.88E-05 |
| rs6714204 | 2 | 149122269 | 576 | rs6714204, rs6720513, rs7577550, rs10803829, rs12621827, rs1026889 | GCGGGA | 1.16E-02 | 2.03E-02 | 2.78E-03 | 1.02E-05 |
| rs6720513 | 2 | 149122777 | 577 | rs6720513, rs7577550, rs10803829, rs12621827, rs1026889 | CGGGA | 1.15E-02 | 1.96E-02 | 3.15E-03 | 2.84E-05 |
| rs6720513 | 2 | 149122777 | 577 | rs6720513, rs7577550, rs10803829, rs12621827, rs1026889, rs17432490 | CGGGAG | 1.13E-02 | 1.98E-02 | 2.68E-03 | 1.14E-05 |

| | | | | | | | | | |
|---|---|---|---|---|---|---|---|---|---|
| rs16828069 | 2 | 150332543 | 622 | rs16828069, rs10177059, rs10180142, rs983231, rs12994436 | GGCAA | 1.23E-01 | 9.76E-02 | 1.49E-01 | 2.24E-05 |
| rs16828069 | 2 | 150332543 | 622 | rs16828069, rs10177059, rs10180142, rs983231, rs12994436, rs6760903 | GGCAAA | 1.23E-01 | 9.77E-02 | 1.49E-01 | 2.50E-05 |
| rs10177059 | 2 | 150344535 | 623 | rs10177059, rs10180142, rs983231, rs12994436 | GCAA | 1.24E-01 | 9.80E-02 | 1.50E-01 | 2.15E-05 |
| rs10177059 | 2 | 150344535 | 623 | rs10177059, rs10180142, rs983231, rs12994436, rs6760903 | GCAAA | 1.24E-01 | 9.80E-02 | 1.50E-01 | 2.41E-05 |
| rs10177059 | 2 | 150344535 | 623 | rs10177059, rs10180142, rs983231, rs12994436, rs6760903, rs7600018 | GCAAAA | 1.24E-01 | 9.80E-02 | 1.50E-01 | 2.41E-05 |
| rs10180142 | 2 | 150345320 | 624 | rs10180142, rs983231, rs12994436 | CAA | 1.23E-01 | 9.78E-02 | 1.50E-01 | 2.16E-05 |
| rs10180142 | 2 | 150345320 | 624 | rs10180142, rs983231, rs12994436, rs6760903 | CAAA | 1.23E-01 | 9.79E-02 | 1.50E-01 | 2.42E-05 |
| rs10180142 | 2 | 150345320 | 624 | rs10180142, rs983231, rs12994436, rs6760903, rs7600018 | CAAAA | 1.23E-01 | 9.79E-02 | 1.50E-01 | 2.50E-05 |
| rs10180142 | 2 | 150345320 | 624 | rs10180142, rs983231, rs12994436, rs6760903, rs7600018, rs10930090 | CAAAAG | 1.24E-01 | 9.79E-02 | 1.50E-01 | 2.41E-05 |
| rs983231 | 2 | 150349292 | 625 | rs983231, rs12994436 | AA | 1.23E-01 | 9.78E-02 | 1.49E-01 | 2.23E-05 |
| rs983231 | 2 | 150349292 | 625 | rs983231, rs12994436, rs6760903 | AAA | 1.23E-01 | 9.78E-02 | 1.50E-01 | 2.49E-05 |
| rs983231 | 2 | 150349292 | 625 | rs983231, rs12994436, rs6760903, rs7600018 | AGGA | 7.56E-01 | 7.88E-01 | 7.23E-01 | 2.31E-05 |
| rs983231 | 2 | 150349292 | 625 | rs983231, rs12994436, rs6760903, rs7600018 | AAAA | 1.23E-01 | 9.77E-02 | 1.49E-01 | 2.77E-05 |
| rs983231 | 2 | 150349292 | 625 | rs983231, rs12994436, rs6760903, rs7600018, rs10930090 | AAAAG | 1.24E-01 | 9.79E-02 | 1.50E-01 | 2.41E-05 |
| rs983231 | 2 | 150349292 | 625 | rs983231, rs12994436, rs6760903, rs7600018, rs10930090, rs17434648 | AAAAGA | 1.24E-01 | 9.80E-02 | 1.50E-01 | 2.41E-05 |
| rs12994436 | 2 | 150354384 | 626 | rs12994436, rs6760903 | AA | 1.24E-01 | 9.80E-02 | 1.50E-01 | 2.41E-05 |
| rs12994436 | 2 | 150354384 | 626 | rs12994436, rs6760903 | GG | 8.71E-01 | 8.95E-01 | 8.46E-01 | 2.41E-05 |
| rs12994436 | 2 | 150354384 | 626 | rs12994436, rs6760903, rs7600018 | GGA | 8.23E-01 | 8.52E-01 | 7.94E-01 | 2.48E-05 |
| rs12994436 | 2 | 150354384 | 626 | rs12994436, rs6760903, rs7600018 | AAA | 1.23E-01 | 9.75E-02 | 1.48E-01 | 3.16E-05 |
| rs12994436 | 2 | 150354384 | 626 | rs12994436, rs6760903, rs7600018, rs10930090 | AAAG | 1.24E-01 | 9.80E-02 | 1.50E-01 | 2.41E-05 |
| rs12994436 | 2 | 150354384 | 626 | rs12994436, rs6760903, rs7600018, rs10930090, rs17434648 | AAAGA | 1.24E-01 | 9.80E-02 | 1.50E-01 | 2.41E-05 |
| rs12994436 | 2 | 150354384 | 626 | rs12994436, rs6760903, rs7600018, rs10930090, rs17434648, rs1866979 | AAAGAG | 1.23E-01 | 9.72E-02 | 1.50E-01 | 1.88E-05 |

| | | | | | | | | | |
|---|---|---|---|---|---|---|---|---|---|
| rs6760903 | 2 | 150361519 | 627 | rs6760903, rs7600018, rs10930090, rs17434648, rs1866979, rs7590701 | AAGAGA | 1.22E-01 | 9.59E-02 | 1.48E-01 | 2.10E-05 |
| rs7600018 | 2 | 150362262 | 628 | rs7600018, rs10930090, rs17434648, rs1866979, rs7590701 | AGAGA | 1.22E-01 | 9.76E-02 | 1.48E-01 | 3.74E-05 |
| rs7600018 | 2 | 150362262 | 628 | rs7600018, rs10930090, rs17434648, rs1866979, rs7590701, rs4287765 | AGAGAC | 1.23E-01 | 9.76E-02 | 1.48E-01 | 4.12E-05 |
| rs10930090 | 2 | 150362575 | 629 | rs10930090, rs17434648, rs1866979 | GAG | 1.83E-01 | 1.54E-01 | 2.13E-01 | 3.88E-05 |
| rs10930090 | 2 | 150362575 | 629 | rs10930090, rs17434648, rs1866979, rs7590701 | GAGA | 1.66E-01 | 1.37E-01 | 1.96E-01 | 1.83E-05 |
| rs10930090 | 2 | 150362575 | 629 | rs10930090, rs17434648, rs1866979, rs7590701, rs4287765 | GAGAC | 1.67E-01 | 1.37E-01 | 1.97E-01 | 2.11E-05 |
| rs10930090 | 2 | 150362575 | 629 | rs10930090, rs17434648, rs1866979, rs7590701, rs4287765, rs10187222 | GAGACA | 1.67E-01 | 1.37E-01 | 1.97E-01 | 2.31E-05 |
| rs17434648 | 2 | 150367344 | 630 | rs17434648, rs1866979, rs7590701 | AGA | 1.75E-01 | 1.45E-01 | 2.04E-01 | 2.91E-05 |
| rs17434648 | 2 | 150367344 | 630 | rs17434648, rs1866979, rs7590701, rs4287765 | AGAC | 1.75E-01 | 1.46E-01 | 2.05E-01 | 2.99E-05 |
| rs17434648 | 2 | 150367344 | 630 | rs17434648, rs1866979, rs7590701, rs4287765, rs10187222 | AGACA | 1.75E-01 | 1.46E-01 | 2.05E-01 | 2.99E-05 |
| rs17434648 | 2 | 150367344 | 630 | rs17434648, rs1866979, rs7590701, rs4287765, rs10187222, rs11684751 | AGACAA | 1.75E-01 | 1.46E-01 | 2.05E-01 | 3.30E-05 |
| rs1866979 | 2 | 150370850 | 631 | rs1866979, rs7590701 | GA | 1.75E-01 | 1.46E-01 | 2.05E-01 | 3.48E-05 |
| rs1866979 | 2 | 150370850 | 631 | rs1866979, rs7590701, rs4287765 | GAC | 1.75E-01 | 1.46E-01 | 2.05E-01 | 3.61E-05 |
| rs1866979 | 2 | 150370850 | 631 | rs1866979, rs7590701, rs4287765, rs10187222 | GACA | 1.75E-01 | 1.46E-01 | 2.05E-01 | 3.61E-05 |
| rs1866979 | 2 | 150370850 | 631 | rs1866979, rs7590701, rs4287765, rs10187222, rs11684751 | GACAA | 1.75E-01 | 1.46E-01 | 2.05E-01 | 3.94E-05 |
| rs1866979 | 2 | 150370850 | 631 | rs1866979, rs7590701, rs4287765, rs10187222, rs11684751, rs976396 | GACAAA | 1.75E-01 | 1.46E-01 | 2.05E-01 | 3.94E-05 |
| rs7590701 | 2 | 150381362 | 632 | rs7590701, rs4287765 | AC | 1.75E-01 | 1.46E-01 | 2.05E-01 | 3.48E-05 |
| rs7590701 | 2 | 150381362 | 632 | rs7590701, rs4287765, rs10187222 | ACA | 1.75E-01 | 1.46E-01 | 2.05E-01 | 3.48E-05 |
| rs7590701 | 2 | 150381362 | 632 | rs7590701, rs4287765, rs10187222, rs11684751 | ACAA | 1.75E-01 | 1.46E-01 | 2.05E-01 | 3.48E-05 |
| rs7590701 | 2 | 150381362 | 632 | rs7590701, rs4287765, rs10187222, rs11684751, rs976396 | ACAAA | 1.75E-01 | 1.46E-01 | 2.05E-01 | 3.48E-05 |
| rs16824126 | 2 | 227889633 | 695 | rs16824126, exm272588, rs7566347, rs16824173, rs2946931, rs11680252 | AAGGCA | 3.71E-01 | 4.06E-01 | 3.36E-01 | 3.73E-05 |
| rs1715828 | 2 | 227893838 | 696 | exm272588, rs7566347, rs16824173, rs2946931, rs11680252 | AGGCA | 3.74E-01 | 4.10E-01 | 3.36E-01 | 3.63E-05 |

| | | | | | | | | | |
|---|---|---|---|---|---|---|---|---|---|
| rs1715828 | 2 | 227893838 | 696 | exm272588, rs7566347, rs16824173, rs2946931, rs11680252, exm-rs10191097 | AGGCAA | 3.75E-01 | 4.12E-01 | 3.38E-01 | 4.15E-05 |
| rs7566347 | 2 | 227894838 | 697 | rs7566347, rs16824173, rs2946931, rs11680252 | GGCA | 3.74E-01 | 4.10E-01 | 3.37E-01 | 3.85E-05 |
| rs7566347 | 2 | 227894838 | 697 | rs7566347, rs16824173, rs2946931, rs11680252, exm-rs10191097 | GGCAA | 3.74E-01 | 4.11E-01 | 3.37E-01 | 3.20E-05 |
| rs7566347 | 2 | 227894838 | 697 | rs7566347, rs16824173, rs2946931, rs11680252, exm-rs10191097, rs3748863 | GGCAAG | 3.73E-01 | 4.09E-01 | 3.36E-01 | 4.45E-05 |
| rs6534338 | 4 | 122105714 | 1535 | rs6534338, rs4833811, rs17644013, rs10024144, rs11938795, rs10005516 | GAAGGG | 2.61E-01 | 2.29E-01 | 2.94E-01 | 4.34E-05 |
| rs4833811 | 4 | 122106568 | 1536 | rs4833811, rs17644013, rs10024144, rs11938795 | AAGG | 2.66E-01 | 2.32E-01 | 3.00E-01 | 2.80E-05 |
| rs4833811 | 4 | 122106568 | 1536 | rs4833811, rs17644013, rs10024144, rs11938795, rs10005516 | AAGGG | 2.66E-01 | 2.32E-01 | 3.00E-01 | 2.10E-05 |
| rs4833811 | 4 | 122106568 | 1536 | rs4833811, rs17644013, rs10024144, rs11938795, rs10005516, rs10471017 | AAGGGA | 2.66E-01 | 2.32E-01 | 3.01E-01 | 2.56E-05 |
| rs17644013 | 4 | 122128482 | 1537 | rs17644013, rs10024144, rs11938795 | AGG | 2.67E-01 | 2.32E-01 | 3.02E-01 | 2.14E-05 |
| rs17644013 | 4 | 122128482 | 1537 | rs17644013, rs10024144, rs11938795, rs10005516 | AGGG | 2.66E-01 | 2.32E-01 | 3.01E-01 | 1.46E-05 |
| rs17644013 | 4 | 122128482 | 1537 | rs17644013, rs10024144, rs11938795, rs10005516, rs10471017 | AGGGA | 2.67E-01 | 2.33E-01 | 3.02E-01 | 1.80E-05 |
| rs17644013 | 4 | 122128482 | 1537 | rs17644013, rs10024144, rs11938795, rs10005516, rs10471017, rs10021037 | AGGGAG | 2.67E-01 | 2.33E-01 | 3.02E-01 | 1.80E-05 |
| rs10024144 | 4 | 122145420 | 1538 | rs10024144, rs11938795 | GA | 6.87E-01 | 7.26E-01 | 6.48E-01 | 6.65E-06 |
| rs10024144 | 4 | 122145420 | 1538 | rs10024144, rs11938795 | GG | 2.68E-01 | 2.33E-01 | 3.02E-01 | 2.55E-05 |
| rs10024144 | 4 | 122145420 | 1538 | rs10024144, rs11938795, rs10005516 | GAG | 6.85E-01 | 7.24E-01 | 6.44E-01 | 5.62E-06 |
| rs10024144 | 4 | 122145420 | 1538 | rs10024144, rs11938795, rs10005516 | GGG | 2.67E-01 | 2.33E-01 | 3.02E-01 | 2.16E-05 |
| rs10024144 | 4 | 122145420 | 1538 | rs10024144, rs11938795, rs10005516, rs10471017 | GAGA | 6.82E-01 | 7.20E-01 | 6.43E-01 | 4.46E-06 |
| rs10024144 | 4 | 122145420 | 1538 | rs10024144, rs11938795, rs10005516, rs10471017 | GGGA | 2.67E-01 | 2.33E-01 | 3.02E-01 | 2.97E-05 |
| rs10024144 | 4 | 122145420 | 1538 | rs10024144, rs11938795, rs10005516, rs10471017, rs10021037 | GAGAG | 6.82E-01 | 7.20E-01 | 6.43E-01 | 4.46E-06 |
| rs10024144 | 4 | 122145420 | 1538 | rs10024144, rs11938795, rs10005516, rs10471017, rs10021037 | GGGAG | 2.67E-01 | 2.33E-01 | 3.02E-01 | 2.96E-05 |

| | | | | | | | | | |
|---|---|---|---|---|---|---|---|---|---|
| rs10024144 | 4 | 122145420 | 1538 | rs10024144, rs11938795, rs10005516, rs10471017, rs10021037, rs7682281 | GGGAGA | 2.68E-01 | 2.34E-01 | 3.03E-01 | 2.93E-05 |
| rs11938795 | 4 | 122151854 | 1539 | rs11938795, rs10005516 | AG | 6.99E-01 | 7.37E-01 | 6.60E-01 | 5.62E-06 |
| rs11938795 | 4 | 122151854 | 1539 | rs11938795, rs10005516 | GG | 2.68E-01 | 2.34E-01 | 3.04E-01 | 2.19E-05 |
| rs11938795 | 4 | 122151854 | 1539 | rs11938795, rs10005516, rs10471017 | AGA | 6.83E-01 | 7.21E-01 | 6.43E-01 | 5.51E-06 |
| rs11938795 | 4 | 122151854 | 1539 | rs11938795, rs10005516, rs10471017 | GGA | 2.68E-01 | 2.34E-01 | 3.03E-01 | 2.24E-05 |
| rs11938795 | 4 | 122151854 | 1539 | rs11938795, rs10005516, rs10471017, rs10021037 | AGAG | 6.83E-01 | 7.21E-01 | 6.43E-01 | 5.52E-06 |
| rs11938795 | 4 | 122151854 | 1539 | rs11938795, rs10005516, rs10471017, rs10021037 | GGAG | 2.68E-01 | 2.34E-01 | 3.03E-01 | 2.24E-05 |
| rs11938795 | 4 | 122151854 | 1539 | rs11938795, rs10005516, rs10471017, rs10021037, rs7682281 | GGAGA | 2.68E-01 | 2.34E-01 | 3.04E-01 | 2.19E-05 |
| rs11938795 | 4 | 122151854 | 1539 | rs11938795, rs10005516, rs10471017, rs10021037, rs7682281, rs74643640 | GGAGAA | 2.69E-01 | 2.34E-01 | 3.04E-01 | 2.28E-05 |
| rs6848868 | 4 | 122229131 | 1549 | rs6848868, rs6848868, rs56363411, rs45510500, rs13119723 | AATGA | 6.98E-01 | 7.40E-01 | 6.55E-01 | 5.73E-07 |
| rs6848868 | 4 | 122229131 | 1549 | rs6848868, rs6848868, rs56363411, rs45510500, rs13119723, rs11734090 | AATGAA | 6.60E-01 | 6.98E-01 | 6.20E-01 | 8.48E-06 |
| rs6848868 | 4 | 122229131 | 1550 | rs6848868, rs56363411, rs45510500, rs13119723 | ATGA | 6.98E-01 | 7.40E-01 | 6.55E-01 | 5.74E-07 |
| rs6848868 | 4 | 122229131 | 1550 | rs6848868, rs56363411, rs45510500, rs13119723, rs11734090 | ATGAA | 6.60E-01 | 6.98E-01 | 6.20E-01 | 8.47E-06 |
| rs56363411 | 4 | 122250504 | 1551 | rs56363411, rs45510500, rs13119723, rs11734090 | TGAA | 6.60E-01 | 6.98E-01 | 6.20E-01 | 8.50E-06 |
| rs45510500 | 4 | 122258745 | 1552 | rs45510500, rs13119723, rs11734090 | GAA | 6.89E-01 | 7.27E-01 | 6.50E-01 | 6.54E-06 |
| rs13119723 | 4 | 122297158 | 1553 | rs13119723, rs11734090 | AA | 7.30E-01 | 7.65E-01 | 6.93E-01 | 1.28E-05 |
| rs11734090 | 4 | 122306958 | 1554 | rs11734090, rs7688384 | GG | 2.70E-01 | 2.35E-01 | 3.07E-01 | 1.28E-05 |
| rs11734090 | 4 | 122306958 | 1554 | rs11734090, rs7688384, rs10027161 | GGA | 2.70E-01 | 2.35E-01 | 3.07E-01 | 1.28E-05 |
| rs11734090 | 4 | 122306958 | 1554 | rs11734090, rs7688384, rs10027161, rs2306369 | GGAA | 2.70E-01 | 2.34E-01 | 3.06E-01 | 1.11E-05 |
| rs11734090 | 4 | 122306958 | 1554 | rs11734090, rs7688384, rs10027161, rs2306369, rs2306369 | GGAAA | 2.70E-01 | 2.34E-01 | 3.06E-01 | 1.11E-05 |
| rs11734090 | 4 | 122306958 | 1554 | rs11734090, rs7688384, rs10027161, rs2306369, rs2306369, rs1127348 | GGAAAA | 2.69E-01 | 2.34E-01 | 3.06E-01 | 1.26E-05 |
| rs10032704 | 4 | 122385068 | 1562 | rs10032704, rs17388568, rs17388568, rs12499753 | AGGA | 2.96E-01 | 2.57E-01 | 3.35E-01 | 3.44E-06 |

| | | | | | | | | | |
|---|---|---|---|---|---|---|---|---|---|
| rs10032704 | 4 | 122385068 | 1562 | rs10032704, rs17388568, rs17388568, rs12499753, rs7684187 | AGGAG | 2.91E-01 | 2.53E-01 | 3.29E-01 | 4.71E-06 |
| rs10032704 | 4 | 122385068 | 1562 | rs10032704, rs17388568, rs17388568, rs12499753, rs7684187, rs1383043 | AGGAGG | 2.91E-01 | 2.53E-01 | 3.29E-01 | 4.69E-06 |
| rs17388568 | 4 | 122408207 | 1563 | rs17388568, rs17388568, rs12499753, rs7684187 | GGAG | 2.91E-01 | 2.53E-01 | 3.29E-01 | 5.42E-06 |
| rs17388568 | 4 | 122408207 | 1563 | rs17388568, rs17388568, rs12499753, rs7684187, rs1383043 | GGAGG | 2.91E-01 | 2.53E-01 | 3.29E-01 | 4.74E-06 |
| rs17388568 | 4 | 122408207 | 1564 | rs17388568, rs12499753, rs7684187 | GAG | 2.91E-01 | 2.53E-01 | 3.29E-01 | 5.42E-06 |
| rs17388568 | 4 | 122408207 | 1564 | rs17388568, rs12499753, rs7684187, rs1383043 | GAGG | 2.91E-01 | 2.53E-01 | 3.29E-01 | 4.74E-06 |
| rs12499753 | 4 | 122412022 | 1565 | rs12499753, rs7684187 | AG | 2.91E-01 | 2.53E-01 | 3.29E-01 | 5.75E-06 |
| rs12499753 | 4 | 122412022 | 1565 | rs12499753, rs7684187, rs1383043 | AGG | 2.91E-01 | 2.53E-01 | 3.29E-01 | 4.74E-06 |
| rs7684187 | 4 | 122420004 | 1566 | rs7684187, rs1383043 | GG | 2.91E-01 | 2.53E-01 | 3.29E-01 | 5.75E-06 |
| rs11732095 | 4 | 122427190 | 1568 | rs11732095, rs10027390 | AA | 7.10E-01 | 7.48E-01 | 6.72E-01 | 6.86E-06 |
| rs10027390 | 4 | 122447361 | 1569 | rs10027390, rs1479923 | GG | 2.90E-01 | 2.52E-01 | 3.28E-01 | 6.86E-06 |
| rs10027390 | 4 | 122447361 | 1569 | rs10027390, rs1479923, rs11575812 | GGG | 2.87E-01 | 2.50E-01 | 3.26E-01 | 5.66E-06 |
| rs10027390 | 4 | 122447361 | 1569 | rs10027390, rs1479923, rs11575812, rs2069763 | GGGC | 2.87E-01 | 2.50E-01 | 3.26E-01 | 5.67E-06 |
| rs10027390 | 4 | 122447361 | 1569 | rs10027390, rs1479923, rs11575812, rs2069763, rs10857092 | GGGCG | 2.87E-01 | 2.50E-01 | 3.26E-01 | 5.35E-06 |
| rs10027390 | 4 | 122447361 | 1569 | rs10027390, rs1479923, rs11575812, rs2069763, rs10857092, rs6848139 | GGGCGA | 2.87E-01 | 2.49E-01 | 3.26E-01 | 4.61E-06 |
| rs57119445 | 4 | 122449232 | 1570 | rs1479923, rs11575812 | GG | 2.87E-01 | 2.50E-01 | 3.26E-01 | 5.43E-06 |
| rs57119445 | 4 | 122449232 | 1570 | rs1479923, rs11575812, rs2069763 | GGC | 2.87E-01 | 2.50E-01 | 3.26E-01 | 5.43E-06 |
| rs57119445 | 4 | 122449232 | 1570 | rs1479923, rs11575812, rs2069763, rs10857092 | GGCG | 2.87E-01 | 2.50E-01 | 3.26E-01 | 5.13E-06 |
| rs57119445 | 4 | 122449232 | 1570 | rs1479923, rs11575812, rs2069763, rs10857092, rs6848139 | GGCGA | 2.87E-01 | 2.49E-01 | 3.26E-01 | 4.44E-06 |
| rs11575812 | 4 | 122449894 | 1571 | rs11575812, rs2069763 | GC | 2.87E-01 | 2.50E-01 | 3.26E-01 | 5.43E-06 |
| rs11575812 | 4 | 122449894 | 1571 | rs11575812, rs2069763, rs10857092 | GCG | 2.87E-01 | 2.50E-01 | 3.26E-01 | 5.13E-06 |
| rs11575812 | 4 | 122449894 | 1571 | rs11575812, rs2069763, rs10857092, rs6848139 | GCGA | 2.87E-01 | 2.49E-01 | 3.26E-01 | 4.45E-06 |
| rs11738463 | 5 | 126344047 | 2047 | rs11738463, rs7730928, rs10068159, rs10057227, rs12513492 | GAAAG | 8.98E-02 | 1.12E-01 | 6.76E-02 | 3.09E-05 |

| | | | | | | | | | |
|---|---|---|---|---|---|---|---|---|---|
| rs11738463 | 5 | 126344047 | 2047 | rs11738463, rs7730928, rs10068159, rs10057227, rs12513492, rs10478724 | GAAAGG | 8.98E-02 | 1.12E-01 | 6.75E-02 | 2.96E-05 |
| rs7730928 | 5 | 126345196 | 2048 | rs7730928, rs10068159, rs10057227, rs12513492 | AAAG | 8.98E-02 | 1.12E-01 | 6.77E-02 | 3.18E-05 |
| rs7730928 | 5 | 126345196 | 2048 | rs7730928, rs10068159, rs10057227, rs12513492, rs10478724 | AAAGG | 8.98E-02 | 1.12E-01 | 6.75E-02 | 2.27E-05 |
| rs7730928 | 5 | 126345196 | 2048 | rs7730928, rs10068159, rs10057227, rs12513492, rs10478724, rs7727523 | AAAGGG | 8.99E-02 | 1.12E-01 | 6.75E-02 | 2.32E-05 |
| rs10068159 | 5 | 126348266 | 2049 | rs10068159, rs10057227, rs12513492 | AAG | 9.07E-02 | 1.13E-01 | 6.84E-02 | 3.04E-05 |
| rs10068159 | 5 | 126348266 | 2049 | rs10068159, rs10057227, rs12513492, rs10478724 | AAGG | 9.06E-02 | 1.12E-01 | 6.83E-02 | 3.20E-05 |
| rs10068159 | 5 | 126348266 | 2049 | rs10068159, rs10057227, rs12513492, rs10478724, rs7727523 | AAGGG | 9.06E-02 | 1.12E-01 | 6.86E-02 | 4.01E-05 |
| rs10057227 | 5 | 126349682 | 2050 | rs10057227, rs12513492 | AG | 9.12E-02 | 1.14E-01 | 6.85E-02 | 2.51E-05 |
| rs10057227 | 5 | 126349682 | 2050 | rs10057227, rs12513492, rs10478724 | AGG | 9.04E-02 | 1.12E-01 | 6.82E-02 | 3.14E-05 |
| rs10057227 | 5 | 126349682 | 2050 | rs10057227, rs12513492, rs10478724, rs7727523 | AGGG | 9.08E-02 | 1.13E-01 | 6.85E-02 | 3.33E-05 |
| rs10057227 | 5 | 126349682 | 2050 | rs10057227, rs12513492, rs10478724, rs7727523, rs1549189 | AGGGA | 9.03E-02 | 1.12E-01 | 6.85E-02 | 4.99E-05 |
| rs12513492 | 5 | 126350206 | 2051 | rs12513492, rs10478724 | GG | 9.12E-02 | 1.14E-01 | 6.86E-02 | 2.50E-05 |
| rs12513492 | 5 | 126350206 | 2051 | rs12513492, rs10478724, rs7727523 | GGG | 9.13E-02 | 1.14E-01 | 6.87E-02 | 2.56E-05 |
| rs12513492 | 5 | 126350206 | 2051 | rs12513492, rs10478724, rs7727523, rs1549189 | GGGA | 9.09E-02 | 1.13E-01 | 6.85E-02 | 3.08E-05 |
| rs12513492 | 5 | 126350206 | 2051 | rs12513492, rs10478724, rs7727523, rs1549189, rs10519907 | GGGAG | 9.06E-02 | 1.12E-01 | 6.86E-02 | 3.89E-05 |
| rs12513492 | 5 | 126350206 | 2051 | rs12513492, rs10478724, rs7727523, rs1549189, rs10519907, rs10519908 | GGGAGA | 9.06E-02 | 1.12E-01 | 6.86E-02 | 3.81E-05 |
| rs10478724 | 5 | 126350217 | 2052 | rs10478724, rs7727523, rs1549189, rs10519907, rs10519908, rs7733479 | GGAGAA | 9.06E-02 | 1.14E-01 | 6.65E-02 | 8.15E-06 |
| rs7727523 | 5 | 126350689 | 2053 | rs7727523, rs1549189, rs10519907, rs10519908, rs7733479 | GAGAA | 9.02E-02 | 1.13E-01 | 6.64E-02 | 8.92E-06 |
| rs10519907 | 5 | 126357690 | 2055 | rs10519907, rs10519908, rs7733479, rs7700582 | GAAC | 6.68E-02 | 8.62E-02 | 4.70E-02 | 2.29E-05 |
| rs10519907 | 5 | 126357690 | 2055 | rs10519907, rs10519908, rs7733479, rs7700582, rs17595984 | GAACG | 6.68E-02 | 8.64E-02 | 4.69E-02 | 2.04E-05 |

| | | | | | | | | | |
|---|---|---|---|---|---|---|---|---|---|
| rs10519907 | 5 | 126357690 | 2055 | rs10519907, rs10519908, rs7733479, rs7700582, rs17595984, rs10519910 | GAACGG | 6.50E-02 | 8.48E-02 | 4.49E-02 | 1.78E-05 |
| rs10519908 | 5 | 126358504 | 2056 | rs10519908, rs7733479, rs7700582 | AAC | 6.72E-02 | 8.64E-02 | 4.77E-02 | 3.05E-05 |
| rs10519908 | 5 | 126358504 | 2056 | rs10519908, rs7733479, rs7700582, rs17595984 | AACG | 6.74E-02 | 8.68E-02 | 4.77E-02 | 2.73E-05 |
| rs10519908 | 5 | 126358504 | 2056 | rs10519908, rs7733479, rs7700582, rs17595984, rs10519910 | AACGG | 6.56E-02 | 8.50E-02 | 4.59E-02 | 2.57E-05 |
| rs7733479 | 5 | 126360998 | 2057 | rs7733479, rs7700582 | AC | 6.80E-02 | 8.78E-02 | 4.79E-02 | 1.90E-05 |
| rs7733479 | 5 | 126360998 | 2057 | rs7733479, rs7700582, rs17595984 | ACG | 6.78E-02 | 8.76E-02 | 4.78E-02 | 1.89E-05 |
| rs7733479 | 5 | 126360998 | 2057 | rs7733479, rs7700582, rs17595984, rs10519910 | ACGG | 6.58E-02 | 8.53E-02 | 4.60E-02 | 1.90E-05 |
| rs1545751 | 5 | 178553791 | 2326 | rs1545751, rs262012, rs34594012, rs262037 | ACGG | 3.68E-02 | 5.10E-02 | 2.23E-02 | 3.46E-05 |
| rs1545751 | 5 | 178553791 | 2326 | rs1545751, rs262012, rs34594012, rs262037, rs185493 | ACGGA | 3.67E-02 | 5.12E-02 | 2.19E-02 | 2.50E-05 |
| rs1545751 | 5 | 178553791 | 2326 | rs1545751, rs262012, rs34594012, rs262037, rs185493, rs625820 | ACGGAC | 3.52E-02 | 5.04E-02 | 1.97E-02 | 6.61E-06 |
| rs262012 | 5 | 178555012 | 2327 | rs262012, rs34594012, rs262037, rs185493, rs625820 | CGGAC | 5.95E-02 | 7.86E-02 | 4.00E-02 | 1.13E-05 |
| rs262012 | 5 | 178555012 | 2327 | rs262012, rs34594012, rs262037, rs185493, rs625820, rs262066 | CGGACG | 4.27E-02 | 5.94E-02 | 2.57E-02 | 6.35E-06 |
| rs34594012 | 5 | 178560739 | 2328 | rs34594012, rs262037, rs185493, rs625820 | GGAC | 6.08E-02 | 7.97E-02 | 4.15E-02 | 1.73E-05 |
| rs34594012 | 5 | 178560739 | 2328 | rs34594012, rs262037, rs185493, rs625820, rs262066 | GGACG | 4.30E-02 | 5.94E-02 | 2.62E-02 | 9.25E-06 |
| rs262037 | 5 | 178563885 | 2329 | rs262037, rs185493, rs625820 | GAC | 8.05E-02 | 1.03E-01 | 5.78E-02 | 8.33E-06 |
| rs262037 | 5 | 178563885 | 2329 | rs262037, rs185493, rs625820, rs262066 | GACG | 6.33E-02 | 8.33E-02 | 4.29E-02 | 6.71E-06 |
| rs11963612 | 6 | 111775142 | 2607 | rs11963612, rs9384805, rs2148709 | GAG | 3.25E-01 | 3.60E-01 | 2.89E-01 | 2.82E-05 |
| rs11963612 | 6 | 111775142 | 2607 | rs11963612, rs9384805, rs2148709, rs2148709 | GAGG | 3.25E-01 | 3.60E-01 | 2.89E-01 | 2.82E-05 |
| rs11963612 | 6 | 111775142 | 2607 | rs11963612, rs9384805, rs2148709, rs2148709, rs7761230 | GAGGG | 3.25E-01 | 3.60E-01 | 2.88E-01 | 3.04E-05 |
| rs11963612 | 6 | 111775142 | 2607 | rs11963612, rs9384805, rs2148709, rs2148709, rs7761230, rs17072912 | GAGGGG | 3.24E-01 | 3.60E-01 | 2.88E-01 | 3.59E-05 |
| rs9384805 | 6 | 111780522 | 2608 | rs9384805, rs2148709 | AG | 3.28E-01 | 3.63E-01 | 2.92E-01 | 3.13E-05 |
| rs9384805 | 6 | 111780522 | 2608 | rs9384805, rs2148709, rs2148709 | AGG | 3.28E-01 | 3.63E-01 | 2.92E-01 | 3.13E-05 |
| rs9384805 | 6 | 111780522 | 2608 | rs9384805, rs2148709, rs2148709, rs7761230 | AGGG | 3.28E-01 | 3.63E-01 | 2.92E-01 | 3.15E-05 |

| | | | | | | | | | |
|---|---|---|---|---|---|---|---|---|---|
| rs9384805 | 6 | 111780522 | 2608 | rs9384805, rs2148709, rs2148709, rs7761230, rs17072912 | AGGGG | 3.28E-01 | 3.63E-01 | 2.92E-01 | 2.93E-05 |
| rs9384805 | 6 | 111780522 | 2608 | rs9384805, rs2148709, rs2148709, rs7761230, rs17072912, rs1022650 | AGGGGC | 3.28E-01 | 3.63E-01 | 2.92E-01 | 2.96E-05 |
| rs17073060 | 6 | 111920678 | 2632 | rs17073060, rs943436, rs580900, rs17073074, rs671271, rs17754910 | AAAGGA | 1.37E-01 | 1.76E-01 | 9.79E-02 | 6.97E-10 |
| rs943436 | 6 | 111920951 | 2633 | rs943436, rs580900, rs17073074, rs671271, rs17754910 | AAGGA | 1.38E-01 | 1.77E-01 | 9.82E-02 | 6.20E-10 |
| rs943436 | 6 | 111920951 | 2633 | rs943436, rs580900, rs17073074, rs671271, rs17754910, rs490080 | AAGGAC | 1.38E-01 | 1.77E-01 | 9.80E-02 | 7.82E-10 |
| rs580900 | 6 | 111936275 | 2634 | rs580900, rs17073074, rs671271, rs17754910 | AGGA | 1.38E-01 | 1.77E-01 | 9.82E-02 | 6.65E-10 |
| rs580900 | 6 | 111936275 | 2634 | rs580900, rs17073074, rs671271, rs17754910, rs490080 | AGGAC | 1.38E-01 | 1.77E-01 | 9.81E-02 | 6.89E-10 |
| rs580900 | 6 | 111936275 | 2634 | rs580900, rs17073074, rs671271, rs17754910, rs490080, rs1327199 | AGGACG | 1.37E-01 | 1.76E-01 | 9.80E-02 | 8.52E-10 |
| rs17073074 | 6 | 111938273 | 2635 | rs17073074, rs671271, rs17754910 | GGA | 1.38E-01 | 1.77E-01 | 9.84E-02 | 6.53E-10 |
| rs17073074 | 6 | 111938273 | 2635 | rs17073074, rs671271, rs17754910, rs490080 | GGAC | 1.38E-01 | 1.77E-01 | 9.84E-02 | 6.83E-10 |
| rs17073074 | 6 | 111938273 | 2635 | rs17073074, rs671271, rs17754910, rs490080, rs1327199 | GGACG | 1.37E-01 | 1.76E-01 | 9.80E-02 | 7.91E-10 |
| rs17073074 | 6 | 111938273 | 2635 | rs17073074, rs671271, rs17754910, rs490080, rs1327199, rs9487771 | GGACGA | 1.37E-01 | 1.75E-01 | 9.73E-02 | 6.87E-10 |
| rs671271 | 6 | 111940182 | 2636 | rs671271, rs17754910 | GA | 1.38E-01 | 1.77E-01 | 9.84E-02 | 6.45E-10 |
| rs671271 | 6 | 111940182 | 2636 | rs671271, rs17754910, rs490080 | GAC | 1.38E-01 | 1.77E-01 | 9.84E-02 | 6.47E-10 |
| rs671271 | 6 | 111940182 | 2636 | rs671271, rs17754910, rs490080, rs1327199 | GACG | 1.37E-01 | 1.76E-01 | 9.79E-02 | 6.36E-10 |
| rs671271 | 6 | 111940182 | 2636 | rs671271, rs17754910, rs490080, rs1327199, rs9487771 | GACGA | 1.37E-01 | 1.75E-01 | 9.72E-02 | 5.47E-10 |
| rs671271 | 6 | 111940182 | 2636 | rs671271, rs17754910, rs490080, rs1327199, rs9487771, rs585057 | GACGAA | 1.34E-01 | 1.73E-01 | 9.42E-02 | 2.73E-10 |
| rs17754910 | 6 | 111942528 | 2637 | rs17754910, rs490080 | AC | 1.38E-01 | 1.77E-01 | 9.83E-02 | 6.93E-10 |
| rs17754910 | 6 | 111942528 | 2637 | rs17754910, rs490080 | GC | 8.39E-01 | 8.00E-01 | 8.78E-01 | 9.38E-09 |
| rs17754910 | 6 | 111942528 | 2637 | rs17754910, rs490080, rs1327199 | ACG | 1.37E-01 | 1.75E-01 | 9.75E-02 | 8.68E-10 |
| rs17754910 | 6 | 111942528 | 2637 | rs17754910, rs490080, rs1327199 | GCG | 8.19E-01 | 7.78E-01 | 8.61E-01 | 8.55E-09 |
| rs17754910 | 6 | 111942528 | 2637 | rs17754910, rs490080, rs1327199, rs9487771 | ACGA | 1.36E-01 | 1.75E-01 | 9.62E-02 | 5.97E-10 |
| rs17754910 | 6 | 111942528 | 2637 | rs17754910, rs490080, rs1327199, rs9487771, rs585057 | ACGAA | 1.34E-01 | 1.73E-01 | 9.41E-02 | 3.50E-10 |

| | | | | | | | | | |
|---|---|---|---|---|---|---|---|---|---|
| rs17754910 | 6 | 111942528 | 2637 | rs17754910, rs490080, rs1327199, rs9487771, rs585057, rs689328 | ACGAAG | 1.34E-01 | 1.73E-01 | 9.41E-02 | 3.34E-10 |
| rs490080 | 6 | 111944006 | 2638 | rs490080, rs1327199, rs9487771, rs585057 | CGAA | 2.08E-01 | 2.51E-01 | 1.63E-01 | 3.96E-09 |
| rs490080 | 6 | 111944006 | 2638 | rs490080, rs1327199, rs9487771, rs585057, rs689328 | CGAAG | 2.07E-01 | 2.50E-01 | 1.62E-01 | 3.85E-09 |
| rs490080 | 6 | 111944006 | 2638 | rs490080, rs1327199, rs9487771, rs585057, rs689328, rs546913 | CGAAGG | 2.07E-01 | 2.50E-01 | 1.62E-01 | 3.58E-09 |
| rs1327199 | 6 | 111954157 | 2639 | rs1327199, rs9487771, rs585057 | GAA | 2.30E-01 | 2.73E-01 | 1.86E-01 | 2.65E-08 |
| rs1327199 | 6 | 111954157 | 2639 | rs1327199, rs9487771, rs585057, rs689328 | GAAG | 2.07E-01 | 2.51E-01 | 1.63E-01 | 5.57E-09 |
| rs1327199 | 6 | 111954157 | 2639 | rs1327199, rs9487771, rs585057, rs689328, rs546913 | GAAGG | 2.07E-01 | 2.51E-01 | 1.63E-01 | 5.30E-09 |
| rs1327199 | 6 | 111954157 | 2639 | rs1327199, rs9487771, rs585057, rs689328, rs546913, rs9387058 | GAAGGG | 2.06E-01 | 2.49E-01 | 1.61E-01 | 4.38E-09 |
| rs9487771 | 6 | 111960074 | 2640 | rs9487771, rs585057 | AA | 2.30E-01 | 2.73E-01 | 1.86E-01 | 2.60E-08 |
| rs9487771 | 6 | 111960074 | 2640 | rs9487771, rs585057, rs689328 | AAG | 2.07E-01 | 2.49E-01 | 1.63E-01 | 1.00E-08 |
| rs9487771 | 6 | 111960074 | 2640 | rs9487771, rs585057, rs689328, rs546913 | AAGG | 2.07E-01 | 2.49E-01 | 1.63E-01 | 1.00E-08 |
| rs9487771 | 6 | 111960074 | 2640 | rs9487771, rs585057, rs689328, rs546913, rs9387058 | AAGGG | 2.05E-01 | 2.49E-01 | 1.61E-01 | 4.64E-09 |
| rs9487771 | 6 | 111960074 | 2640 | rs9487771, rs585057, rs689328, rs546913, rs9387058 | AAGGG | 2.05E-01 | 2.49E-01 | 1.61E-01 | 4.64E-09 |
| rs585057 | 6 | 111964664 | 2641 | rs585057, rs689328 | AG | 3.21E-01 | 3.56E-01 | 2.85E-01 | 4.31E-05 |
| rs585057 | 6 | 111964664 | 2641 | rs585057, rs689328, rs546913 | AGG | 3.21E-01 | 3.56E-01 | 2.85E-01 | 4.02E-05 |
| rs585057 | 6 | 111964664 | 2641 | rs585057, rs689328, rs546913, rs9387058 | AGGG | 2.15E-01 | 2.58E-01 | 1.71E-01 | 8.83E-09 |
| rs585057 | 6 | 111964664 | 2641 | rs585057, rs689328, rs546913, rs9387058 | AGGG | 2.15E-01 | 2.58E-01 | 1.71E-01 | 8.83E-09 |
| rs585057 | 6 | 111964664 | 2641 | rs585057, rs689328, rs546913, rs9387058 | AGGG | 2.15E-01 | 2.58E-01 | 1.71E-01 | 8.83E-09 |
| rs2286294 | 7 | 42172250 | 2742 | rs2286294, rs3801214, rs9886211, rs3779173, rs12540671, rs2108368 | AAAGGA | 2.16E-02 | 9.26E-03 | 3.42E-02 | 3.88E-06 |
| rs3801214 | 7 | 42173472 | 2743 | rs3801214, rs9886211, rs3779173, rs12540671, rs2108368 | AAGGA | 2.70E-02 | 1.43E-02 | 4.01E-02 | 1.82E-05 |
| rs3801214 | 7 | 42173472 | 2743 | rs3801214, rs9886211, rs3779173, rs12540671, rs2108368, rs10951671 | AAGGAA | 2.70E-02 | 1.43E-02 | 4.01E-02 | 1.83E-05 |
| rs9886211 | 7 | 42177428 | 2744 | rs9886211, rs3779173, rs12540671, rs2108368 | AGGA | 2.69E-02 | 1.42E-02 | 3.99E-02 | 1.92E-05 |
| rs9886211 | 7 | 42177428 | 2744 | rs9886211, rs3779173, rs12540671, rs2108368, rs10951671 | AGGAA | 2.68E-02 | 1.42E-02 | 3.98E-02 | 1.93E-05 |

| | | | | | | | | | |
|---|---|---|---|---|---|---|---|---|---|
| rs9886211 | 7 | 42177428 | 2744 | rs9886211, rs3779173, rs12540671, rs2108368, rs10951671, rs10261063 | AGGAAA | 2.65E-02 | 1.36E-02 | 3.98E-02 | 1.16E-05 |
| rs3779173 | 7 | 42181706 | 2745 | rs3779173, rs12540671, rs2108368 | GGA | 2.71E-02 | 1.43E-02 | 4.02E-02 | 1.65E-05 |
| rs3779173 | 7 | 42181706 | 2745 | rs3779173, rs12540671, rs2108368, rs10951671 | GGAA | 2.71E-02 | 1.43E-02 | 4.02E-02 | 1.65E-05 |
| rs3779173 | 7 | 42181706 | 2745 | rs3779173, rs12540671, rs2108368, rs10951671, rs10261063 | GGAAA | 2.67E-02 | 1.36E-02 | 4.01E-02 | 9.48E-06 |
| rs3779173 | 7 | 42181706 | 2745 | rs3779173, rs12540671, rs2108368, rs10951671, rs10261063, rs3801216 | GGAAAG | 2.69E-02 | 1.39E-02 | 4.02E-02 | 1.22E-05 |
| rs12540671 | 7 | 42182214 | 2746 | rs12540671, rs2108368 | GA | 2.71E-02 | 1.43E-02 | 4.02E-02 | 1.65E-05 |
| rs12540671 | 7 | 42182214 | 2746 | rs12540671, rs2108368, rs10951671 | GAA | 2.71E-02 | 1.43E-02 | 4.02E-02 | 1.65E-05 |
| rs12540671 | 7 | 42182214 | 2746 | rs12540671, rs2108368, rs10951671, rs10261063 | GAAA | 2.71E-02 | 1.43E-02 | 4.02E-02 | 1.66E-05 |
| rs12540671 | 7 | 42182214 | 2746 | rs12540671, rs2108368, rs10951671, rs10261063, rs3801216 | GAAAG | 2.71E-02 | 1.43E-02 | 4.02E-02 | 1.63E-05 |
| rs3735030 | 7 | 131386942 | 3705 | rs3735030, rs3847106 | GG | 8.72E-01 | 8.46E-01 | 8.98E-01 | 2.44E-05 |
| rs3735030 | 7 | 131386942 | 3705 | rs3735030, rs3847106, rs17165599 | GGA | 8.71E-01 | 8.44E-01 | 8.98E-01 | 2.18E-05 |
| rs3847106 | 7 | 131395288 | 3706 | rs3847106, rs17165599 | GA | 8.72E-01 | 8.45E-01 | 8.99E-01 | 1.49E-05 |
| rs1625957 | 7 | 131527672 | 3738 | rs1625957, rs9649034 | AG | 2.61E-01 | 2.95E-01 | 2.26E-01 | 2.57E-05 |
| rs17162012 | 7 | 141246510 | 3804 | rs17162012, rs4726433, rs10256379, rs4726441 | AAGA | 9.20E-02 | 1.14E-01 | 6.97E-02 | 3.79E-05 |
| rs17162012 | 7 | 141246510 | 3804 | rs17162012, rs4726433, rs10256379, rs4726441, rs216996 | AAGAA | 9.11E-02 | 1.13E-01 | 6.88E-02 | 3.38E-05 |
| rs17162012 | 7 | 141246510 | 3804 | rs17162012, rs4726433, rs10256379, rs4726441, rs216996, rs4726443 | AAGAAC | 9.14E-02 | 1.13E-01 | 6.91E-02 | 3.78E-05 |
| rs4726433 | 7 | 141254813 | 3805 | rs4726433, rs10256379, rs4726441 | AGA | 9.33E-02 | 1.15E-01 | 7.10E-02 | 4.15E-05 |
| rs4726433 | 7 | 141254813 | 3805 | rs4726433, rs10256379, rs4726441, rs216996 | AGAA | 9.25E-02 | 1.14E-01 | 7.01E-02 | 3.73E-05 |
| rs4726433 | 7 | 141254813 | 3805 | rs4726433, rs10256379, rs4726441, rs216996, rs4726443 | AGAAC | 9.25E-02 | 1.14E-01 | 7.01E-02 | 4.11E-05 |
| rs4726433 | 7 | 141254813 | 3805 | rs4726433, rs10256379, rs4726441, rs216996, rs4726443, rs216997 | AGAACG | 9.26E-02 | 1.15E-01 | 7.03E-02 | 4.15E-05 |
| rs10256379 | 7 | 141255339 | 3806 | rs10256379, rs4726441 | GA | 9.33E-02 | 1.15E-01 | 7.10E-02 | 4.23E-05 |
| rs10256379 | 7 | 141255339 | 3806 | rs10256379, rs4726441, rs216996 | GAA | 9.25E-02 | 1.14E-01 | 7.02E-02 | 4.01E-05 |
| rs10256379 | 7 | 141255339 | 3806 | rs10256379, rs4726441, rs216996, rs4726443 | GAAC | 9.26E-02 | 1.14E-01 | 7.02E-02 | 4.00E-05 |

| | | | | | | | | | |
|---|---|---|---|---|---|---|---|---|---|
| rs10256379 | 7 | 141255339 | 3806 | rs10256379, rs4726441, rs216996, rs4726443, rs216997 | GAACG | 9.26E-02 | 1.14E-01 | 7.03E-02 | 4.04E-05 |
| rs13262708 | 8 | 1943903 | 3996 | rs13262708, rs17064407, rs17830077, rs2280823, rs2280822, rs17064417 | AAAAAG | 2.85E-02 | 1.52E-02 | 4.20E-02 | 1.20E-05 |
| rs17064407 | 8 | 1949008 | 3997 | rs17064407, rs17830077, rs2280823, rs2280822, rs17064417 | AAAAG | 3.02E-02 | 1.70E-02 | 4.37E-02 | 2.49E-05 |
| rs17064407 | 8 | 1949008 | 3997 | rs17064407, rs17830077, rs2280823, rs2280822, rs17064417, rs17830107 | AAAAGA | 3.02E-02 | 1.69E-02 | 4.37E-02 | 2.31E-05 |
| rs17830077 | 8 | 1949008 | 3998 | rs17830077, rs2280823, rs2280822, rs17064417 | AAAG | 3.02E-02 | 1.70E-02 | 4.37E-02 | 2.57E-05 |
| rs17830077 | 8 | 1949008 | 3998 | rs17830077, rs2280823, rs2280822, rs17064417, rs17830107 | AAAGA | 3.02E-02 | 1.70E-02 | 4.37E-02 | 2.60E-05 |
| rs17830077 | 8 | 1949008 | 3998 | rs17830077, rs2280823, rs2280822, rs17064417, rs17830107, rs17750595 | AAAGAA | 2.94E-02 | 1.56E-02 | 4.35E-02 | 8.32E-06 |
| rs2280823 | 8 | 1949491 | 3999 | rs2280823, rs2280822, rs17064417 | AAG | 3.02E-02 | 1.70E-02 | 4.37E-02 | 2.57E-05 |
| rs2280823 | 8 | 1949491 | 3999 | rs2280823, rs2280822, rs17064417, rs17830107 | AAGA | 3.02E-02 | 1.70E-02 | 4.37E-02 | 2.59E-05 |
| rs2280823 | 8 | 1949491 | 3999 | rs2280823, rs2280822, rs17064417, rs17830107, rs17750595 | AAGAA | 2.94E-02 | 1.56E-02 | 4.35E-02 | 8.26E-06 |
| rs2280823 | 8 | 1949491 | 3999 | rs2280823, rs2280822, rs17064417, rs17830107, rs17750595, rs3779713 | AAGAAG | 2.94E-02 | 1.56E-02 | 4.35E-02 | 7.90E-06 |
| rs2280822 | 8 | 1949561 | 4000 | rs2280822, rs17064417 | AG | 3.02E-02 | 1.70E-02 | 4.37E-02 | 2.62E-05 |
| rs2280822 | 8 | 1949561 | 4000 | rs2280822, rs17064417, rs17830107 | AGA | 3.02E-02 | 1.70E-02 | 4.36E-02 | 2.68E-05 |
| rs2280822 | 8 | 1949561 | 4000 | rs2280822, rs17064417, rs17830107, rs17750595 | AGAA | 2.93E-02 | 1.55E-02 | 4.33E-02 | 9.44E-06 |
| rs2280822 | 8 | 1949561 | 4000 | rs2280822, rs17064417, rs17830107, rs17750595, rs3779713 | AGAAG | 2.93E-02 | 1.56E-02 | 4.33E-02 | 8.94E-06 |
| rs2280822 | 8 | 1949561 | 4000 | rs2280822, rs17064417, rs17830107, rs17750595, rs3779713, rs4876271 | AGAAGG | 2.91E-02 | 1.51E-02 | 4.35E-02 | 5.49E-06 |
| rs17064417 | 8 | 1950381 | 4001 | rs17064417, rs17830107 | GA | 3.01E-02 | 1.70E-02 | 4.36E-02 | 2.61E-05 |
| rs17064417 | 8 | 1950381 | 4001 | rs17064417, rs17830107, rs17750595 | GAA | 2.90E-02 | 1.54E-02 | 4.29E-02 | 1.04E-05 |
| rs17064417 | 8 | 1950381 | 4001 | rs17064417, rs17830107, rs17750595, rs3779713 | GAAG | 2.93E-02 | 1.56E-02 | 4.33E-02 | 9.00E-06 |
| rs17064417 | 8 | 1950381 | 4001 | rs17064417, rs17830107, rs17750595, rs3779713, rs4876271 | GAAGG | 2.91E-02 | 1.51E-02 | 4.34E-02 | 5.40E-06 |

| | | | | | | | | | |
|---|---|---|---|---|---|---|---|---|---|
| rs17064417 | 8 | 1950381 | 4001 | rs17064417, rs17830107, rs17750595, rs3779713, rs4876271, rs7386942 | GAAGGA | 2.81E-02 | 1.51E-02 | 4.14E-02 | 1.31E-05 |
| rs7009635 | 8 | 140584782 | 5933 | rs7009635, rs6985156, rs7843258 | AGA | 1.77E-01 | 2.07E-01 | 1.46E-01 | 1.38E-05 |
| rs7009635 | 8 | 140584782 | 5933 | rs7009635, rs6985156, rs7843258, rs4961271 | AGAG | 1.77E-01 | 2.07E-01 | 1.46E-01 | 1.31E-05 |
| rs7009635 | 8 | 140584782 | 5933 | rs7009635, rs6985156, rs7843258, rs4961271, rs7001653 | AGAGG | 1.76E-01 | 2.06E-01 | 1.44E-01 | 1.01E-05 |
| rs7009635 | 8 | 140584782 | 5933 | rs7009635, rs6985156, rs7843258, rs4961271, rs7001653, rs2176397 | AGAGGG | 1.75E-01 | 2.05E-01 | 1.44E-01 | 1.23E-05 |
| rs6985156 | 8 | 140584924 | 5934 | rs6985156, rs7843258 | GA | 1.85E-01 | 2.16E-01 | 1.54E-01 | 1.83E-05 |
| rs6985156 | 8 | 140584924 | 5934 | rs6985156, rs7843258, rs4961271 | GAG | 1.85E-01 | 2.16E-01 | 1.54E-01 | 1.74E-05 |
| rs6985156 | 8 | 140584924 | 5934 | rs6985156, rs7843258, rs4961271, rs7001653 | GAGG | 1.84E-01 | 2.15E-01 | 1.52E-01 | 1.12E-05 |
| rs6985156 | 8 | 140584924 | 5934 | rs6985156, rs7843258, rs4961271, rs7001653, rs2176397 | GAGGG | 1.84E-01 | 2.15E-01 | 1.52E-01 | 1.06E-05 |
| rs7843258 | 8 | 140591443 | 5935 | rs7843258, rs4961271 | AG | 1.85E-01 | 2.16E-01 | 1.54E-01 | 1.74E-05 |
| rs7843258 | 8 | 140591443 | 5935 | rs7843258, rs4961271 | GA | 8.10E-01 | 7.79E-01 | 8.41E-01 | 1.74E-05 |
| rs7843258 | 8 | 140591443 | 5935 | rs7843258, rs4961271, rs7001653 | AGG | 1.84E-01 | 2.15E-01 | 1.52E-01 | 1.19E-05 |
| rs7843258 | 8 | 140591443 | 5935 | rs7843258, rs4961271, rs7001653, rs2176397 | AGGG | 1.84E-01 | 2.15E-01 | 1.52E-01 | 1.14E-05 |
| rs4961271 | 8 | 140596890 | 5936 | rs4961271, rs7001653 | GG | 1.88E-01 | 2.19E-01 | 1.57E-01 | 1.60E-05 |
| rs4961271 | 8 | 140596890 | 5936 | rs4961271, rs7001653, rs2176397 | GGG | 1.84E-01 | 2.15E-01 | 1.52E-01 | 1.26E-05 |
| rs7035057 | 9 | 76903299 | 6407 | rs7035057, rs10869843, rs3843581, rs17787855, rs10869844 | AGAGA | 1.57E-02 | 2.51E-02 | 6.30E-03 | 4.50E-05 |
| rs10869843 | 9 | 76906820 | 6408 | rs10869843, rs3843581, rs17787855, rs10869844 | GAGA | 1.59E-02 | 2.54E-02 | 6.27E-03 | 3.53E-05 |
| rs3843581 | 9 | 76909840 | 6409 | rs3843581, rs17787855, rs10869844 | AGA | 1.60E-02 | 2.54E-02 | 6.45E-03 | 4.78E-05 |
| rs3843581 | 9 | 76909840 | 6409 | rs3843581, rs17787855, rs10869844, rs1929533 | AGAA | 1.60E-02 | 2.54E-02 | 6.45E-03 | 4.87E-05 |
| rs17787855 | 9 | 76911896 | 6410 | rs17787855, rs10869844, rs1929533, rs10121560 | GAAG | 1.79E-02 | 2.78E-02 | 7.81E-03 | 4.78E-05 |
| rs655500 | 9 | 87956825 | 6464 | rs655500, rs668108, rs1167 | AAG | 8.99E-01 | 8.77E-01 | 9.22E-01 | 4.70E-05 |
| rs655500 | 9 | 87956825 | 6464 | rs655500, rs668108, rs1167 | CGA | 9.51E-02 | 1.17E-01 | 7.28E-02 | 4.70E-05 |
| rs655500 | 9 | 87956825 | 6464 | rs655500, rs668108, rs1167, rs560019 | CGAG | 9.41E-02 | 1.16E-01 | 7.19E-02 | 4.47E-05 |
| rs655500 | 9 | 87956825 | 6464 | rs655500, rs668108, rs1167, rs560019, rs2271992 | CGAGG | 9.42E-02 | 1.16E-01 | 7.19E-02 | 4.42E-05 |

| | | | | | | | | | |
|---|---|---|---|---|---|---|---|---|---|
| rs655500 | 9 | 87956825 | 6464 | rs655500, rs668108, rs1167, rs560019, rs2271992, rs622418 | CGAGGG | 9.42E-02 | 1.16E-01 | 7.19E-02 | 4.43E-05 |
| rs668108 | 9 | 87966510 | 6465 | rs668108, rs1167 | AG | 9.00E-01 | 8.77E-01 | 9.24E-01 | 1.33E-05 |
| rs668108 | 9 | 87966510 | 6465 | rs668108, rs1167 | GA | 9.86E-02 | 1.22E-01 | 7.42E-02 | 1.33E-05 |
| rs668108 | 9 | 87966510 | 6465 | rs668108, rs1167, rs560019 | GAG | 9.76E-02 | 1.21E-01 | 7.33E-02 | 1.26E-05 |
| rs668108 | 9 | 87966510 | 6465 | rs668108, rs1167, rs560019 | AGA | 6.25E-01 | 5.89E-01 | 6.62E-01 | 3.37E-05 |
| rs668108 | 9 | 87966510 | 6465 | rs668108, rs1167, rs560019, rs2271992 | GAGG | 9.76E-02 | 1.21E-01 | 7.33E-02 | 1.25E-05 |
| rs668108 | 9 | 87966510 | 6465 | rs668108, rs1167, rs560019, rs2271992, rs622418 | GAGGG | 9.76E-02 | 1.21E-01 | 7.33E-02 | 1.25E-05 |
| rs668108 | 9 | 87966510 | 6465 | rs668108, rs1167, rs560019, rs2271992, rs622418, rs665983 | GAGGGG | 9.25E-02 | 1.15E-01 | 6.91E-02 | 1.67E-05 |
| rs1167 | 9 | 87966831 | 6466 | rs1167, rs560019 | AG | 9.79E-02 | 1.22E-01 | 7.33E-02 | 9.27E-06 |
| rs1167 | 9 | 87966831 | 6466 | rs1167, rs560019 | GA | 6.26E-01 | 5.89E-01 | 6.63E-01 | 3.15E-05 |
| rs1167 | 9 | 87966831 | 6466 | rs1167, rs560019, rs2271992 | AGG | 9.79E-02 | 1.22E-01 | 7.33E-02 | 9.16E-06 |
| rs1167 | 9 | 87966831 | 6466 | rs1167, rs560019, rs2271992, rs622418 | AGGG | 9.79E-02 | 1.22E-01 | 7.33E-02 | 9.16E-06 |
| rs1167 | 9 | 87966831 | 6466 | rs1167, rs560019, rs2271992, rs622418, rs665983 | AGGGG | 9.28E-02 | 1.16E-01 | 6.91E-02 | 1.23E-05 |
| rs1167 | 9 | 87966831 | 6466 | rs1167, rs560019, rs2271992, rs622418, rs665983, rs28364935 | AGGGGC | 9.28E-02 | 1.16E-01 | 6.91E-02 | 1.13E-05 |
| rs560019 | 9 | 87968606 | 6467 | rs560019, rs2271992 | GG | 3.73E-01 | 4.10E-01 | 3.36E-01 | 3.19E-05 |
| rs560019 | 9 | 87968606 | 6467 | rs560019, rs2271992, rs622418 | GGG | 3.73E-01 | 4.10E-01 | 3.36E-01 | 3.20E-05 |
| rs560019 | 9 | 87968606 | 6467 | rs560019, rs2271992, rs622418, rs665983, rs28364935, rs572992 | GGGGCC | 9.06E-02 | 1.13E-01 | 6.74E-02 | 1.37E-05 |
| rs2271992 | 9 | 87969563 | 6468 | rs2271992, rs622418, rs665983, rs28364935, rs572992 | GGGCC | 9.15E-02 | 1.14E-01 | 6.90E-02 | 2.56E-05 |
| rs2271992 | 9 | 87969563 | 6468 | rs2271992, rs622418, rs665983, rs28364935, rs572992, rs3898637 | GGGCCA | 9.16E-02 | 1.14E-01 | 6.92E-02 | 2.61E-05 |
| rs622418 | 9 | 87971128 | 6469 | rs622418, rs665983, rs28364935, rs572992 | GGCC | 9.91E-02 | 1.21E-01 | 7.64E-02 | 4.71E-05 |
| rs665983 | 9 | 87973988 | 6470 | rs665983, rs28364935, rs572992 | GCC | 1.02E-01 | 1.25E-01 | 7.82E-02 | 2.87E-05 |
| rs665983 | 9 | 87973988 | 6470 | rs665983, rs28364935, rs572992, rs3898637 | GCCA | 1.02E-01 | 1.25E-01 | 7.82E-02 | 3.44E-05 |
| rs665983 | 9 | 87973988 | 6470 | rs665983, rs28364935, rs572992, rs3898637, rs473082 | GCCAC | 1.02E-01 | 1.25E-01 | 7.84E-02 | 3.80E-05 |
| rs665983 | 9 | 87973988 | 6470 | rs665983, rs28364935, rs572992, rs3898637, rs473082, rs10868677 | GCCACG | 1.02E-01 | 1.25E-01 | 7.84E-02 | 3.72E-05 |

| | | | | | | | | | |
|---|---|---|---|---|---|---|---|---|---|
| rs28364935 | 9 | 87974055 | 6471 | rs28364935, rs572992 | CC | 3.53E-01 | 3.93E-01 | 3.13E-01 | 8.57E-06 |
| rs28364935 | 9 | 87974055 | 6471 | rs28364935, rs572992, rs3898637 | CCA | 3.54E-01 | 3.93E-01 | 3.14E-01 | 8.63E-06 |
| rs28364935 | 9 | 87974055 | 6471 | rs28364935, rs572992, rs3898637, rs473082 | CCAC | 3.52E-01 | 3.92E-01 | 3.11E-01 | 7.81E-06 |
| rs28364935 | 9 | 87974055 | 6471 | rs28364935, rs572992, rs3898637, rs473082, rs10868677 | CCACG | 3.52E-01 | 3.91E-01 | 3.12E-01 | 1.08E-05 |
| rs28364935 | 9 | 87974055 | 6471 | rs28364935, rs572992, rs3898637, rs473082, rs10868677, rs652529 | CCACGG | 3.52E-01 | 3.91E-01 | 3.11E-01 | 8.88E-06 |
| rs572992 | 9 | 87975735 | 6472 | rs572992, rs3898637 | CA | 3.54E-01 | 3.93E-01 | 3.15E-01 | 9.87E-06 |
| rs572992 | 9 | 87975735 | 6472 | rs572992, rs3898637, rs473082 | CAC | 3.53E-01 | 3.92E-01 | 3.13E-01 | 7.58E-06 |
| rs572992 | 9 | 87975735 | 6472 | rs572992, rs3898637, rs473082, rs10868677 | CACG | 3.53E-01 | 3.92E-01 | 3.13E-01 | 7.59E-06 |
| rs572992 | 9 | 87975735 | 6472 | rs572992, rs3898637, rs473082, rs10868677, rs652529 | CACGG | 3.53E-01 | 3.93E-01 | 3.12E-01 | 6.54E-06 |
| rs572992 | 9 | 87975735 | 6472 | rs572992, rs3898637, rs473082, rs10868677, rs652529, rs7043672 | CACGGG | 1.18E-01 | 1.43E-01 | 9.26E-02 | 3.16E-05 |
| rs3898637 | 9 | 87986088 | 6473 | rs3898637, rs473082 | AC | 3.72E-01 | 4.10E-01 | 3.34E-01 | 1.89E-05 |
| rs3898637 | 9 | 87986088 | 6473 | rs3898637, rs473082, rs10868677 | ACG | 3.72E-01 | 4.10E-01 | 3.34E-01 | 1.89E-05 |
| rs3898637 | 9 | 87986088 | 6473 | rs3898637, rs473082, rs10868677, rs652529 | ACGG | 3.59E-01 | 3.98E-01 | 3.18E-01 | 6.95E-06 |
| rs3898637 | 9 | 87986088 | 6473 | rs3898637, rs473082, rs10868677, rs652529, rs7043672 | ACGGG | 1.21E-01 | 1.46E-01 | 9.60E-02 | 4.89E-05 |
| rs3898637 | 9 | 87986088 | 6473 | rs3898637, rs473082, rs10868677, rs652529, rs7043672, rs13296013 | ACGGGA | 1.19E-01 | 1.43E-01 | 9.37E-02 | 4.24E-05 |
| rs473082 | 9 | 87988200 | 6474 | rs473082, rs10868677 | CG | 3.72E-01 | 4.10E-01 | 3.34E-01 | 1.89E-05 |
| rs473082 | 9 | 87988200 | 6474 | rs473082, rs10868677, rs652529 | CGG | 3.59E-01 | 3.98E-01 | 3.18E-01 | 6.95E-06 |
| rs473082 | 9 | 87988200 | 6474 | rs473082, rs10868677, rs652529, rs7043672 | CGGG | 1.21E-01 | 1.45E-01 | 9.57E-02 | 4.92E-05 |
| rs473082 | 9 | 87988200 | 6474 | rs473082, rs10868677, rs652529, rs7043672, rs13296013 | CGGGA | 1.18E-01 | 1.42E-01 | 9.40E-02 | 4.35E-05 |
| rs473082 | 9 | 87988200 | 6474 | rs473082, rs10868677, rs652529, rs7043672, rs13296013, rs7864469 | CGGGAA | 1.18E-01 | 1.42E-01 | 9.25E-02 | 2.21E-05 |
| rs10868677 | 9 | 87996957 | 6475 | rs10868677, rs652529 | GG | 3.58E-01 | 3.98E-01 | 3.18E-01 | 6.95E-06 |
| rs10868677 | 9 | 87996957 | 6475 | rs10868677, rs652529, rs7043672, rs13296013 | GGGA | 1.17E-01 | 1.41E-01 | 9.30E-02 | 4.05E-05 |
| rs10868677 | 9 | 87996957 | 6475 | rs10868677, rs652529, rs7043672, rs13296013, rs7864469 | GGGAA | 1.16E-01 | 1.41E-01 | 9.14E-02 | 2.01E-05 |
| rs652529 | 9 | 87999000 | 6476 | rs652529, rs7043672 | GG | 1.20E-01 | 1.44E-01 | 9.47E-02 | 3.81E-05 |
| rs652529 | 9 | 87999000 | 6476 | rs652529, rs7043672, rs13296013 | GGA | 1.17E-01 | 1.42E-01 | 9.28E-02 | 3.15E-05 |

| | | | | | | | | | |
|---|---|---|---|---|---|---|---|---|---|
| rs652529 | 9 | 87999000 | 6476 | rs652529, rs7043672, rs13296013, rs7864469 | GGAA | 1.17E-01 | 1.41E-01 | 9.20E-02 | 2.12E-05 |
| rs7849325 | 9 | 107148803 | 6608 | rs7849325, rs784677 | AA | 3.84E-01 | 3.47E-01 | 4.21E-01 | 3.55E-05 |
| rs7849325 | 9 | 107148803 | 6608 | rs7849325, rs784677, rs784676 | AAC | 3.84E-01 | 3.47E-01 | 4.22E-01 | 3.61E-05 |
| rs784677 | 9 | 107150170 | 6609 | rs784677, rs784676 | AC | 3.86E-01 | 3.49E-01 | 4.23E-01 | 4.06E-05 |
| rs784670 | 9 | 107153435 | 6611 | rs784670, rs908725 | GA | 6.16E-01 | 6.52E-01 | 5.78E-01 | 4.71E-05 |
| rs2418267 | 9 | 114128316 | 6664 | rs2418267, rs13300798, rs891721, rs891720, rs1017360, rs7868176 | GGACGA | 1.50E-01 | 1.81E-01 | 1.19E-01 | 2.77E-06 |
| rs13300798 | 9 | 114133107 | 6665 | rs13300798, rs891721, rs891720, rs1017360, rs7868176 | GACGA | 1.56E-01 | 1.85E-01 | 1.26E-01 | 9.86E-06 |
| rs13300798 | 9 | 114133107 | 6665 | rs13300798, rs891721, rs891720, rs1017360, rs7868176, rs737174 | GACGAC | 1.51E-01 | 1.81E-01 | 1.19E-01 | 2.56E-06 |
| rs891721 | 9 | 114134686 | 6666 | rs891721, rs891720, rs1017360, rs7868176 | ACGA | 1.52E-01 | 1.79E-01 | 1.24E-01 | 3.40E-05 |
| rs891721 | 9 | 114134686 | 6666 | rs891721, rs891720, rs1017360, rs7868176, rs737174 | ACGAC | 1.50E-01 | 1.81E-01 | 1.19E-01 | 2.44E-06 |
| rs891721 | 9 | 114134686 | 6666 | rs891721, rs891720, rs1017360, rs7868176, rs737174, rs2043195 | ACGACG | 1.46E-01 | 1.76E-01 | 1.15E-01 | 2.53E-06 |
| rs1017360 | 9 | 114139646 | 6668 | rs1017360, rs7868176, rs737174, rs2043195, rs10817573, rs2002284 | GACGGA | 1.42E-01 | 1.71E-01 | 1.14E-01 | 1.10E-05 |
| rs7868176 | 9 | 114140502 | 6669 | rs7868176, rs737174, rs2043195, rs10817573, rs2002284 | ACGGA | 1.42E-01 | 1.71E-01 | 1.14E-01 | 1.15E-05 |
| rs737174 | 9 | 114141130 | 6670 | rs737174, rs2043195, rs10817573, rs2002284 | CGGA | 1.42E-01 | 1.71E-01 | 1.14E-01 | 1.15E-05 |
| rs2043195 | 9 | 114142254 | 6671 | rs2043195, rs10817573, rs2002284 | GGA | 1.42E-01 | 1.70E-01 | 1.13E-01 | 1.19E-05 |
| rs10817573 | 9 | 114145168 | 6672 | rs10817573, rs2002284 | GA | 1.43E-01 | 1.71E-01 | 1.14E-01 | 8.87E-06 |
| rs7427 | 10 | 23119349 | 6951 | rs7427, rs6482254 | GG | 3.08E-01 | 3.43E-01 | 2.72E-01 | 3.90E-05 |
| rs3847356 | 10 | 70069081 | 7182 | rs3847356, rs4385785, rs10999120, rs10999122, rs10823479, rs16927253 | GGAAGA | 4.13E-02 | 5.62E-02 | 2.60E-02 | 4.04E-05 |
| rs7914787 | 10 | 70102898 | 7199 | rs7914787, rs7917290, rs7905431, rs2271699 | AGGG | 7.89E-02 | 9.92E-02 | 5.82E-02 | 4.33E-05 |
| rs7914787 | 10 | 70102898 | 7199 | rs7914787, rs7917290, rs7905431, rs2271699, rs2271698 | AGGGG | 7.85E-02 | 9.87E-02 | 5.80E-02 | 4.70E-05 |
| rs7917290 | 10 | 70106405 | 7200 | rs7917290, rs7905431, rs2271699 | GGG | 7.90E-02 | 9.93E-02 | 5.83E-02 | 3.99E-05 |
| rs7917290 | 10 | 70106405 | 7200 | rs7917290, rs7905431, rs2271699, rs2271698 | GGGG | 7.90E-02 | 9.93E-02 | 5.83E-02 | 3.99E-05 |
| rs2271699 | 10 | 70108860 | 7202 | rs2271699, rs2271698 | GG | 8.27E-02 | 1.03E-01 | 6.17E-02 | 4.47E-05 |
| rs7074815 | 10 | 99350066 | 7257 | rs7074815, rs11190065, rs3750895, rs12098418, rs11190074, rs17568778 | AAGGAA | 6.85E-02 | 4.82E-02 | 8.93E-02 | 1.42E-05 |

| | | | | | | | | | |
|---|---|---|---|---|---|---|---|---|---|
| rs11190065 | 10 | 99355700 | 7258 | rs11190065, rs3750895, rs12098418, rs11190074, rs17568778 | AGGAA | 6.86E-02 | 4.83E-02 | 8.94E-02 | 1.29E-05 |
| rs11190065 | 10 | 99355700 | 7258 | rs11190065, rs3750895, rs12098418, rs11190074, rs17568778, rs12268069 | AGGAAG | 6.86E-02 | 4.83E-02 | 8.95E-02 | 1.29E-05 |
| rs3750895 | 10 | 99357800 | 7259 | rs3750895, rs12098418, rs11190074, rs17568778 | GGAA | 7.00E-02 | 4.94E-02 | 9.11E-02 | 1.18E-05 |
| rs3750895 | 10 | 99357800 | 7259 | rs3750895, rs12098418, rs11190074, rs17568778, rs12268069 | GGAAG | 6.94E-02 | 4.90E-02 | 9.03E-02 | 1.32E-05 |
| rs3750895 | 10 | 99357800 | 7259 | rs3750895, rs12098418, rs11190074, rs17568778, rs12268069, rs4919304 | GGAAGG | 6.93E-02 | 4.90E-02 | 9.02E-02 | 1.38E-05 |
| rs12098418 | 10 | 99357932 | 7260 | rs12098418, rs11190074, rs17568778 | GAA | 6.90E-02 | 4.81E-02 | 9.04E-02 | 7.05E-06 |
| rs12098418 | 10 | 99357932 | 7260 | rs12098418, rs11190074, rs17568778, rs12268069 | GAAG | 6.93E-02 | 4.86E-02 | 9.05E-02 | 8.94E-06 |
| rs12098418 | 10 | 99357932 | 7260 | rs12098418, rs11190074, rs17568778, rs12268069, rs4919304 | GAAGG | 6.93E-02 | 4.87E-02 | 9.03E-02 | 1.01E-05 |
| rs12098418 | 10 | 99357932 | 7260 | rs12098418, rs11190074, rs17568778, rs12268069, rs4919304, rs1892510 | GAAGGA | 6.93E-02 | 4.88E-02 | 9.03E-02 | 1.14E-05 |
| rs11190074 | 10 | 99361961 | 7261 | rs11190074, rs17568778 | AA | 6.90E-02 | 4.83E-02 | 9.02E-02 | 8.16E-06 |
| rs11190074 | 10 | 99361961 | 7261 | rs11190074, rs17568778, rs12268069 | AAG | 6.89E-02 | 4.84E-02 | 8.99E-02 | 9.85E-06 |
| rs11190074 | 10 | 99361961 | 7261 | rs11190074, rs17568778, rs12268069, rs4919304 | AAGG | 6.90E-02 | 4.88E-02 | 8.97E-02 | 1.22E-05 |
| rs11190074 | 10 | 99361961 | 7261 | rs11190074, rs17568778, rs12268069, rs4919304, rs1892510 | AAGGA | 6.91E-02 | 4.89E-02 | 8.98E-02 | 1.10E-05 |
| rs11190074 | 10 | 99361961 | 7261 | rs11190074, rs17568778, rs12268069, rs4919304, rs1892510, rs10509735 | AAGGAC | 6.86E-02 | 4.87E-02 | 8.90E-02 | 1.50E-05 |
| rs17568778 | 10 | 99368605 | 7262 | rs17568778, rs12268069 | AG | 7.13E-02 | 5.10E-02 | 9.22E-02 | 1.56E-05 |
| rs17568778 | 10 | 99368605 | 7262 | rs17568778, rs12268069, rs4919304 | AGG | 7.13E-02 | 5.10E-02 | 9.21E-02 | 1.47E-05 |
| rs17568778 | 10 | 99368605 | 7262 | rs17568778, rs12268069, rs4919304, rs1892510 | AGGA | 7.11E-02 | 5.08E-02 | 9.18E-02 | 1.44E-05 |
| rs17568778 | 10 | 99368605 | 7262 | rs17568778, rs12268069, rs4919304, rs1892510, rs10509735 | AGGAC | 7.03E-02 | 5.05E-02 | 9.05E-02 | 1.81E-05 |
| rs17568778 | 10 | 99368605 | 7262 | rs17568778, rs12268069, rs4919304, rs1892510, rs10509735, rs7084684 | AGGACA | 7.03E-02 | 5.01E-02 | 9.11E-02 | 1.39E-05 |
| rs2289711 | 12 | 27524572 | 7479 | rs2289711, rs2254279, rs7971294, rs306655, rs306664 | CGAGA | 2.85E-01 | 3.19E-01 | 2.51E-01 | 3.71E-05 |

| | | | | | | | | | |
|---|---|---|---|---|---|---|---|---|---|
| rs2289711 | 12 | 27524572 | 7479 | rs2289711, rs2254279, rs7971294, rs306655, rs306664, rs753891 | CGAGAA | 2.85E-01 | 3.19E-01 | 2.51E-01 | 3.12E-05 |
| rs2254279 | 12 | 27524884 | 7480 | rs2254279, rs7971294, rs306655, rs306664, rs753891 | GAGAA | 2.86E-01 | 3.19E-01 | 2.52E-01 | 3.70E-05 |
| rs306655 | 12 | 27539792 | 7482 | rs306655, rs306664 | GG | 5.85E-01 | 5.48E-01 | 6.23E-01 | 3.30E-05 |
| rs306664 | 12 | 27545818 | 7483 | rs306664, rs753891 | AA | 3.59E-01 | 3.96E-01 | 3.20E-01 | 1.71E-05 |
| rs306664 | 12 | 27545818 | 7483 | rs306664, rs753891, rs899069 | AAA | 3.04E-01 | 3.40E-01 | 2.68E-01 | 2.45E-05 |
| rs306664 | 12 | 27545818 | 7483 | rs306664, rs753891, rs899069, rs7963570 | AAAG | 3.04E-01 | 3.40E-01 | 2.68E-01 | 1.90E-05 |
| rs306664 | 12 | 27545818 | 7483 | rs306664, rs753891, rs899069, rs7963570, rs7966993 | AAAGG | 3.04E-01 | 3.40E-01 | 2.68E-01 | 1.69E-05 |
| rs753891 | 12 | 27548131 | 7484 | rs753891, rs899069 | AA | 3.12E-01 | 3.47E-01 | 2.75E-01 | 2.53E-05 |
| rs753891 | 12 | 27548131 | 7484 | rs753891, rs899069, rs7963570 | AAG | 3.12E-01 | 3.48E-01 | 2.75E-01 | 2.54E-05 |
| rs753891 | 12 | 27548131 | 7484 | rs753891, rs899069, rs7963570, rs7966993 | AAGG | 3.12E-01 | 3.48E-01 | 2.75E-01 | 2.67E-05 |
| rs899069 | 12 | 27550546 | 7485 | rs899069, rs7963570 | AG | 3.12E-01 | 3.48E-01 | 2.75E-01 | 2.54E-05 |
| rs899069 | 12 | 27550546 | 7485 | rs899069, rs7963570, rs7966993 | AGG | 3.12E-01 | 3.48E-01 | 2.75E-01 | 2.54E-05 |
| rs10842934 | 12 | 27565844 | 7489 | rs10842934, rs10743603 | AA | 3.13E-01 | 3.49E-01 | 2.76E-01 | 2.40E-05 |
| rs256662 | 12 | 30804286 | 7592 | rs256662, rs7301387, rs256679 | AGA | 3.35E-01 | 3.73E-01 | 2.97E-01 | 1.28E-05 |
| rs7301387 | 12 | 30807504 | 7593 | rs7301387, rs256679 | GA | 3.35E-01 | 3.73E-01 | 2.97E-01 | 1.31E-05 |
| rs17566447 | 12 | 42536475 | 7686 | rs17566447, rs7316200, rs1452106, rs1669947, rs2406870, rs10785347 | GGGAGG | 2.30E-01 | 1.99E-01 | 2.62E-01 | 4.43E-05 |
| rs7316200 | 12 | 42539126 | 7687 | rs7316200, rs1452106, rs1669947, rs2406870, rs10785347 | GGAGG | 2.31E-01 | 1.99E-01 | 2.64E-01 | 3.11E-05 |
| rs7316200 | 12 | 42539126 | 7687 | rs7316200, rs1452106, rs1669947, rs2406870, rs10785347, rs12581019 | GGAGGG | 2.31E-01 | 1.99E-01 | 2.64E-01 | 3.95E-05 |
| rs1452106 | 12 | 42540905 | 7688 | rs1452106, rs1669947, rs2406870 | GAG | 2.36E-01 | 2.03E-01 | 2.70E-01 | 1.58E-05 |
| rs1452106 | 12 | 42540905 | 7688 | rs1452106, rs1669947, rs2406870, rs10785347 | GAGG | 2.35E-01 | 2.00E-01 | 2.70E-01 | 8.05E-06 |
| rs1452106 | 12 | 42540905 | 7688 | rs1452106, rs1669947, rs2406870, rs10785347, rs12581019 | GAGGG | 2.32E-01 | 1.98E-01 | 2.67E-01 | 8.93E-06 |
| rs1452106 | 12 | 42540905 | 7688 | rs1452106, rs1669947, rs2406870, rs10785347, rs12581019, rs11181563 | GAGGGG | 2.27E-01 | 1.92E-01 | 2.61E-01 | 7.27E-06 |
| rs1669947 | 12 | 42548432 | 7689 | rs1669947, rs2406870, rs10785347, rs12581019, rs11181563 | AGGGG | 2.53E-01 | 2.18E-01 | 2.89E-01 | 7.54E-06 |
| rs1669947 | 12 | 42548432 | 7689 | rs1669947, rs2406870, rs10785347, rs12581019, rs11181563, rs11181566 | AGGGGA | 2.45E-01 | 2.13E-01 | 2.78E-01 | 1.84E-05 |

| | | | | | | | | | |
|---|---|---|---|---|---|---|---|---|---|
| rs12873430 | 13 | 26971307 | 7871 | rs12873430, rs12875586, rs9581791, rs2421035, rs9512476 | GAGAA | 3.73E-01 | 3.37E-01 | 4.10E-01 | 4.74E-05 |
| rs12875586 | 13 | 26971748 | 7872 | rs12875586, rs9581791, rs2421035, rs9512476 | AGAA | 3.74E-01 | 3.38E-01 | 4.12E-01 | 4.97E-05 |
| rs4780171 | 15 | 33738664 | 8759 | rs4780171, rs2288608 | AG | 3.72E-01 | 3.32E-01 | 4.12E-01 | 1.13E-05 |
| rs4780171 | 15 | 33738664 | 8759 | rs4780171, rs2288608, rs2288609 | AGG | 3.70E-01 | 3.32E-01 | 4.09E-01 | 1.49E-05 |
| rs4461048 | 15 | 38756030 | 8919 | rs4461048, rs2958267, rs4608310, rs2912367 | GGGG | 1.48E-01 | 1.77E-01 | 1.18E-01 | 9.04E-06 |
| rs4461048 | 15 | 38756030 | 8919 | rs4461048, rs2958267, rs4608310, rs2912367, rs2929655 | GGGGA | 1.46E-01 | 1.74E-01 | 1.18E-01 | 1.39E-05 |
| rs4461048 | 15 | 38756030 | 8919 | rs4461048, rs2958267, rs4608310, rs2912367, rs2929655, rs2929657 | GGGGAA | 1.46E-01 | 1.73E-01 | 1.18E-01 | 2.30E-05 |
| rs2958267 | 15 | 38758551 | 8920 | rs2958267, rs4608310, rs2912367 | GGG | 1.57E-01 | 1.88E-01 | 1.26E-01 | 5.09E-06 |
| rs2958267 | 15 | 38758551 | 8920 | rs2958267, rs4608310, rs2912367, rs2929655 | GGGA | 1.54E-01 | 1.84E-01 | 1.24E-01 | 6.48E-06 |
| rs2958267 | 15 | 38758551 | 8920 | rs2958267, rs4608310, rs2912367, rs2929655, rs2929657 | GGGAA | 1.54E-01 | 1.83E-01 | 1.24E-01 | 9.25E-06 |
| rs2958267 | 15 | 38758551 | 8920 | rs2958267, rs4608310, rs2912367, rs2929655, rs2929657, rs16967429 | GGGAAA | 1.54E-01 | 1.83E-01 | 1.24E-01 | 8.89E-06 |
| rs4608310 | 15 | 38759783 | 8921 | rs4608310, rs2912367 | GG | 1.57E-01 | 1.88E-01 | 1.26E-01 | 5.10E-06 |
| rs4608310 | 15 | 38759783 | 8921 | rs4608310, rs2912367, rs2929655 | GGA | 1.54E-01 | 1.84E-01 | 1.24E-01 | 6.48E-06 |
| rs4608310 | 15 | 38759783 | 8921 | rs4608310, rs2912367, rs2929655, rs2929657 | GGAA | 1.54E-01 | 1.83E-01 | 1.24E-01 | 9.23E-06 |
| rs4608310 | 15 | 38759783 | 8921 | rs4608310, rs2912367, rs2929655, rs2929657, rs16967429 | GGAAA | 1.53E-01 | 1.82E-01 | 1.24E-01 | 9.40E-06 |
| rs4608310 | 15 | 38759783 | 8921 | rs4608310, rs2912367, rs2929655, rs2929657, rs16967429, rs2929660 | GGAAAG | 1.52E-01 | 1.80E-01 | 1.24E-01 | 2.66E-05 |
| rs2912367 | 15 | 38763383 | 8922 | rs2912367, rs2929655 | AG | 8.15E-01 | 7.81E-01 | 8.50E-01 | 1.85E-06 |
| rs2912367 | 15 | 38763383 | 8922 | rs2912367, rs2929655 | GA | 1.54E-01 | 1.84E-01 | 1.24E-01 | 6.47E-06 |
| rs2912367 | 15 | 38763383 | 8922 | rs2912367, rs2929655, rs2929657 | GAA | 1.54E-01 | 1.83E-01 | 1.24E-01 | 9.33E-06 |
| rs2912367 | 15 | 38763383 | 8922 | rs2912367, rs2929655, rs2929657 | AGC | 7.85E-01 | 7.53E-01 | 8.18E-01 | 3.64E-05 |
| rs2912367 | 15 | 38763383 | 8922 | rs2912367, rs2929655, rs2929657, rs16967429 | GAAA | 1.54E-01 | 1.83E-01 | 1.24E-01 | 9.27E-06 |
| rs2912367 | 15 | 38763383 | 8922 | rs2912367, rs2929655, rs2929657, rs16967429, rs2929660 | GAAAG | 1.52E-01 | 1.80E-01 | 1.24E-01 | 2.34E-05 |
| rs2912367 | 15 | 38763383 | 8922 | rs2912367, rs2929655, rs2929657, rs16967429, rs2929660, rs11636712 | GAAAGG | 1.52E-01 | 1.80E-01 | 1.24E-01 | 2.33E-05 |
| rs2929655 | 15 | 38765644 | 8923 | rs2929655, rs2929657 | AA | 1.81E-01 | 2.14E-01 | 1.48E-01 | 2.88E-06 |
| rs2929655 | 15 | 38765644 | 8923 | rs2929655, rs2929657 | GC | 7.86E-01 | 7.54E-01 | 8.18E-01 | 2.89E-05 |

| | | | | | | | | | |
|---|---|---|---|---|---|---|---|---|---|
| rs2929655 | 15 | 38765644 | 8923 | rs2929655, rs2929657, rs16967429 | AAA | 1.54E-01 | 1.83E-01 | 1.24E-01 | 7.85E-06 |
| rs2929655 | 15 | 38765644 | 8923 | rs2929655, rs2929657, rs16967429, rs2929660 | AAAG | 1.53E-01 | 1.81E-01 | 1.24E-01 | 1.64E-05 |
| rs2929655 | 15 | 38765644 | 8923 | rs2929655, rs2929657, rs16967429, rs2929660, rs11636712 | AAAGG | 1.52E-01 | 1.80E-01 | 1.23E-01 | 1.60E-05 |
| rs2929655 | 15 | 38765644 | 8923 | rs2929655, rs2929657, rs16967429, rs2929660, rs11636712 | AAAGG | 1.52E-01 | 1.80E-01 | 1.23E-01 | 1.60E-05 |
| rs2929660 | 15 | 38773223 | 8926 | rs2929660, rs11636712 | GG | 1.87E-01 | 2.17E-01 | 1.57E-01 | 3.05E-05 |
| rs2929660 | 15 | 38773223 | 8926 | rs2929660, rs11636712 | GG | 1.87E-01 | 2.17E-01 | 1.57E-01 | 3.05E-05 |
| rs2929660 | 15 | 38773223 | 8926 | rs2929660, rs11636712 | GG | 1.87E-01 | 2.17E-01 | 1.57E-01 | 3.05E-05 |
| rs2929660 | 15 | 38773223 | 8926 | rs2929660, rs11636712 | GG | 1.87E-01 | 2.17E-01 | 1.57E-01 | 3.05E-05 |
| rs2929660 | 15 | 38773223 | 8926 | rs2929660, rs11636712 | GG | 1.87E-01 | 2.17E-01 | 1.57E-01 | 3.05E-05 |
| rs9909462 | 17 | 80182492 | 9337 | rs9909462, rs9902358, rs4889990, rs4889991, rs2044102, rs2044103 | GCAACG | 9.38E-02 | 7.17E-02 | 1.16E-01 | 4.65E-05 |
| rs4889990 | 17 | 80184196 | 9339 | rs4889990, rs4889991, rs2044102, rs2044103, rs8065364 | AACGA | 9.20E-02 | 6.98E-02 | 1.15E-01 | 2.94E-05 |
| rs4889990 | 17 | 80184196 | 9339 | rs4889990, rs4889991, rs2044102, rs2044103, rs8065364, rs11655682 | AACGAA | 9.19E-02 | 6.92E-02 | 1.15E-01 | 1.80E-05 |
| rs4889991 | 17 | 80184264 | 9340 | rs4889991, rs2044102, rs2044103, rs8065364 | ACGA | 1.49E-01 | 1.18E-01 | 1.80E-01 | 2.48E-06 |
| rs4889991 | 17 | 80184264 | 9340 | rs4889991, rs2044102, rs2044103, rs8065364, rs11655682 | ACGAA | 9.89E-02 | 7.54E-02 | 1.23E-01 | 1.61E-05 |
| rs4889991 | 17 | 80184264 | 9340 | rs4889991, rs2044102, rs2044103, rs8065364, rs11655682, rs8068433 | ACGAAA | 9.51E-02 | 7.03E-02 | 1.20E-01 | 3.06E-06 |
| rs2044102 | 17 | 80185561 | 9341 | rs2044102, rs2044103, rs8065364 | CGA | 1.49E-01 | 1.18E-01 | 1.80E-01 | 2.47E-06 |
| rs2044102 | 17 | 80185561 | 9341 | rs2044102, rs2044103, rs8065364, rs11655682 | CGAA | 9.90E-02 | 7.57E-02 | 1.23E-01 | 1.83E-05 |
| rs2044102 | 17 | 80185561 | 9341 | rs2044102, rs2044103, rs8065364, rs11655682, rs8068433 | CGAAA | 9.50E-02 | 7.03E-02 | 1.20E-01 | 3.17E-06 |
| rs2044102 | 17 | 80185561 | 9341 | rs2044102, rs2044103, rs8065364, rs11655682, rs8068433, rs8068452 | CGAAAA | 9.51E-02 | 6.99E-02 | 1.21E-01 | 2.16E-06 |
| rs8110480 | 19 | 5195906 | 9523 | rs8110480, rs8105746 | GA | 1.42E-01 | 1.70E-01 | 1.14E-01 | 1.71E-05 |
| rs8110480 | 19 | 5195906 | 9523 | rs8110480, rs8105746, rs11880284 | GAG | 1.37E-01 | 1.63E-01 | 1.11E-01 | 3.53E-05 |
| rs8110480 | 19 | 5195906 | 9523 | rs8110480, rs8105746, rs11880284, rs737004 | GAGA | 1.33E-01 | 1.59E-01 | 1.06E-01 | 2.19E-05 |
| rs8110480 | 19 | 5195906 | 9523 | rs8110480, rs8105746, rs11880284, rs737004, rs2302224 | GAGAA | 1.13E-01 | 1.38E-01 | 8.81E-02 | 1.44E-05 |

| | | | | | | | | | |
|---|---|---|---|---|---|---|---|---|---|
| rs8110480 | 19 | 5195906 | 9523 | rs8110480, rs8105746, rs11880284, rs737004, rs2302224, rs1143699 | GAGAAA | 1.10E-01 | 1.33E-01 | 8.72E-02 | 4.95E-05 |
| rs8105746 | 19 | 5200845 | 9524 | rs8105746, rs11880284, rs737004 | AGA | 1.33E-01 | 1.58E-01 | 1.06E-01 | 3.12E-05 |
| rs8105746 | 19 | 5200856 | 9524 | rs8105746, rs11880284, rs737004, rs2302224 | AGAA | 1.13E-01 | 1.38E-01 | 8.78E-02 | 1.56E-05 |
| rs17210722 | 21 | 17169100 | 9774 | rs17210722, rs11702385, rs2824249, rs2824253, rs13047117, rs1501811 | AGCAGA | 1.92E-01 | 1.61E-01 | 2.24E-01 | 1.65E-05 |
| rs11702385 | 21 | 17169191 | 9775 | rs11702385, rs2824249, rs2824253, rs13047117, rs1501811 | GCAGA | 1.92E-01 | 1.61E-01 | 2.24E-01 | 1.65E-05 |
| rs11702385 | 21 | 17169191 | 9775 | rs11702385, rs2824249, rs2824253, rs13047117, rs1501811, rs1604416 | GCAGAA | 1.90E-01 | 1.58E-01 | 2.21E-01 | 1.28E-05 |
| rs2824249 | 21 | 17170030 | 9776 | rs2824249, rs2824253, rs13047117, rs1501811 | CAGA | 1.92E-01 | 1.61E-01 | 2.23E-01 | 1.72E-05 |
| rs2824249 | 21 | 17170030 | 9776 | rs2824249, rs2824253, rs13047117, rs1501811, rs1604416 | CAGAA | 1.89E-01 | 1.58E-01 | 2.21E-01 | 1.34E-05 |
| rs2824249 | 21 | 17170030 | 9776 | rs2824249, rs2824253, rs13047117, rs1501811, rs1604416, rs202892 | CAGAAG | 1.89E-01 | 1.58E-01 | 2.20E-01 | 1.50E-05 |
| rs2824253 | 21 | 17171821 | 9777 | rs2824253, rs13047117, rs1501811 | AGA | 1.91E-01 | 1.60E-01 | 2.23E-01 | 1.30E-05 |
| rs2824253 | 21 | 17171821 | 9777 | rs2824253, rs13047117, rs1501811, rs1604416 | AGAA | 1.89E-01 | 1.57E-01 | 2.21E-01 | 1.11E-05 |
| rs2824253 | 21 | 17171821 | 9777 | rs2824253, rs13047117, rs1501811, rs1604416, rs202892 | AGAAG | 1.89E-01 | 1.57E-01 | 2.21E-01 | 1.12E-05 |
| rs2824253 | 21 | 17171821 | 9777 | rs2824253, rs13047117, rs1501811, rs1604416, rs202892, rs7282090 | AGAAGG | 1.89E-01 | 1.58E-01 | 2.21E-01 | 1.11E-05 |
| rs367799 | 21 | 28216295 | 9862 | rs367799, rs420178 | GG | 4.67E-02 | 6.33E-02 | 2.98E-02 | 1.94E-05 |
| rs367799 | 21 | 28216295 | 9862 | rs367799, rs420178, rs2409294 | GGA | 4.67E-02 | 6.33E-02 | 2.98E-02 | 1.94E-05 |
| rs367799 | 21 | 28216295 | 9862 | rs367799, rs420178, rs2409294, rs2205243 | GGAC | 4.66E-02 | 6.30E-02 | 2.98E-02 | 2.26E-05 |
| rs367799 | 21 | 28216295 | 9862 | rs367799, rs420178, rs2409294, rs2205243, rs2831668 | GGACC | 4.56E-02 | 6.23E-02 | 2.85E-02 | 1.25E-05 |

**Table S1. Sliding window haplotypes of 2-6 SNVs in length meeting a threshold of 5 x 10^-5.** First marker = first SNV defining the haplotype; CHR = chromosome; POS = position; Block = block number; Markers Used = combined markers defining the haplotype; Haplotype = haplotype sequence; EM Freq = haplotype frequency; Case = haplotype frequency within patients diagnosed at ≤ 45 years of age; Control = haplotype frequency within patients at > 45 years-of-age; P = $\chi^2$ p value.
