## Supplemental Table S3 for "Retrospective analysis of *The Two Sister Study* using haplotype-based association testing to identify loci associated with early-onset breast cancer"

**Supplemental Table S3. Nested haploblock analysis.**

| Haploblock clusters | Most Significant Haploblock | Most Significant Haplotype | EM Freq | Case | Control | P |
|---|---|---|---|---|---|---|
| 57-59 | 57 | AGGA | 0.3422 | 0.3032 | 0.3819 | 1.36E-05 |
| 162-163 | 163 | AA | 0.6638 | 0.7000 | 0.6269 | 2.99E-05 |
| 349-342 | 342 | AGGA | 0.0120 | 0.0204 | 0.0035 | 2.74E-05 |
| 574-577 | 576 | GCGGGA | 0.0116 | 0.0203 | 0.0028 | 1.02E-05 |
| 622-632 | 629 | GAGA | 0.1665 | 0.1371 | 0.1964 | 1.83E-05 |
| 695-697 | 697 | GGCAA | 0.3743 | 0.4111 | 0.3368 | 3.20E-05 |
| 1535-1539 | 1538 | GAGA | 0.6820 | 0.7202 | 0.6429 | 4.46E-06 |
| 1549-1554 | 1549 | AATGA | 0.6984 | 0.7404 | 0.6554 | 5.73E-07 |
| 1562-1566 | 1562 | AGGA | 0.2957 | 0.2568 | 0.3353 | 3.44E-06 |
| 1568-1571 | 1570 | GGCGA | 0.2871 | 0.2490 | 0.3259 | 4.44E-06 |
| 2047-2053 | 2052 | GGAGAA | 0.0906 | 0.1142 | 0.0665 | 8.15E-06 |
| 2055-2057 | 2057 | ACG | 0.0678 | 0.0876 | 0.0478 | 1.89E-05 |
| 2326-2329 | 2327 | CGGACG | 0.0427 | 0.0594 | 0.0257 | 6.35E-06 |
| 2632-2641 | 2636 | GACGAA | 0.1340 | 0.1731 | 0.0942 | 2.73E-10 |
| 2742-2746 | 2742 | AAAGGA | 0.0216 | 0.0093 | 0.0342 | 3.88E-06 |
| 3705-3706 | 3706 | GA | 0.8717 | 0.8450 | 0.8988 | 1.49E-05 |
| 3804-3806 | 3804 | AAGAA | 0.0911 | 0.1131 | 0.0688 | 3.38E-05 |
| 3996-4001 | 4001 | GAAGG | 0.0291 | 0.0151 | 0.0434 | 5.40E-06 |
| 5933-5936 | 5933 | AGAGG | 0.1756 | 0.2063 | 0.1444 | 1.01E-05 |
| 6407-6410 | 6407 | AGAGA | 0.0157 | 0.0251 | 0.0063 | 4.50E-05 |
| 6464-6476 | 6472 | CACGG | 0.3527 | 0.3925 | 0.3121 | 6.54E-06 |
| 6608-6609 | 6608 | AA | 0.3837 | 0.3467 | 0.4214 | 3.55E-05 |
| 6664-6672 | 6666 | ACGAC | 0.1502 | 0.1810 | 0.1188 | 2.44E-06 |
| 7199-7200 | 7200 | GGG | 0.0790 | 0.0993 | 0.0583 | 3.99E-05 |
| 7257-7262 | 7260 | GAA | 0.0690 | 0.0481 | 0.0904 | 7.05E-06 |
| 7479-7485 | 7483 | AAAGG | 0.3044 | 0.3399 | 0.2683 | 1.69E-05 |

| | | | | | | |
|---|---|---|---|---|---|---|
| 7592-7593 | 7592 | AGA | 0.3354 | 0.3732 | 0.2968 | 1.28E-05 |
| 7686-7689 | 7688 | GAGGGG | 0.2265 | 0.1925 | 0.2612 | 7.27E-06 |
| 7871-7872 | 7871 | GAGAA | 0.3732 | 0.3367 | 0.4104 | 4.74E-05 |
| 8919-8923 | 8922 | AG | 0.8152 | 0.7809 | 0.8501 | 1.85E-06 |
| 9339-9341 | 9341 | CGAAAA | 0.0951 | 0.0699 | 0.1208 | 2.16E-06 |
| 9523-9524 | 9523 | GAGAA | 0.1135 | 0.1384 | 0.0881 | 1.44E-05 |
| 9774-9777 | 9777 | AGAA | 0.1885 | 0.1572 | 0.2206 | 1.11E-05 |

**Table S3. Nested analysis of haplotype clusters for candidate prioritization.** Filtering of previously defined haplotypes using a threshold of $p \leq 5 \times 10^{-5}$ identified 165 unique haploblocks consisting of 466 unique SNVs. Selecting the most significant haplotype associated with each cluster reduced the number of candidates to 33 chromosomal regions defined by 154 unique SNVs. Haploblock clusters = the series of immediately adjacent haploblocks wherein each block meets the 5 x 10-5 threshold; Most Significant Haploblock = the single haploblock within each cluster achieving the highest degree of significance; Most Significant Haplotype = the most significant haplotype within the most significant haploblock associated with each cluster.
