## Supplemental Table S4 for "Retrospective analysis of *The Two Sister Study* using haplotype-based association testing to identify loci associated with early-onset breast cancer"

**Supplemental Table S4. Marker characteristics.**

| Marker | CHR | POS | Gene | Case Hom Minor | Control Hom Minor | Case Het | Control Het | Case Hom Major | Control Hom Major | Case HWE P | Control HWE P |
|---|---|---|---|---|---|---|---|---|---|---|---|
| rs1885872 | 1 | 3688533 | TP73 | 13 | 3 | 115 | 84 | 607 | 634 | 0.01 | 0.90 |
| rs3765731 | 1 | 3695328 | TP73 | 68 | 53 | 320 | 311 | 347 | 357 | 0.64 | 0.19 |
| rs12032272 | 1 | 3698592 | TP73 | 23 | 20 | 218 | 188 | 494 | 513 | 0.86 | 0.58 |
| rs3765736 | 1 | 3700037 | TP73 | 74 | 117 | 316 | 329 | 345 | 275 | 0.90 | 0.27 |
| rs7425814 | 2 | 149116355 | LYPD6B | 110 | 102 | 341 | 333 | 284 | 284 | 0.64 | 0.78 |
| rs4349313 | 2 | 149118982 | LYPD6B | 4 | 2 | 27 | 26 | 704 | 693 | 0.00 | 0.00 |
| rs6714204 | 2 | 149122269 | LYPD6B | 3 | 2 | 27 | 25 | 705 | 694 | 0.00 | 0.00 |
| rs6720513 | 2 | 149122777 | LYPD6B | 105 | 79 | 350 | 328 | 280 | 314 | 0.79 | 0.63 |
| rs7577550 | 2 | 149123213 | LYPD6B | 19 | 16 | 191 | 182 | 525 | 523 | 0.74 | 0.97 |
| rs10803829 | 2 | 149124999 | LYPD6B | 187 | 154 | 365 | 363 | 183 | 204 | 0.85 | 0.75 |
| rs10024144 | 4 | 122145420 | KIAA1109 (p) | 1 | 3 | 58 | 66 | 676 | 652 | 0.83 | 0.35 |
| rs11938795 | 4 | 122151854 | KIAA1109 (p) | 31 | 78 | 282 | 282 | 422 | 361 | 0.06 | 0.04 |
| rs10005516 | 4 | 122163719 | KIAA1109 (p) | 0 | 2 | 43 | 49 | 692 | 670 | 0.41 | 0.28 |
| rs10471017 | 4 | 122169665 | KIAA1109 (p) | 2 | 1 | 20 | 23 | 713 | 697 | 0.00 | 0.09 |
| rs6848868 | 4 | 122229131 | KIAA1109 coding | 3 | 11 | 106 | 132 | 626 | 578 | 0.51 | 0.28 |
| rs56363411 | 4 | 122250504 | KIAA1109 coding | 1 | 0 | 41 | 43 | 693 | 678 | 0.63 | 0.41 |
| rs45510500 | 4 | 122258745 | KIAA1109 coding | 2 | 1 | 52 | 61 | 680 | 659 | 0.35 | 0.74 |
| rs13119723 | 4 | 122297158 | KIAA1109 coding | 8 | 31 | 154 | 174 | 570 | 512 | 0.50 | 0.00 |
| rs10032704 | 4 | 122385068 | ADAD1 | 125 | 105 | 348 | 331 | 256 | 283 | 0.72 | 0.61 |
| rs17388568 | 4 | 122408207 | ADAD1 | 47 | 35 | 287 | 273 | 401 | 413 | 0.65 | 0.23 |
| rs12499753 | 4 | 122412022 | ADAD1 | 79 | 59 | 332 | 296 | 324 | 365 | 0.66 | 0.93 |
| rs1479923 | 4 | 122449232 | IL2 | 77 | 49 | 324 | 295 | 334 | 377 | 0.90 | 0.39 |
| rs11575812 | 4 | 122449894 | IL2 | 34 | 89 | 299 | 292 | 402 | 340 | 0.02 | 0.04 |
| rs2069763 | 4 | 122456327 | IL2 | 79 | 59 | 329 | 296 | 327 | 366 | 0.78 | 0.94 |
| rs10857092 | 4 | 122468064 | 3' | 6 | 2 | 106 | 73 | 623 | 646 | 0.53 | 0.97 |
| rs6848139 | 4 | 122473886 | 3' | 6 | 14 | 99 | 116 | 630 | 591 | 0.34 | 0.00 |
| rs671271 | 6 | 111940182 | regulatory | 32 | 39 | 182 | 185 | 521 | 497 | 0.00 | 0.00 |

| | | | | | | | | | | | |
|---|---|---|---|---|---|---|---|---|---|---|---|
| rs17754910 | 6 | 111942528 | regulatory | 29 | 13 | 203 | 116 | 503 | 592 | 0.14 | 0.01 |
| rs490080 | 6 | 111944006 | regulatory | 1 | 0 | 32 | 34 | 700 | 687 | 0.32 | 0.52 |
| rs1327199 | 6 | 111954157 | regulatory | 0 | 0 | 35 | 26 | 700 | 695 | 0.51 | 0.62 |
| rs9487771 | 6 | 111960074 | regulatory | 21 | 28 | 173 | 178 | 541 | 514 | 0.12 | 0.01 |
| rs585057 | 6 | 111964664 | intergenic | 138 | 112 | 340 | 279 | 257 | 330 | 0.18 | 0.00 |
| rs17064417 | 8 | 1950381 | ARHGEF10 | 0 | 1 | 25 | 61 | 710 | 659 | 0.64 | 0.74 |
| rs17830107 | 8 | 1951525 | ARHGEF10 | 100 | 107 | 324 | 329 | 311 | 284 | 0.29 | 0.46 |
| rs17750595 | 8 | 1952204 | ARHGEF10 | 122 | 118 | 329 | 328 | 284 | 275 | 0.11 | 0.23 |
| rs3779713 | 8 | 1952244 | ARHGEF10 | 86 | 74 | 337 | 323 | 312 | 324 | 0.73 | 0.62 |
| rs4876271 | 8 | 1953764 | ARHGEF10 | 11 | 11 | 171 | 142 | 553 | 567 | 0.59 | 0.54 |
| rs7009635 | 8 | 140584782 | AGO2 | 151 | 133 | 384 | 349 | 199 | 239 | 0.17 | 0.78 |
| rs6985156 | 8 | 140584924 | AGO2 | 8 | 8 | 144 | 158 | 583 | 555 | 0.79 | 0.38 |
| rs7843258 | 8 | 140591443 | AGO2 | 26 | 16 | 265 | 190 | 444 | 515 | 0.07 | 0.76 |
| rs4961271 | 8 | 140596890 | AGO2 | 27 | 16 | 270 | 197 | 437 | 508 | 0.06 | 0.54 |
| rs7001653 | 8 | 140610939 | AGO2 | 132 | 158 | 384 | 377 | 219 | 186 | 0.11 | 0.20 |
| rs12098418 | 10 | 99357932 | CNNM1 | 14 | 8 | 156 | 148 | 565 | 565 | 0.40 | 0.62 |
| rs11190074 | 10 | 99361961 | CNNM1 | 5 | 5 | 121 | 99 | 609 | 617 | 0.70 | 0.64 |
| rs17568778 | 10 | 99368605 | CNNM1 | 4 | 7 | 67 | 119 | 664 | 594 | 0.11 | 0.70 |
| rs306664 | 12 | 27545818 | PPFIBP1 | 142 | 103 | 381 | 337 | 212 | 281 | 0.21 | 0.90 |
| rs753891 | 12 | 27548131 | PPFIBP1 | 5 | 5 | 110 | 123 | 620 | 593 | 0.96 | 0.61 |
| rs899069 | 12 | 27550546 | PPFIBP1 | 84 | 50 | 343 | 297 | 308 | 374 | 0.43 | 0.39 |
| rs7963570 | 12 | 27553796 | PPFIBP1 | 63 | 84 | 320 | 326 | 352 | 311 | 0.42 | 0.92 |
| rs7966993 | 12 | 27554420 | PPFIBP1 | 33 | 45 | 240 | 246 | 462 | 430 | 0.80 | 0.22 |
| rs256662 | 12 | 30804286 | AC010198.2 | 35 | 34 | 237 | 255 | 461 | 432 | 0.52 | 0.64 |
| rs7301387 | 12 | 30807504 | AC010198.2 | 31 | 35 | 221 | 243 | 483 | 443 | 0.37 | 0.82 |
| rs256679 | 12 | 30809469 | AC010198.2 | 146 | 167 | 346 | 367 | 242 | 187 | 0.27 | 0.61 |
| rs12873430 | 13 | 26971307 | AL160035.1 | 8 | 9 | 122 | 128 | 605 | 584 | 0.51 | 0.51 |
| rs12875586 | 13 | 26971748 | AL160035.1 | 1 | 4 | 73 | 75 | 661 | 642 | 0.49 | 0.27 |
| rs9581791 | 13 | 26973204 | RPS21P8 | 81 | 58 | 312 | 272 | 342 | 391 | 0.44 | 0.27 |
| rs2421035 | 13 | 26978564 | RPS21P8 | 8 | 9 | 111 | 102 | 616 | 610 | 0.24 | 0.05 |
| rs9512476 | 13 | 26979157 | RPS21P8 | 86 | 122 | 333 | 354 | 315 | 244 | 0.89 | 0.74 |

| | | | | | | | | | | | |
|---|---|---|---|---|---|---|---|---|---|---|---|
| rs2824253 | 21 | 17171821 | - | 125 | 133 | 367 | 338 | 243 | 250 | 0.50 | 0.32 |
| rs13047117 | 21 | 17177871 | - | 72 | 55 | 322 | 274 | 341 | 392 | 0.75 | 0.46 |
| rs1501811 | 21 | 17201098 | - | 155 | 106 | 338 | 326 | 242 | 289 | 0.07 | 0.37 |
| rs1604416 | 21 | 17206922 | NEK4P1 | 185 | 133 | 351 | 344 | 194 | 223 | 0.30 | 0.99 |

**Table S4. Haplotype marker characteristics. H**omozygous minor allele counts for the case and control populations are defined under columns E and F; heterozygous allele count are listed under columns G and H; homozygous major allele counts are listed under columns I and J; hardy-weinberg p values for the case and control populations are defined under columns K and L.
