## Supplemental Figures for "Retrospective analysis of *The Two Sister Study* using haplotype-based association testing to identify loci associated with early-onset breast cancer"

*Funding*: The work described within this study was funded through the Children's Fund of Children's Hospital of Pittsburgh of UPMC and through the Ross H. Musgrave Endowment (J.E.L).

*Conflicts of Interest*: There are no conflicts of interest to disclose.

*Corresponding Author*:

Dr. Gregory M. Cooper
Department of Plastic Surgery
3533 Rangos Research Building
530 45th St, Pittsburgh, PA 15201
412/692-5384 (office)


*Keywords*: young-onset; early-onset; cancer; familial; breast cancer

### SUPPLEMENTAL INFORMATION

#### SUPPLEMENTAL FIGURE LEGENDS

**Figure S1. LOESS curve based upon age-at-diagnosis in *The Two Sister Study* breast cancer population.** The optimal cutoff for distinguishing between younger and older patients affected by breast cancer is calculated to be 45 years-of-age at the time of diagnosis. Martingale residuals plotted against age-at-diagnosis suggest that age-at-diagnosis is a discontinuous variable. The assumption of linearity is not fulfilled.

**Figure S2. Quantile-quantile plot.** An initial haplotype-based analysis was performed using a static 10 kb window to identify regions of interest. A QQ plot was constructed by plotting -log[observed p] versus -log[expected p]. The dashed line depicts an ideal line with a slope of 1. Note that individual data points closely follow the theoretical ideal with a slight tail becoming evident at the outermost extremes of the plot.

**Figure S3. Haplomaps representing candidate regions identified by haplotype trend regression with full scan permuted p values ≤ 0.05.** Images include chromosomal position using GRCh38 coordinates. Affected genes are defined beneath each haplomaps and solid black lines represent non-coding introns. Coding exons and their relative position within the displayed haplomaps are represented by solid black blocks. No gene is associated with the block displayed in **Fig S3g** or **Fig S3n**.

FIGURES

**Figure S1. Statistical testing warrants dichotomization of *The Two Sister Study* breast cancer population by age-at-diagnosis.**

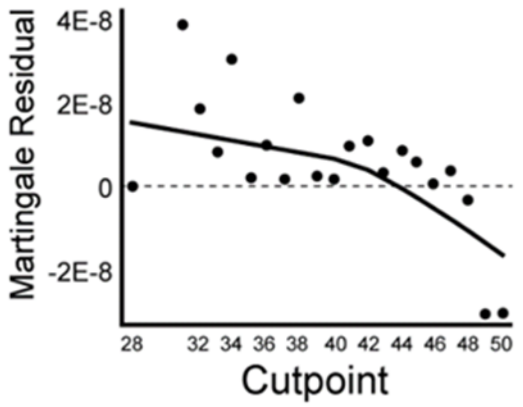


**Figure S2. Using age-at-diagnosis to dichotomize breast cancer patients yields normally distributed data.**

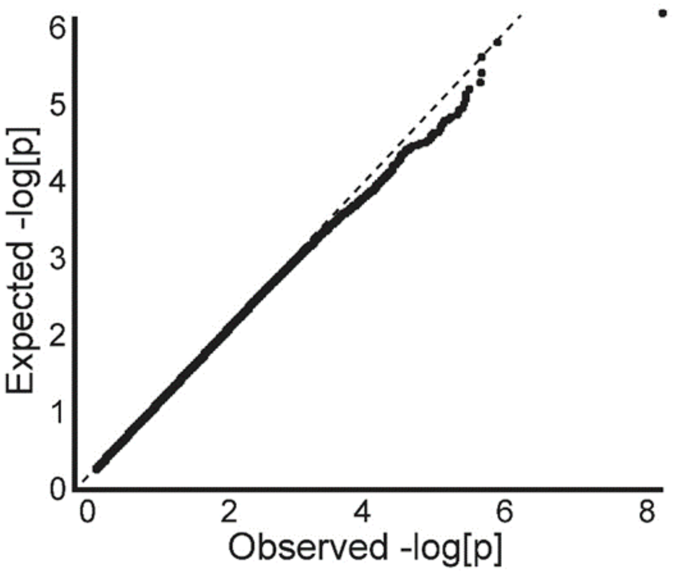

**Figure S3. Mapping of the most significant haploblocks defined by haplotype trend regression provides supporting evidence of underlying haplotype structure within candidate regions.**

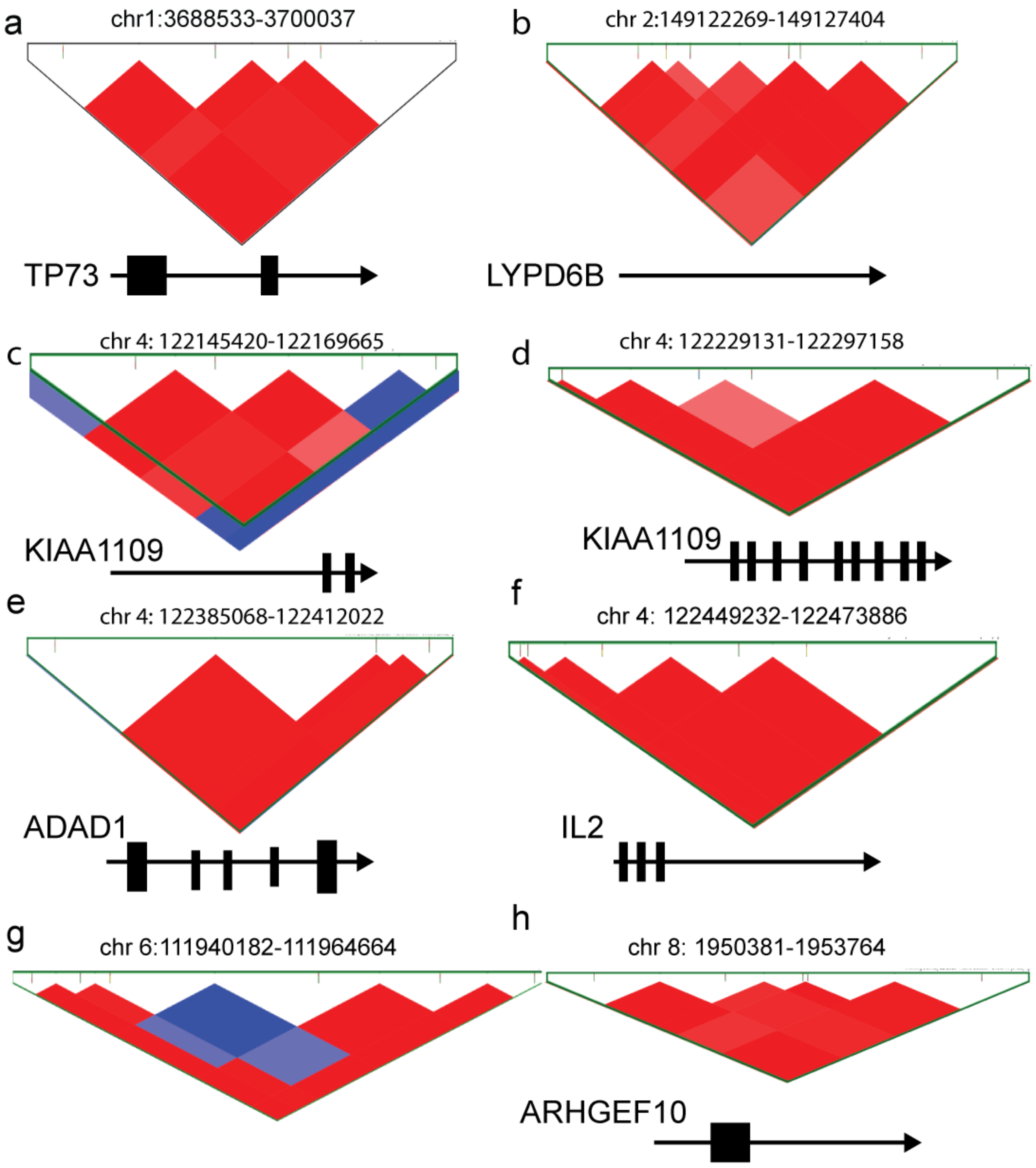

**Figure S3, continued.**

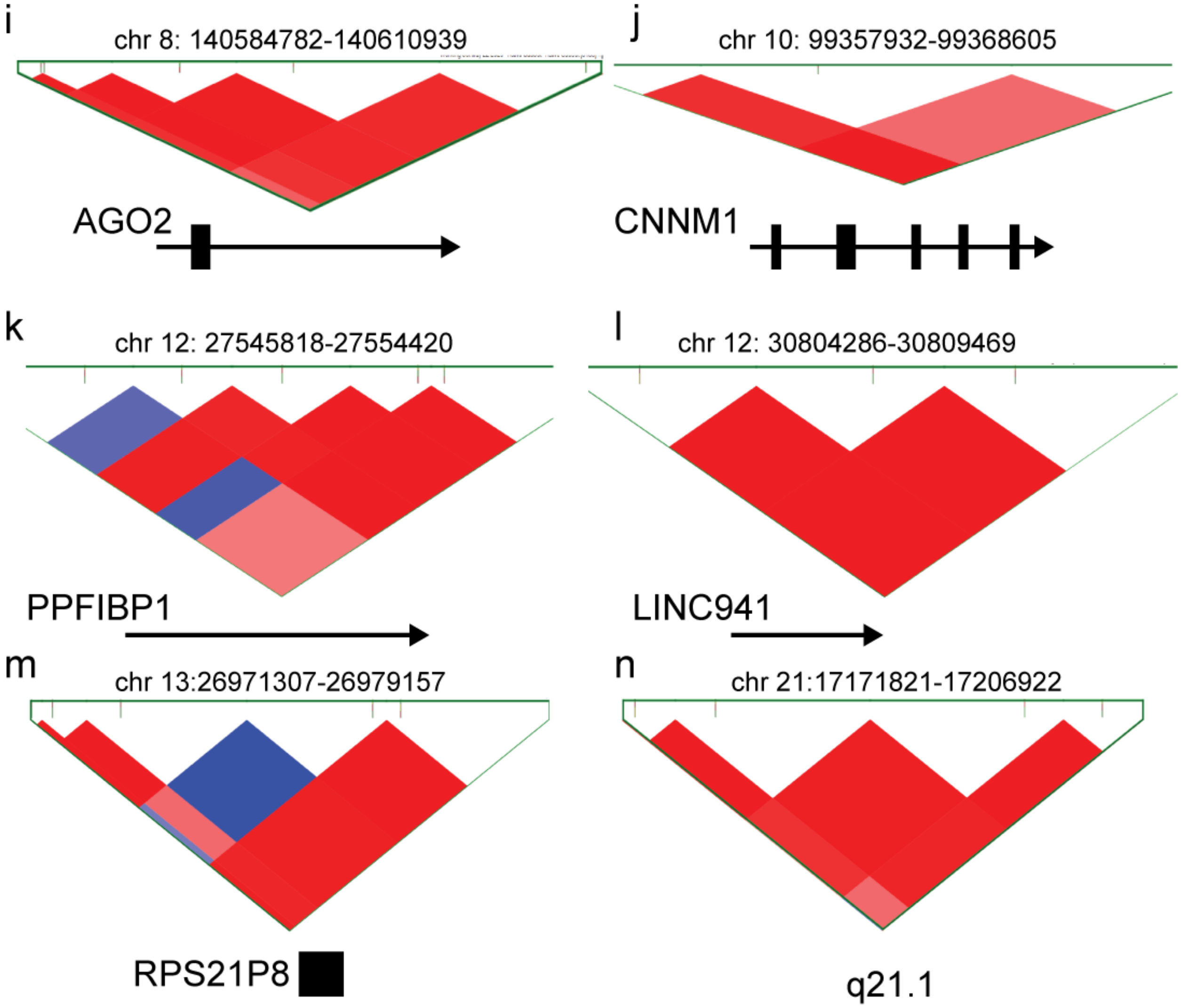
